# Behaviour, booster vaccines and waning immunity: modelling the medium-term dynamics of SARS-CoV-2 transmission in England in the Omicron era

**DOI:** 10.1101/2021.11.22.21266584

**Authors:** Rosanna C. Barnard, Nicholas G. Davies, Centre for Mathematical Modelling of Infectious Diseases COVID-19 working group, Mark Jit, W. John Edmunds

**Affiliations:** Centre for Mathematical Modelling of Infectious Diseases, London School of Hygiene & Tropical Medicine, Keppel Street, London, WC1E 7HT, UK; Department of Infectious Disease Epidemiology, London School of Hygiene & Tropical Medicine, Keppel Street, London, WC1E 7HT, UK

## Abstract

England has experienced a heavy burden of COVID-19, with multiple waves of SARS-CoV-2 transmission since early 2020 and high infection levels following the emergence and spread of Omicron variants since late 2021. In response to rising Omicron cases, booster vaccinations were accelerated and offered to all adults in England. Using a model fitted to more than 2 years of epidemiological data, we project potential dynamics of SARS-CoV-2 infections, hospital admissions and deaths in England to December 2022. We consider key uncertainties including future behavioural change and waning immunity, and assess the effectiveness of booster vaccinations in mitigating SARS-CoV-2 disease burden between October 2021 and December 2022. If no new variants emerge, SARS-CoV-2 transmission is expected to decline, with low levels remaining in the coming months. The extent to which projected SARS-CoV-2 transmission resurges later in 2022 depends largely on assumptions around waning immunity and to some extent, behaviour and seasonality.

## Main

Over two years into the COVID-19 pandemic, more than 500 million confirmed cases and 6 million deaths have been attributed to severe acute respiratory syndrome coronavirus 2 (SARS-CoV-2) worldwide^1^. In England as of 12th May 2022, cumulative confirmed COVID-19 cases exceed 18 million, with more than 700,000 hospitalisations and 150,000 deaths within 28 days of a positive test being recorded, respectively^2^. Different variants of SARS-CoV-2 have emerged, with five (Alpha, Beta, Gamma, Delta and Omicron) currently designated as variants of concern (VOC) associated with either increased transmissibility, severity, or changes in immunity by the World Health Organisation^3^.

England saw the emergence and fixation of the Alpha B.1.1.7 variant in late 2020, which was subsequently overtaken by the Delta B.1.617.2 variant in Spring 2021, followed by the Omicron B.1.1.529 variant in late 2021 and the Omicron BA.2 sublineage in early 2022. Various public health and social measures (PHSMs) have been implemented and relaxed at different times to control SARS-CoV-2 transmission in England, including national lockdowns, staged relaxations of lockdowns, tiered regional restrictions^4^, and so-called ‘Plan B’ measures which were introduced in response to Omicron’s emergence. Following the large wave of Omicron transmission beginning in late 2021, all legal restrictions in England were lifted on 24th February 2022^5^.

Safe and effective COVID-19 vaccines have been developed at unprecedented speed, with six currently approved for use by the UK’s Medicines and Healthcare products Regulatory Agency^6^. The COVID-19 vaccine rollout in England began on the 8th of December 2020 and to date more than 44 million people have received at least their first COVID-19 vaccine dose^2^. The rollout of vaccines followed guidance issued by the UK’s Joint Committee on Vaccination and Immunisation (JCVI)^7^, with vaccines being targeted to health and social care workers and those in the highest risk categories first. Vaccines were then offered to sequentially younger age groups of adults (18 years and above). In August 2021, children aged 16 and 17 years old and clinically vulnerable children aged 12-15 were offered COVID-19 vaccines^8^. In September 2021, healthy 12-15-year-olds in England were offered their first COVID-19 vaccination and from April 2022 vaccination has been extended to 5-11 year olds^9^.

A COVID-19 booster vaccination programme began in England in September 2021, initially targeting the same highest-risk priority groups that were first vaccinated. A full dose of the Pfizer/BioNTech or a half dose of the Moderna vaccine are recommended as a booster dose, regardless of what vaccine was received previously, to those at least 6 months after their primary course of vaccination. On 15th November 2021, the JCVI issued advice recommending that the widespread COVID-19 booster vaccination programme be extended to individuals aged 40-49 years^10^. However, following the World Health Organization (WHO) designating the Omicron SARS-CoV-2 variant as a variant of concern in late November 2021^11^, with Omicron cases being detected in England^12^ and with two vaccine doses found to offer little protection against Omicron a few months after vaccination^13^, the JCVI announced an extension and acceleration of the booster vaccination campaign^14^. This announcement recommended that 18-39 year olds should be offered booster vaccinations in descending age order, that the minimum gap between the primary vaccination course and the booster vaccine should be reduced from 6 to 3 months, and that children aged 12-15 years of age should receive their second vaccine dose^15^. The JCVI also recommended that children aged 16 and 17 years of age could receive booster vaccinations at least 3 months after completion of their primary vaccination course^16^. Reduced dose COVID-19 vaccinations were offered to the most vulnerable 5-11 year old children from the end of January 2022^17^, and in February 2022 the JCVI recommended a non-urgent extension of this rollout to children aged 5-11 who are not in a clinical risk group^18^. Given the high levels of Omicron transmission in England since late 2021, emergence of the BA.2 sublineage, the accelerated booster vaccination campaign, and evidence suggesting that vaccine protection wanes over time^19, 20^, it is important to assess the likely medium-term dynamics of SARS-CoV-2 transmission as the Omicron wave subsides and as, in the absence of legal restrictions, behaviour returns to a (potentially new) baseline.

Using an age-structured deterministic compartmental model of SARS-CoV-2 community transmission, we explore the consequences of key uncertainties on projected COVID-19 cases, hospitalisations and deaths to December 2022. We fit the model to regional data up to May 2022 on COVID-19 deaths, hospitalisations, and hospital bed occupancy, as well as PCR prevalence, seroprevalence, and the emergence of the Alpha B.1.1.7, Delta B.1.617.2, and Omicron B.1.1.529 VOCs. We also integrate data on: the number of vaccinations delivered at the level of NHS England regions and by age group over time, historic school attendance and community mobility data to inform behavioural changes over time since the start of the COVID-19 pandemic, and genomic sequencing data to inform the proportion of Omicron cases attributable to the BA.1 and BA.2 sublineages over time. We consider the effects of future behaviour change, waning immunity, seasonality, and vaccination of children and adolescents at different levels of uptake on potential future dynamics of SARS-CoV-2 transmission in England to September 2022. We also assess the effectiveness of COVID-19 booster vaccinations by exploring counterfactual booster vaccination scenarios and their effect on SARS-CoV-2 transmission in England between October 2021 and December 2022.

## Results

### Model fitting & uncertainty

Our compartmental model (**Fig. 1**) fits the observed dynamics of SARS-CoV-2 community transmission during the COVID-19 epidemic in England between mid-February 2020 and May 2022 (**Figs. 2, S1A and S1B**), reproducing NHS England region-specific observed deaths, hospitalisations, hospital and intensive care unit (ICU) bed occupancy, PCR prevalence, and seropositivity. Following the initial differential evolution Markov chain Monte Carlo (DE-MCMC) fitting, the model applies continuous-time multipliers to the infection fatality, hospital admissions, hospital bed occupancy and the ICU bed occupancy rates for each region in order to match real-world measured outcomes (**Fig. S1C**). There are notable peaks in the fatality rate adjustment corresponding to the initial SARS-CoV-2 wave in early 2020 and to the Alpha B.1.1.7 wave in the winter of 2020-21 (**Fig. S1C**). All four rate adjustments have reached low levels in early 2022; we carry forward the last adjustment for forward projections (**Fig. S1C**).

**Figure 1.**
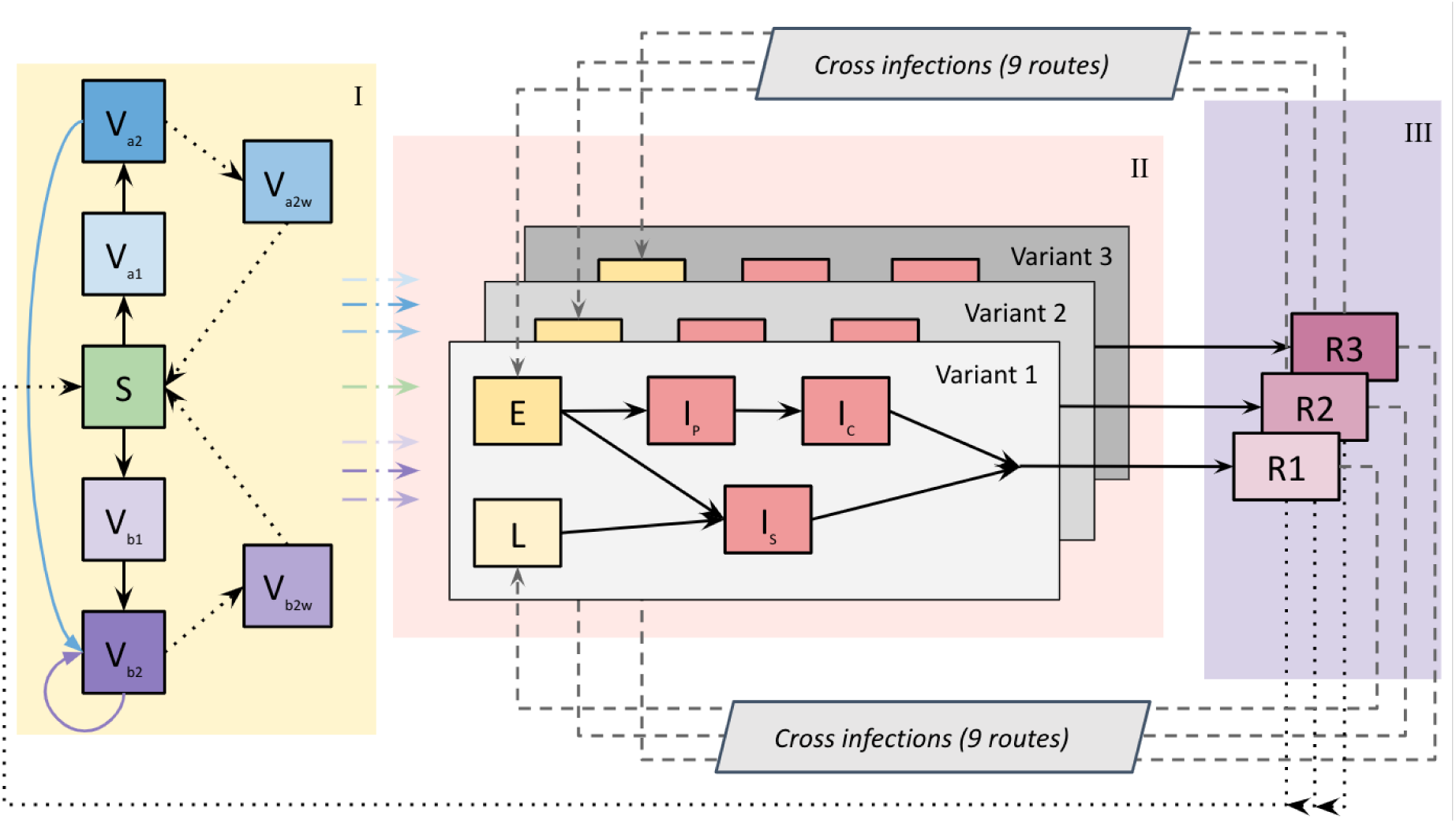
Compartmental model diagram. A three-variant deterministic dynamic compartmental model with vaccination describes SARS-CoV-2 transmission in England. We model seven NHS England regions^25^ separately, with each divided into 16 five-year age groups: 0-4 years up to 70-74 years, and 75 years and older. The model incorporates COVID-19 vaccination with two vaccine products (corresponding to the viral-vector (Va) and mRNA-based (Vb) vaccines in use in England), each with first- (Va1, Vb1) and second-dose (Va2, Vb2) protection, and each with lower levels of protection for individuals who received their primary vaccinations but no booster vaccine and have waned (Va2w, Vb2w). All vaccinated individuals have increased protection against different SARS-CoV-2 outcomes compared to susceptible individuals, according to the vaccine product administered and their vaccine dose/waned status (**Table S2**). Vaccinated individuals transition from the susceptible (S) compartment into first-dose vaccinated compartments (Va1, Vb1) depending on which vaccine product was received. Following an assumed first-dose duration (**Table S5B**), individuals move into second-dose compartments (Va2, Vb2). Following an assumed second-dose duration (**Table S5B**), individuals either receive a booster vaccine or transition into waned states (Va2w/Vb2w). We assume that individuals receiving primary courses of a viral-vector vaccine (Va2) or an mRNA vaccine (Vb2) both move to the Vb2 compartment following their booster vaccination, with their second-dose duration beginning again from zero (**Table S5B**). This assumptions reflects the fact that all booster vaccinations in England are either the Pfizer/BioNTech or Moderna mRNA vaccines, and evidence finding higher immunogenicity for individuals receiving Pfizer/BioNTech following Oxford-AstraZeneca, compared with individuals receiving both Oxford-AstraZeneca vaccine doses^26^. We model an additional temporary increase in vaccine protection for individuals receiving their first booster vaccine in late 2021/early 2022, which lasts for 180 days. We utilise three separate SARS-CoV-2 variants in the model to capture the introduction and spread of the Alpha, Delta and Omicron variants of concern in England. The model assumes a traditional infection process: upon being infected, individuals leave the susceptible (S), vaccinated (V) or recovered (R) states and move through exposed (E), infectious (I) and recovered (R) states. The latent (L) state is used in addition to the exposed (E) state for breakthrough infections following vaccination and for re- and cross-infections, to achieve additional vaccine protection against disease (**Tables S2, S3**). When individuals are infectious (I), they either progress through a subclinical (Is) pathway or a clinical pathway with pre-clinical (Ip) and clinical (Ic) states. Once individuals have been infected and recover (R), we allow for loss of immunity (where individuals return to a susceptible (S) state) and re- and cross-infections (where individuals with immunity become infected, see **Table S3**). On the left hand side (yellow shaded background labelled I), solid black arrows represent primary vaccinations, solid coloured arrows represent booster vaccinations, and dotted black arrows represent loss of immunity. Coloured dash-dotted arrows denote susceptible and vaccinated individuals becoming infected and moving into the SARS-CoV-2 infection process (grey boxes on red shaded background labelled II). Here, solid black arrows denote individuals moving through the infection process and recovering (R) (purple boxes on purple shaded background labelled III). Recovered individuals can lose their immunity (dotted black arrows) and return to the susceptible disease state (S) (**Table S4**) or be re- or cross-infected (dashed grey arrows) with other SARS-CoV-2 variants (**Table S3**).

**Figure 2.**
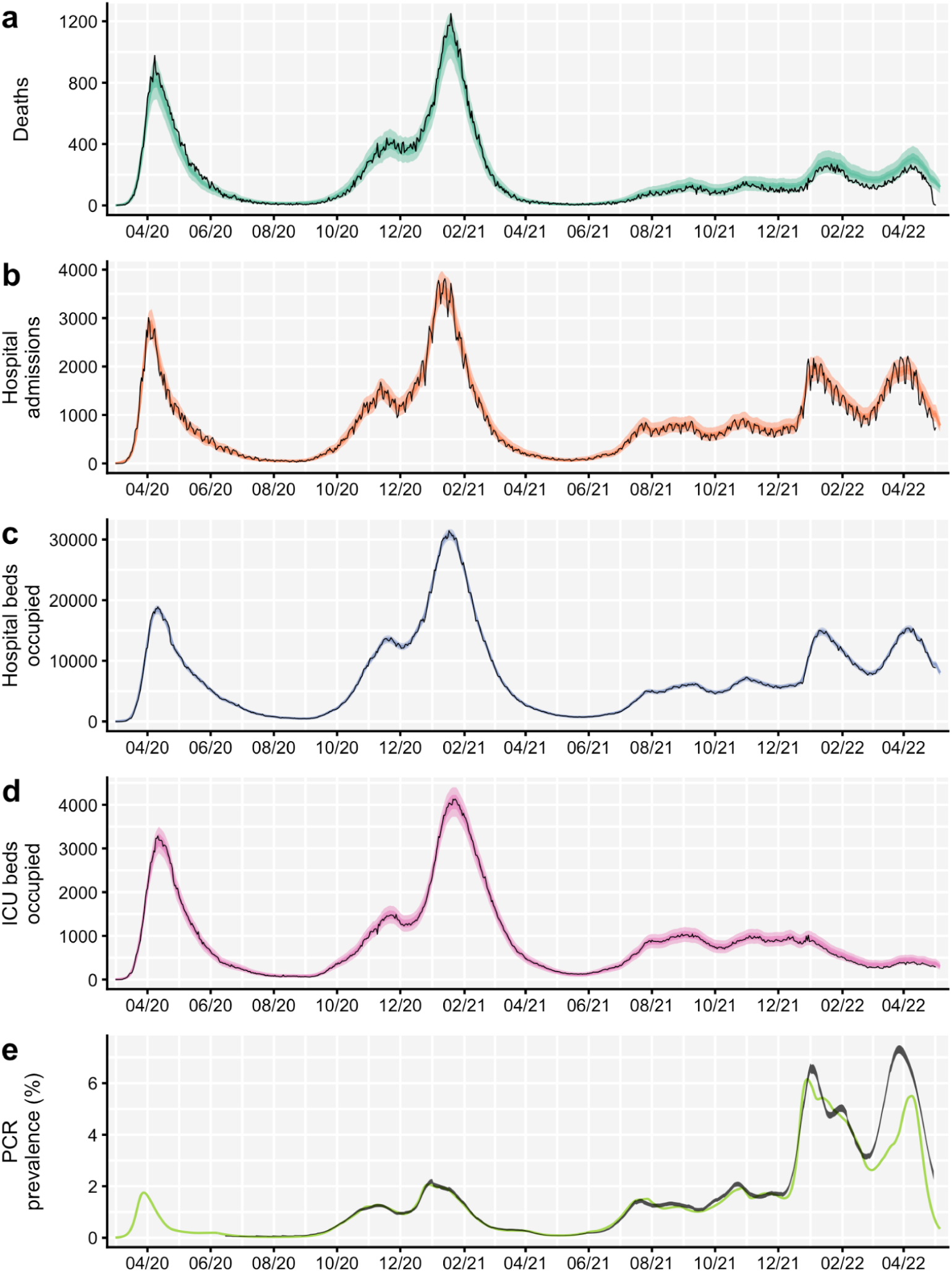
Comparison between aggregated model fits and epidemiological data from England between March 2020 and May 2022. Black lines show reported data, with black ribbons showing 95% confidence intervals for PCR prevalence. Coloured lines and shaded areas show medians, 50% and 90% interquantile ranges from the fitted model. The original model fitting is done independently for each NHS England region (see **Figs. S1A-B**), with the aggregated model output for the whole of England shown here. **(a)** COVID-19 deaths over time, where data was provided by the UK Health Security Agency (UKHSA). **(b)** COVID-19 hospital admissions over time, where data was provided by NHS England. **(c)** COVID-19 hospital bed occupancy over time, where data was provided by NHS England. **(d)** COVID-19 ICU bed occupancy over time, where data was provided by NHS England. **(e)** COVID-19 PCR prevalence over time, where publicly-available PCR prevalence data was obtained from the Office for National Statistics’ COVID-19 Infection Survey (ONS-CIS)^27^. The data sources for COVID-19 deaths, hospital admissions, hospital and ICU bed occupancy are unpublished and not publicly available, but are closely aligned with the UK Government’s COVID-19 dashboard^2^. ICU = intensive care unit. NHS = National Health Service.

The model also captures the emergence and spread of the Alpha B.1.1.7, Delta B.1.617.2, and Omicron B.1.1.529 VOCs in late 2020, early 2021, and late 2021, fitting to the prevalence of S gene target failure (**Figs. S2a and S2c**) as a proxy for the proportion of cases attributable to the Alpha and Omicron variants, and to the proportion of Delta sequenced cases (**Fig. S2b**). Model estimates for increased transmissibility of Alpha relative to previously circulating SARS-CoV-2 variants, of Delta relative to Alpha, and of the earlier Omicron BA.1 sublineage relative to Delta, as well as the overall estimated increases in transmissibility between early SARS-CoV-2 variants and each VOC and Omicron sublineage are given in **Tables S1A and S1B**, respectively. Note that we assume that the Omicron BA.2 sublineage confers a 50% increase in transmissibility compared to the previous Omicron BA.1 sublineage.

To capture historic behavioural changes, the model uses Google Community Mobility indices^21^ over time to derive contact rates for each NHS England region modelled, based upon a measured relationship between Google Mobility indices and age-specific contact rates as measured by the CoMix study^22^, and in combination with school attendance data^23^ and assumptions about school terms (**Fig. 3**). The model also fits a time-varying “transmission adjustment” component for each NHS England region in order to capture additional variability in transmission that is not explained by the mobility data (**Fig. 3**, middle row; Online Methods). To project behavioural changes forwards from May to December 2022, we combine various assumptions on future mobility changes (**Fig. 3**) with simulated trajectories for future transmission adjustments based on the historic fitted transmission adjustment (full details are given in the Online Methods section).

**Figure 3.**
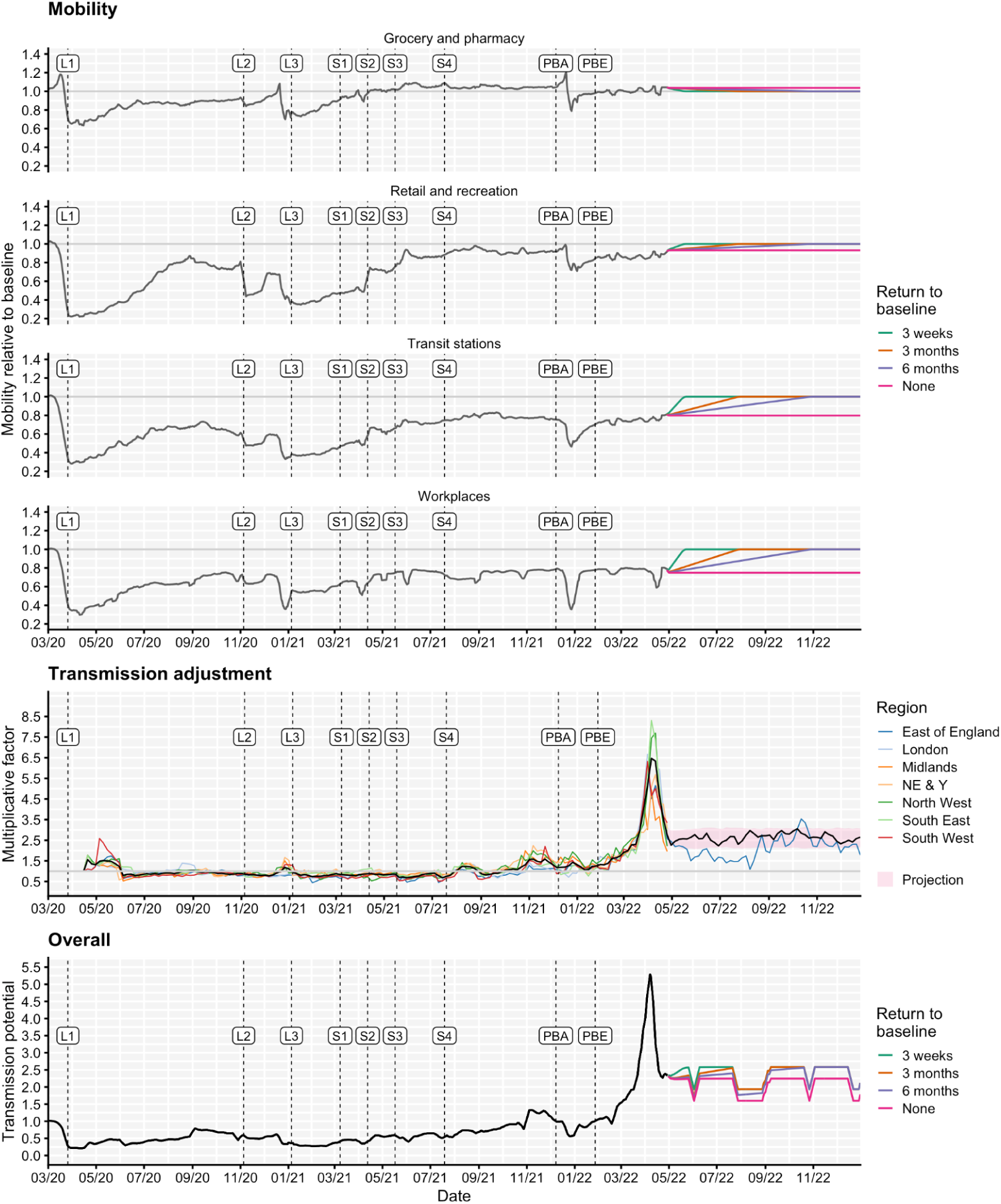
Mobility scenarios, transmission adjustments and overall transmission potential for the fitted model, shown from March 2020 to December 2022. Top: Historic Google Community Mobility data^21^ (grey) and assumed future mobility in England for no change (pink), a 3-week return to pre-pandemic baseline levels (green), a 3-month return to pre-pandemic baseline levels (orange) and a 6-month return to pre-pandemic baseline levels (purple) scenarios used for model projections. Mobility indices are measured relative to baseline mobility levels recorded during early 2020, prior to the COVID-19 pandemic. The beginning of each lockdown and each roadmap Step^4^ is marked with a vertical dashed line and ‘L’ and ‘S’ labels, respectively. Vertical dashed lines with ‘PBA’ and ‘PBE’ labels correspond to the announcement of ‘Plan B’ measures for England on 8th December 2021 and the ending of these measures on 27th January 2022^28^. Middle: Fitted transmission adjustments between April 2020 and May 2022 by NHS England region (coloured lines) and the average across regions (black line), example projection between May and December 2022 for East of England (blue line) and mean (black line) and interquartile range (red shaded) for projected transmission adjustments between May and December 2022 across NHS England regions. Bottom: The overall “transmission potential” captures the combined impact of mobility and transmission adjustments on the time-varying potential for effective transmission, ignoring the impact of immunity and novel variants, though including the impact of school vacation periods. NE & Y = North East & Yorkshire. NHS = National Health Service.

We present the majority of results here by plotting the median and interquantile ranges of a number of simulated future trajectories of SARS-CoV-2 transmission in England, but it is important to note that individual epidemic trajectories can fall outside of the model’s projection intervals (**Fig. 4c**). A comparison of the projected cumulative number of SARS-CoV-2 infections, hospital admissions and deaths between May and December 2022 across all scenarios considering future uncertainties (namely, behaviour, waning immunity, seasonality and vaccination uptake for children aged 5 and above) is shown in **Figure 5**. Detailed results related to each type of uncertainty considered are given in the Supplementary material (**Figs. S3, S4A-B, S5-S8**). A complete list of scenarios considered and key assumptions for each scenario is given in **Table 1**.

**Table 1.** List of modelling scenarios and key assumptions. Scenarios marked with an asterisk (*) are equivalent. Mobility scenarios are shown in **Figure 3**. Assumptions for waning scenarios are given in Table S4. Assumptions for vaccine effectiveness against different SARS-CoV-2 variants are given in Table S2. Assumptions for cross-protection against different SARS-CoV-2 variants given prior infection are given in Table S3.

| Scenario | Result type | Assumptions |  |  |  |  |
| --- | --- | --- | --- | --- | --- | --- |
|  |  | Mobility | Waning | Booster uptake | Seasonality | Vaccine policy |
| Basecase* | - | 6-month | Central | Actual | 20% | 5+ 80% uptake |
| No return | Behaviour | No change | Central | Actual | 20% | 5+ 80% uptake |
| 6-month return* |  | 6-month | Central | Actual | 20% | 5+ 80% uptake |
| 3-month return |  | 3-month | Central | Actual | 20% | 5+ 80% uptake |
| 3-week return |  | 3-week | Central | Actual | 20% | 5+ 80% uptake |
| No boosters | Boosters eligibility / uptake (relative to second dose levels) | 6-month | Central | None | 20% | 5+ 80% uptake |
| 50+ boosters |  | 6-month | Central | 95% in 50+ only | 20% | 5+ 80% uptake |
| Actual boosters* |  | 6-month | Central | Age-specific measured uptake as of 14th April 2022<sup>†</sup>. Individuals aged 50 and above have at least 86.1% and at most 96.7% uptake. Individuals aged 15-49 have at least 40% and at most 80.9% uptake, relative to second dose levels of uptake. | 20% | 5+ 80% uptake |
| High uptake |  | 6-month | Central | 90% in <50s; 98% in 50+ | 20% | 5+ 80% uptake |
| Central waning* | Waning | 6-month | Central | Actual | 20% | 5+ 80% uptake |
| High waning |  | 6-month | High | Actual | 20% | 5+ 80% uptake |
| Very high waning |  | 6-month | Very high | Actual | 20% | 5+ 80% uptake |
| 10% | Seasonality | 6-month | Central | Actual | 10% | 5+ 80% uptake |
| 20%* |  | 6-month | Central | Actual | 20% | 5+ 80% uptake |
| 30% |  | 6-month | Central | Actual | 30% | 5+ 80% uptake |
| 40% |  | 6-month | Central | Actual | 40% | 5+ 80% uptake |
| 5+, 80%* | Vaccine uptake for children | 6-month | Central | Actual | 20% | 5+ 80% uptake |
| 5+, 50% |  | 6-month | Central | Actual | 20% | 5+ 50% uptake |

<sup>†</sup>The detailed age-specific probability of receiving a booster vaccine for this scenario is listed in **Table S5B**.

**Figure 4.**
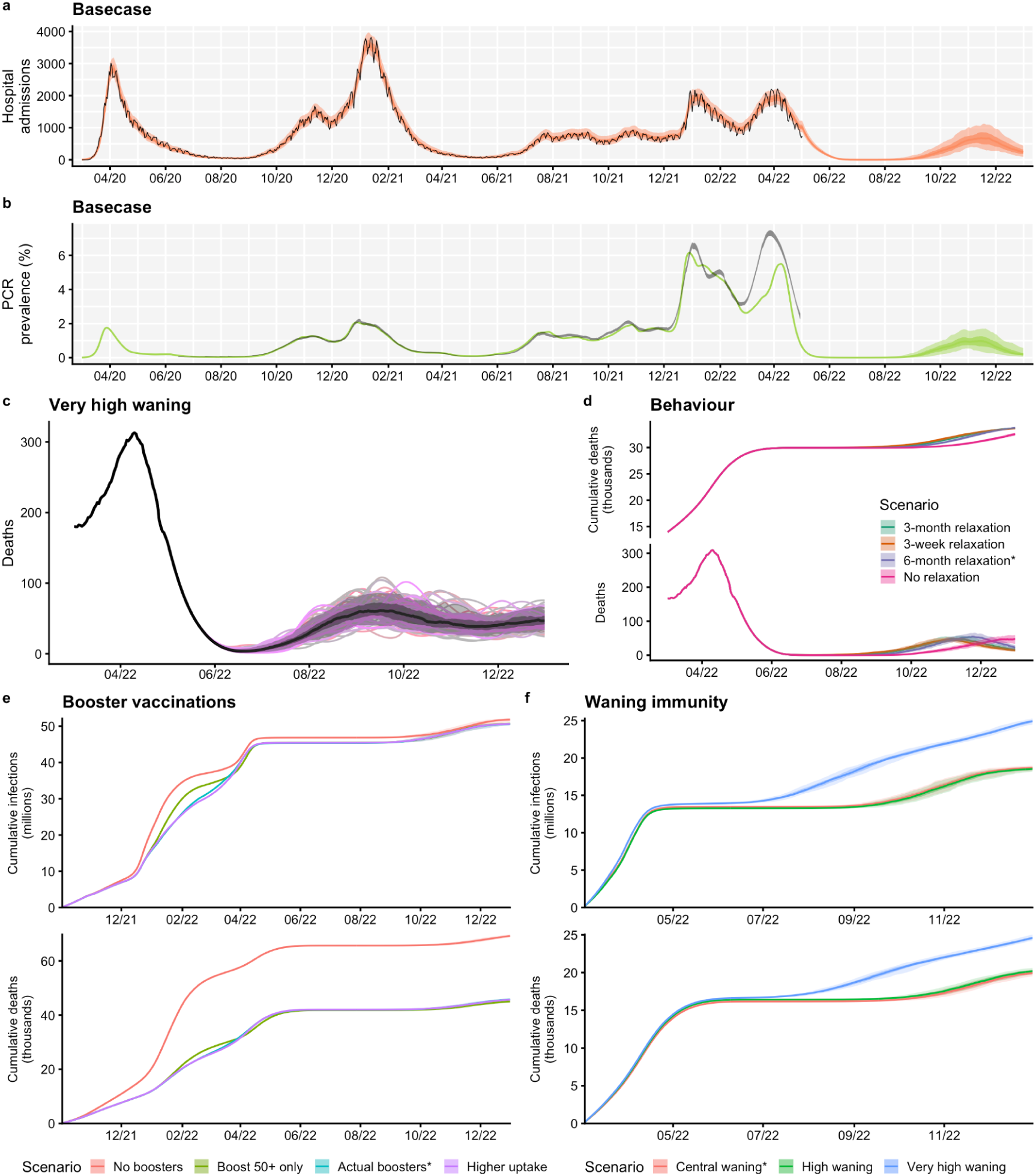
Summary of basecase model fits and projections and key results on uncertainty, behaviour, booster vaccinations and waning immunity. **(a)** The number of COVID-19 hospital admissions in England, for the basecase scenario, between March 2020 and December 2022. Black lines show reported data, provided by UKHSA. Coloured lines and shaded areas show medians, 50% and 90% interquantile ranges from the fitted model and from the model projection. (**b)** PCR prevalence in England, for the basecase scenario, between March 2020 and December 2022. The black ribbons show 95% confidence intervals for PCR prevalence data. Coloured lines and shaded areas show medians, 50% and 90% interquantile ranges from the fitted model and from the model projection. (**c)** The fitted and projected number of COVID-19 deaths in England between March and December 2022, shown for the very high waning scenario (see **Table S4**). The black line shows the median trajectory of COVID-19 deaths in England over time, with the shaded areas showing the 50% and 90% interquantile ranges. Individual model trajectories are plotted in coloured lines. (**d)** The effect of future behaviour on COVID-19 deaths and cumulative deaths (thousands) over time is shown with four scenarios for future mobility: a 3-week, a 3-month and a 6-month return to baseline levels, and a no change scenario (see **Table 1**). (**e**) The effect of booster vaccination policy on cumulative infections and deaths since October 2021 is shown with four scenarios for booster policies (**Table 1**). (**f**) The effect of waning immunity on cumulative infections and deaths between March and December 2022 is shown with three scenarios for waning (**Tables 1, S4**). The basecase scenarios (shown in panels **a** and **b**) and scenarios marked with an asterisk (*) are equivalent.

**Figure 5.**
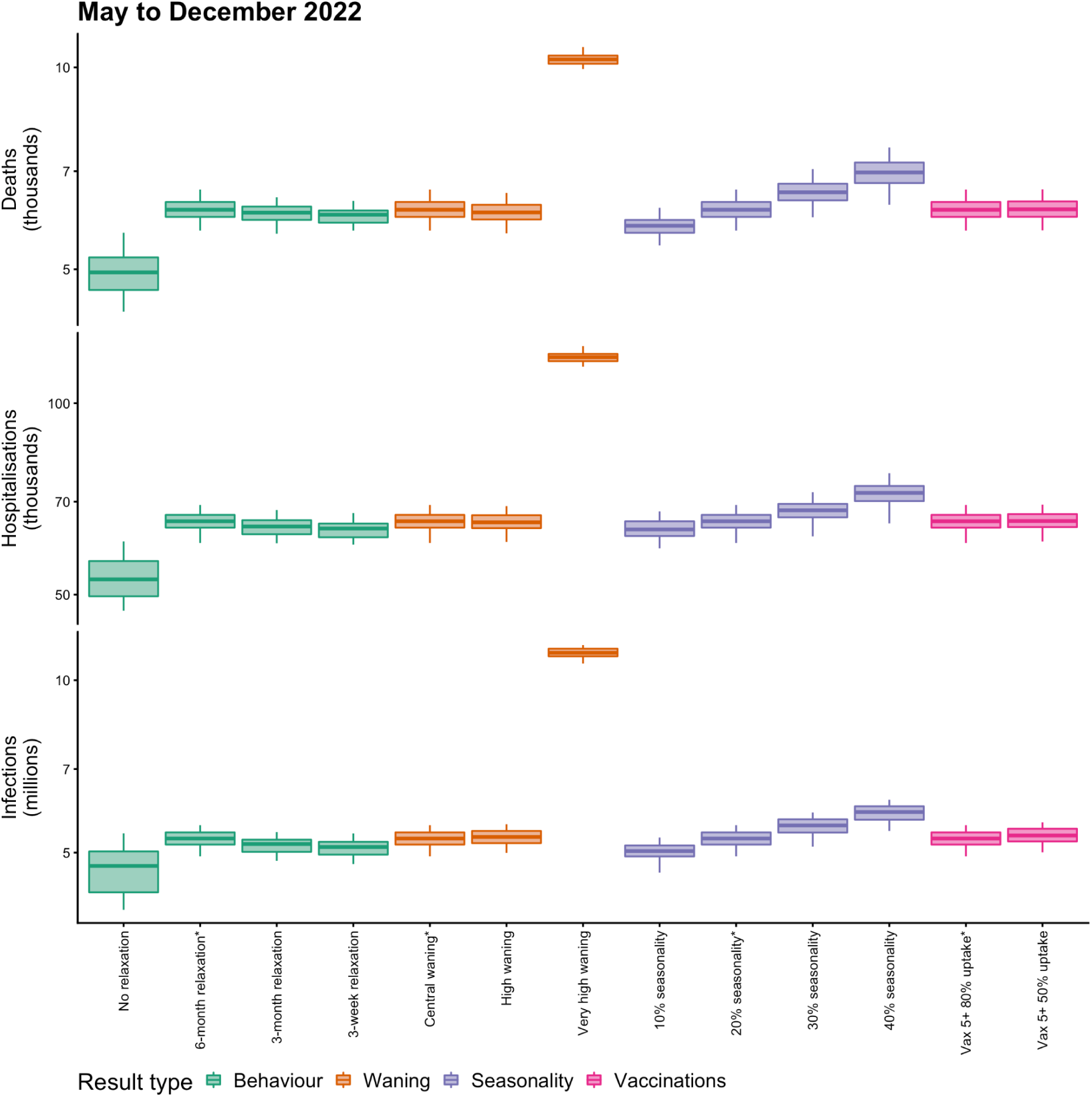
Summary of projected cumulative numbers (log-scale) of COVID-19 deaths (thousands), hospital admissions (thousands) and infections (millions) in England between May and December 2022, across behavioural, waning, seasonality and vaccination scenarios considered. Each box plot shows the projected median, 5th, 25th, 75th and 95th percentile values across all simulations for the relevant scenario, calculated between May and December 2022. Scenarios are coloured according to the result type (from left to right: behaviour, waning immunity, seasonality, and vaccination policies for children aged 5 years and older). A full list of scenarios and relevant modelling assumptions is given in **Table 1**. Scenarios marked with an asterisk (*) are equivalent and correspond to the basecase scenario. N.B. The y-axes are plotted on a log scale, and are truncated and do not extend to zero.

### Basecase scenario

Following the Omicron wave of transmission beginning in late 2021, our basecase scenario projects a continued reduction in transmission until August 2022, with a resurgence in transmission from August 2022 onwards (**Figs. 4a, 4b**). This period of low infection incidence results from very high levels of immunity derived from both vaccination and prior infection within the population in early 2022 following the Omicron wave of infection (**Fig. S8**), and assumes that no novel variants of SARS-CoV-2 emerge and outcompete the current Omicron BA.2 sublineage.

### Future behaviour

Assumptions about future levels of mobility have a small effect on projected dynamics of SARS-CoV-2 transmission in 2022 (**Figs. 4d, 5, S3**). We consider four scenarios for future mobility following measured levels on 29th April 2022 (no change versus a 3-week, 3-month or 6-month return to pre-pandemic baseline levels, shown in **Figure 3**). All behavioural change scenarios considered project a reduction in SARS-CoV-2 transmission to very low levels during the summer of 2022 (**Fig. S3**). However, the timing and extent of the projected resurgence later in 2022 depend on the assumed changes in future mobility (**Fig. S3**), with a later resurgence occurring if mobility remains at current levels. The timing and speed of the return to baseline mobility interacts with school terms and other modelling assumptions such as waning and seasonality. A 3-week return to baseline mobility results in the earliest projected resurgence, while a 6-month return to baseline mobility results in a later, but steeper resurgence in transmission towards December 2022 (**Fig. S3**).

### Booster vaccinations

An influential factor considered here is the availability and uptake of COVID-19 booster vaccinations (**Figs. 4e, S4A-B**). Here, we consider a number of scenarios exploring the effects of different booster vaccination rollouts on SARS-CoV-2 transmission in England between October 2021 and December 2022 (**Table 1**). A counterfactual scenario in which no COVID-19 booster programme was deployed has a large effect, with an additional 23,400 deaths and 108,000 hospital admissions between October 2021 and December 2022, compared to our basecase scenario using levels of booster vaccination uptake measured in April 2022 (**Fig. S4A Tables**).

There are minimal differences among the remaining three booster vaccination scenarios which consider age-specific booster uptake as measured on the 14th of April 2022, booster vaccines being administered to 95% of individuals aged 50 years and above only, and booster vaccines being administered to 90% of individuals aged 15-49 and 98% of individuals aged 50 years and above, representing higher booster uptake than was measured to date (**Table 1**). Burdens are similar across these three scenarios, with the scenario where booster vaccinations were only offered to individuals aged 50 and above resulting in higher levels of infections and deaths during the first wave of Omicron transmission in late 2021 to early 2022 (**Figs. 4e, S4A-B**).

### Waning immunity

The extent to which vaccine-induced and natural immunity wanes over time is a key driver of projected transmission dynamics in 2022 (**Figs. 4f, 5, S5**). We consider three waning immunity scenarios which apply to vaccinated individuals and to individuals who have recovered from a prior infection: basecase, high and very high waning (**Table S4**). All three of these scenarios assume booster vaccination uptake as measured in April 2022^24^. The basecase waning scenario is projected to result in a total of 5.29 million infections, 65,200 hospital admissions and 6,130 deaths between May and December 2022 (**Fig. S5 Tables**). The high waning scenario is projected to result in slightly more infections but slightly fewer hospital admissions and deaths, compared to the central waning scenario, over this time period. The very high waning scenario results in an earlier resurgence beginning in June 2022, with an additional 6 million infections, 52,800 hospital admissions and 4,170 deaths projected between May and December 2022, compared to the basecase waning scenario (**Fig. S5 Tables**).

### Seasonality

Different assumptions related to the extent of seasonality also influence the projected dynamics of SARS-CoV-2 transmission (**Figs. 5, S6**). Scenarios assuming the most extreme seasonal effects result in the largest cumulative burdens between May and December 2022 (**Figs. 5, S6**). However, scenarios assuming the least extreme seasonal effects result in resurgences in transmission earlier in 2022 than scenarios with greater seasonality (**Fig. S6**). To fully explore projected SARS-CoV-2 dynamics with different seasonal effects requires consideration of much longer time horizons than those considered in this study.

### Vaccination of children

We consider two scenarios for the vaccination of children aged 5 years and above between May and December 2022 (**Figs. 5, S7**). Of all the uncertainties considered here, using different assumptions related to the vaccination of children had the smallest overall effect on projected SARS-CoV-2 infection and disease until December 2022 (**Fig. 5**). The scenario assuming higher uptake of COVID-19 vaccinations in children aged 5 and above is projected to result in small reductions in infections, hospital admissions and deaths between May and December 2022 (**Fig. S7 Table**), but not to significantly alter projected transmission dynamics (**Fig. S7**). This is due to high levels of infection in the model in these age groups during the autumn of 2021 and winter 2021-22, leading to naturally acquired immunity (**Fig. S8**).

## Discussion

We have fitted a deterministic compartmental model of SARS-CoV-2 transmission to data between March 2020 and May 2022 on COVID-19 deaths, hospital admissions, hospital bed and ICU bed occupancy, PCR prevalence, seroprevalence, and the emergence and spread of the Alpha B.1.1.7, Delta B.1.617.2 and Omicron B.1.1.529 variants of concern, including the BA.1 and BA.2 sublineages of Omicron, and incorporating additional data on vaccination coverage and behaviour over time. Projecting forwards to December 2022, we have considered a number of future uncertainties around behaviour, waning immunity, seasonality, and vaccination of children aged 5 years and above, as well as retrospectively assessing the effectiveness of booster vaccinations on SARS-CoV-2 transmission in England.

Modelled levels of immunity in England as a result of the extensive COVID-19 primary and booster vaccination campaigns and following the recent wave of Omicron infections (**Fig. S8**) suggest that in the absence of further new variants or Omicron sublineages outcompeting the currently-dominant Omicron BA.2 sublineage, SARS-CoV-2 transmission will continue to fall in the next few months, remaining at low levels during the summer months of 2022. Our modelling suggests that higher SARS-CoV-2 transmission may resurge later in 2022, with the timing and extent of this resurgence largely dependent on our modelling assumptions related to waning immunity and, to some extent, future behaviour and seasonality. There remains significant uncertainty around both the extent and the timescale to which immunity wanes, as well as future behaviour and the speed and level that any return towards pre-pandemic baseline behaviours will reach.

Our modelling suggests that the COVID-19 booster vaccination campaign was highly effective in reducing transmission and in particular mitigating severe outcomes during recent months, with a counterfactual scenario without any booster vaccinations projected to result in a large peak of hospital admissions in England exceeding the peak levels recorded throughout the COVID-19 pandemic to date (**Fig. S4A**). Importantly, the model shows that this effect mostly derives from boosting immunity in those over 50 years of age, with little additional benefit being derived from boosting younger age groups. We also show that the projected dynamics of SARS-CoV-2 transmission depend on modelling assumptions about the extent of seasonal effects and that achieving higher levels of vaccination coverage for children aged 5 years and older will not significantly influence the epidemiological dynamics of SARS-CoV-2 in England until December 2022, primarily due to the late initiation of paediatric vaccination, high rates of natural immunity that has already accumulated (**Fig. S8**), the lower risk of severe disease, and lower inherent susceptibility to infection for this age group^29^.

Although a number of our modelling scenarios project resurgences in SARS-CoV-2 transmission occurring later in 2022, none of these are expected to reach the peak levels of infections, hospital admissions or deaths recorded so far during the COVID-19 pandemic in England, at least during the time periods we are considering. This is largely due to very high levels of vaccine and natural protection that have built up in the population, particularly following the recent Omicron BA.1 and BA.2 waves of infection, and which under our central waning immunity scenarios remain high through December 2022 (**Fig. S8**). However, it is important to note that we do not consider any projected SARS-CoV-2 dynamics past December 2022. Since many of the scenarios considered here appear to be resurging towards the end of the simulation time period, comparing overall burdens across these scenarios must therefore be done with caution. Moreover, although we consider differential contact-making across the 5-year age groups in the model, we do not explore any differences in contact-making behaviour within age-groups, such as for individuals in vulnerable or at-risk groups. These mixing assumptions may not be sufficient to capture all heterogeneities in behaviour that could lead to some individuals avoiding SARS-CoV-2 infection.

Furthermore, these modelling scenarios do not consider the introduction of any additional SARS-CoV-2 variants, or sublineages of Omicron aside from BA.1 and BA.2, which may possess characteristics such as increased transmissibility, immune evasion or increased pathogenicity relative to existing variants and sublineages circulating in the population. As we have seen in England, novel VOCs with transmission advantages can spread to dominance very quickly (**Fig. S2**), and in particular the introduction of a new VOC which evades existing immune protection can significantly alter the dynamics of an epidemic, even in a previously highly immune population (**Fig. S8**), as we have seen with the Omicron BA.1 sublineage. Since the earliest SARS-CoV-2 variants, we have observed both increases and reductions in the intrinsic pathogenicity of the virus as different VOCs have emerged and spread to dominance globally^30–33^. The currently-dominant Omicron VOC has reduced pathogenicity compared to the previously-circulating Delta VOC^32^, and these levels of severity are carried forward in our modelling projections. We do not explore the effects of any future change in intrinsic pathogenicity in this work.

In addition to future emergence of VOCs, it is impossible to predict future policy or behavioural changes with any certainty. It remains unclear whether mobility and behaviour will return to the same levels seen prior to the COVID-19 pandemic. We have considered one scenario where behaviour remains largely unchanged, and three scenarios in which mobility increases to pre-pandemic levels in 3 weeks, 3 months or 6 months, to capture this uncertainty. We do not consider scenarios where behaviour increases above the baseline levels recorded prior to the COVID-19 pandemic, which may increase the extent and/or change the timing of any future resurgences if such an extreme behavioural change were to occur. In addition to longer-term changes, we are not able to forecast sudden shorter-term behavioural changes that have been observed in the data, e.g. around Christmas holidays (**Fig. 3**), although our 3-week return to baseline mobility scenario is intended to explore the consequences of a sudden increase in mobility (**Fig. S3**). Behavioural changes can also occur in response to awareness of a growing or declining epidemic^34, 35^, which is something that we do not capture in any forward projections.

There are a number of other limitations in this work which are important to consider. The fitted model does not accurately capture the dynamics of PCR prevalence in England from March 2022 onwards, during Omicron BA.2’s dominance (**Figs. 2, S1A-B**). The modelled transmission adjustment during this time increases, temporarily, to an extremely high level (**Fig. 3**). This suggests that we may not have captured some properties of the Omicron BA.2 sublineage, e.g., if BA.2’s transmission advantage over BA.1 is larger than the assumed 50%, or if BA.2 has a shorter serial interval^36^ or longer infectious period. Additionally or alternatively, it may be that behaviours that are not captured within mobility data—such as mask wearing, physical distancing, and testing—have changed substantially in recent months following the removal of all legal restrictions in February 2022^5, 37^. Further, our modelled estimates of the proportion of the population in England who have some form of immunity against SARS-CoV-2 are high (**Fig. S8**), which may explain the poor fitting to PCR prevalence during the Omicron BA.2 era, particularly if we are overestimating the level of protection conferred by vaccines and/or underestimating the rate at which immune protection from SARS-CoV-2 infection and from COVID-19 vaccination is lost.

The extent to which immunity from a prior infection and/or from COVID-19 vaccination and boosting wanes is uncertain. Individuals may retain very long-term protection against severe outcomes, but have faster rates of waning against less severe outcomes such as mild or asymptomatic infection. In our basecase scenario, we model waning vaccine protection for individuals who do not receive a booster vaccination by moving them into a vaccinated and waned disease state where their vaccine protection against different SARS-CoV-2 outcomes is reduced to different extents (see assumptions in **Tables S2 and S5B**). Vaccinated and waned individuals and individuals who have recovered following a SARS-CoV-2 infection can then further wane back to a completely susceptible disease state (based on assumed rates of waning shown in **Table S4**). For our basecase scenario, we have parameterised these waning rates in relation to measured reductions in protection against severe outcomes, but our assumption that individuals return to being completely naive to infection may still be overly pessimistic. On the other hand, our basecase scenario assumes that individuals who receive booster vaccinations do not wane back to complete susceptibility and that the protection afforded by their vaccinations remains at least at second-dose assumed levels throughout the time periods considered here (see vaccine effectiveness assumptions in **Table S2**).

Over the long term, individuals in the model who receive their first booster vaccination will eventually reach the end of their second-dose duration, where the same probability of boosting remains; therefore, some individuals in the model will continually receive boosters and retain second-dose levels of protection, whilst some individuals are not boosted by their second or third decision point and move into lower levels of protection. Given our assumed second-dose duration (see **Table S5B**), and given that boosters were first administered in England in September 2021, the earliest that individuals in the model will reach their second booster decision point is April 2022. Given that the JCVI in England has recommended a spring 2022 COVID-19 booster vaccine for the most vulnerable individuals who received their first boosters in September and October 2021^38^, ahead of a wider autumn 2022 booster programme, we think that it is reasonable to assume (i.e. as we have done in our central basecase scenario) that the majority of individuals will continue to receive booster vaccines. However, the modelling scenarios with higher rates of waning (high and very high waning, see **Table S4**) can be considered as a sensitivity analysis looking at a situation where fewer people continue to be boosted.

Throughout this work we have also assumed identical levels of protection and rates of waning across age groups, and we do not differentiate across risk groups. It is likely that protection will differ from person to person and that more clinically vulnerable individuals may have lower levels of protection or faster reductions in immunological protection following a previous infection or a vaccination. Future work needs to consider differential waning over time whereby higher protection against severe outcomes can be maintained but protection against infection is reduced, as well as of more complex immune dynamics induced by the numerous possible orders and timings of infections, vaccinations, reinfections and booster vaccinations and heterogeneity in individual’s immune responses.

We have assumed that following an infection with Omicron, individuals have complete cross protection against reinfection with Omicron or any other pre-existing variant (see cross protection assumptions in **Table S3**) whilst they remain in the recovered disease state. If reinfections following early Omicron infections are occuring^39, 40^, particularly since the BA.2 Omicron sublineage has become dominant in England^36^ and other sublineages are emerging worldwide, we may see a slower reduction in SARS-CoV-2 transmission in England than our modelling projects, and the potential for higher levels of transmission throughout the time period under consideration.

Following the Omicron BA.1 and BA.2 waves of SARS-CoV-2 transmission, and an extensive vaccination and booster vaccination campaign, our work suggests that levels of immunity in England are sufficiently high to lead to a sustained reduction in transmission in the coming months, assuming no new variants of concern emerge. The extent to which SARS-CoV-2 transmission resurges later in 2022 depends upon a combination of factors, including behaviour, the rate that immunity wanes, and seasonality. It is clear that the extent to which immunity, both derived from natural infection and from COVID-19 vaccination, wanes will become extremely important in the medium to long term. It is crucial to improve our current understanding of the dynamics of SARS-CoV-2 immunity and to consider the consequences of this on community transmission in the context of varying levels of PHSMs and vaccination coverage, both in England and worldwide.

## Online Methods

### Epidemiological model

We use an age-structured and region-specific deterministic dynamic compartmental model of SARS-CoV-2 transmission (**Fig. 1**), building on previous work^22, 41–45^. Geographic structure is by NHS England region (of which there are seven in England) and age groups are divided into 5-year age bands from 0–4 to 70–74 years, with an additional age group comprising individuals aged 75 years and over. The model tracks three variants of SARS-CoV-2 which enables us to capture wild-type, Alpha B.1.1.7, Delta B.1.617.2, and Omicron B.1.1.529 variants separately. The second and third modelled variants track Alpha B.1.1.7 and Delta B.1.617.2 respectively; the first modelled variant initially describes wildtype and other variants circulating prior to the emergence of Alpha B.1.1.7, before switching to describe the Omicron B.1.1.529 variant on 22nd September 2021. The date of 22nd September 2021 is chosen to be sufficiently before the emergence of Omicron in England and well after wildtype and other variants circulating prior to Alpha B.1.1.7 have been competitively excluded. On this date, we update all of the variant-specific parameters and assumptions for the first modelled variant to represent the Omicron variant, and ensure that any individuals remaining in the recovered disease state for the first variant are moved into the recovered disease state for the third variant, Delta B.1.617.2. To capture the effect of the BA.2 Omicron sublineage taking over from the previously-dominant BA.1 Omicron sublineage^36, 39^, we use sequencing data from the Wellcome Sanger Institute^46^ to inform the proportion of modelled Omicron infections attributable to BA.2 over time, assuming that BA.2 confers a 50% increase in transmissibility relative to BA.1^36, 39, 47–50^. We assume the same levels of vaccine protection and rates of waning immunity for both Omicron BA.1 and BA.2 sublineages^51^.

When individuals have recovered following a SARS-CoV-2 infection with one variant, they move into a recovered disease compartment where (depending on parameter assumptions, see **Table S3**) we allow for re-infections and cross-infections with other variants to occur. We model COVID-19 vaccination with separate compartments for two vaccine products—for each of the viral vector and mRNA-based COVID-19 vaccines in use in England—and with each vaccine product having three compartments for three levels of protection (one dose, two doses, and waned from two doses). Initially, vaccinated individuals move from the susceptible compartment to a first-dose vaccinated compartment (Va1/Vb1 in **Figure 1**) 28-days following receipt of their vaccination.

Individuals remain in the first-dose vaccinated states for an assumed duration (see dVa1/dVb1 in **Table S5B**), before transitioning into the second-dose vaccinated compartments (Va2/Vb2 in **Figure 1**). The first-dose duration assumptions are based on measured delays between first and second doses in UKHSA vaccination data, separated into two periods (before and after the JCVI issued guidance on widening the dosing gap from 3 weeks to a maximum of 12 weeks), see **Table S5B** for details. Upon leaving the first-dose vaccine state, individuals transition into the second-dose vaccine state with increased levels of protection (**Table S2**). Another assumed distribution governs the duration that individuals remain in the second-dose state (see dVa2/dVb2 in **Table S5B**). The distribution governing the duration that individuals have second-dose levels of vaccine protection is chosen to match the time between the start of the COVID-19 vaccination rollout (8th December 2020) and the start of the COVID-19 booster dose rollout (September 2021) less the assumed average duration of first-dose protection.

When individuals reach the end of their second-dose duration, they either receive a booster vaccination (using an age-specific probability of receiving a booster vaccination, see **Table S5B**) and return to the start of the mRNA second-dose vaccine compartment (Vb2 in **Figure 1**), or do not receive a booster and transition into vaccine product-specific vaccinated and waned compartments with lower levels of protection (Va2w/Vb2w in **Figure 1**). We assume that all boosted individuals move into the mRNA two-dose vaccinated (Vb2 in **Figure 1**) compartment to reflect the fact that all booster vaccinations in England are either the Pfizer/BioNTech or Moderna mRNA vaccines, regardless of which vaccine product was received for the primary COVID-19 vaccination course, and evidence finding higher immunogenicity for individuals receiving Pfizer/BioNTech following Oxford-AstraZeneca, compared with individuals receiving both Oxford-AstraZeneca vaccine doses^26^. In addition to individuals who receive a booster vaccination remaining in the second-dose mRNA vaccine compartment, for first booster vaccinations administered in late 2021/early 2022, we increase the level of protection within the two doses compartment to account for an additional booster effect (see *Booster vaccinations*). The model also captures eventual full waning of immunity that has been derived from a previous infection and/or vaccination. A full description of assumptions related to vaccine protection against different outcomes, cross protection and waning immunity is provided in the Supplementary material in **Tables S2- S4** and **Tables S6- S7**.

Hospital admissions and occupancy data were provided by NHS England and deaths, immunisations and variant data were provided by the UK Health Security Agency (UKHSA). These data sources are unpublished and not public, but are closely aligned with healthcare, deaths and vaccinations data on the UK Government COVID-19 Dashboard^2^. Seroprevalence data were obtained from the UK Biobank^52^ and the REACT-2 study^53^, and seroprevalence and PCR positivity data were obtained from the the Office for National Statistics COVID-19 Infection Survey (ONS-CIS)^27, 54^.

The age-specific susceptibility to infection and age-specific probability of clinical symptoms for SARS-CoV-2 are adopted from a study using data from 6 countries^29^. This study found that susceptibility to SARS-CoV-2 infection for 0–19-year-olds was roughly half that for >20-year-olds, and that the probability of clinical symptoms also increased with age^29^. The age-specific probability of hospital admission, ICU admission, and death given infection are fitted to data from England, with the relative rates by age group based on data collected by a large meta-analysis of the COVID-19 infection fatality rate^55^ and based on data collected by ISARIC (the CO-CIN study) for England^22^, then adjusted over time to better match observed hospitalisations and deaths in England (see *Model fitting*). Each of these age-specific probabilities of severe outcomes is allowed to vary over the course of the epidemic in England and vary between pre-existing variants and Alpha B.1.1.7. For the third variant Delta B.1.617.2, we assume that the probability of severe outcomes is twice that of Alpha B.1.1.7, in line with estimates from Public Health Scotland and UKHSA^31^. For the fourth variant Omicron B.1.1.529 (and both modelled sublineages BA.1 and BA.2), we assume that the probability of severe outcomes is half that of Delta B.1.617.2, in line with estimates from the UK^36, 56, 57^. The model fitted adjustments to the infection fatality, hospital admission, hospital bed occupancy and ICU bed occupancy rates over time is shown in **Figure S1C.** A full description of fitted and assumed parameters is provided in **Tables S5A** and **S5B**.

The model uses age-specific contact rates as measured by the POLYMOD study^58^ in the UK as a baseline contact matrix representing pre-pandemic age-specific mixing rates. The POLYMOD study estimates age-specific mixing rates for contacts made at home, work, school, and “other” settings, such that the “baseline” contact rate between age group *i* and age group *j* is 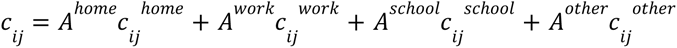, where *A^home^, A^work^, A^school^* and *A^other^* are coefficients on the four component matrices. For the pre-pandemic baseline matrix, then, *A^home^* = *A^work^*= *A^school^* = *A^other^* = 1 coefficients are allowed to vary over time according to mobility data, school schedules and school attendance data as follows. The model uses Google Community Mobility data^21^ to capture mobility in various settings: workplaces, retail & recreation venues, transit stations, and grocery & pharmacy locations. In turn, the relationship between mobility data and social contact rates^22^ is derived from the historical relationship between Google Community Mobility indices and social contact rates as measured by the CoMix study in 2020. This relationship furnishes how *A^home^, A^work^,* and *A^other^* vary over time as a function of mobility patterns.

School openings and closings are accounted for in contacts among school-aged children, university-aged young adults and school/university staff. We assume that schools in England follow their traditional schedules (i.e. are closed during holiday periods), and we combine these assumptions with school attendance data in England published on the 4th of May 2022^23^. To reflect the introduction of mass testing within educational facilities in the Spring of 2021, we have assumed an additional 30% reduction in transmission related to educational settings between the reopening of schools on 8th March 2021 and school closures in July 2021. This reduction in transmission is reflected in the model with a 30% reduction in school-related contacts. Seasonality is modelled as a sinusoidally-varying multiplier on transmission with the peak occurring on January 1st and the trough on July 1st of each year. By default, we assume the amplitude of the seasonal component is 20% from trough to peak and is introduced from 1st April 2021.

### Model fitting

The model is fitted using a two-stage process. In the first stage, the model parameters are fitted by Bayesian inference using Markov chain Monte Carlo (MCMC) to reported regional data on hospital admissions, hospital and ICU bed occupancy, seroprevalence, PCR positivity, and deaths within 28 days of a patient’s first positive SARS-CoV-2 test, as well as to data tracking the emergence and spread of the Alpha B.1.1.7 and Omicron B.1.1.529 variants (using the frequency of S-gene target failure in PCR tests) in late 2020 and late 2021 respectively, and of the Delta B.1.617.2 variant (using the frequency of Delta in genomic sequencing data) in 2021. We use the DE-MCMC algorithm^59^ implemented in C++ (see analysis code).

The introduction time and relative transmissibility of both the Alpha, Delta, and Omicron variants are fitted for each geographic region in the model. We use data recording the number of first COVID-19 vaccine doses delivered by age, geography and vaccine product from 8th December 2020 to 24th March 2022 to inform the fraction of first-dose vaccinated individuals in each age group, NHS England region and by vaccine type over time. Additionally, in this initial stage, 18 additional parameters are fitted to define a “transmission multiplier” function, with each of the 18 parameters defining a stepwise change in transmission occurring at fixed six-week intervals beginning 12th April 2020 and ending 8th May 2022. This transmission multiplier function allows the modelled epidemic trajectory to better capture changes in SARS-CoV-2 transmission over time, and reflects residual changes in transmission that are not captured by mobility data alone, e.g. as resulting from changes in personal protective behaviours such as mask-wearing or from changes in social behaviour such as during holidays.

In the second stage of model fitting, a particle filtering algorithm^60^ is used to refine the “rough” transmission multiplier function, while holding other fitted model parameters constant, to achieve a more fine-scaled function with stepwise changes to the transmission multiplier every 5 days, instead of the cruder 6-week increment used in the initial stage of fitting. In the second stage, the transmission multiplier is fitted as a random walk on a logarithmic scale, that is, multiplicative increments to the transmission multiplier are proposed rather than absolute levels.

The same likelihood is used for Bayesian inference in both stages of the model fitting process. In particular, the likelihood allows the infection fatality rate (IFR), infection hospitalisation rate (IHR), and infection critical-illness rate (ICR) to vary over the course of the epidemic, to reflect changes in treatment success, admissions criteria, and availability of hospital resources over time. To achieve this, the model’s initial output of deaths and hospital burdens, which are based upon a fixed IFR, IHR, and ICR (except as modified by variant-specific characteristics and by vaccine protection), are treated as the prior expectation for deaths and hospital burdens on each day. This expectation is used as the mean of a gamma distribution, with standard deviation set to 0.3 times the mean, which, in turn, is taken as the prior distribution for the mean of a Poisson distribution from which the observed burden for a given day is assumed to be drawn. This process allows the IFR/IHR/ICR to change over time, while not straying too far from the expected burden based upon the underlying fixed IFR/IHR/ICR.

### Model assumptions

#### Vaccine effectiveness

We model vaccine protection against five separate outcomes for each SARS-CoV-2 variant: infection, disease (i.e. symptomatic infection), hospitalisation, mortality and onward transmission following a breakthrough infection (i.e. when an individual who has vaccine protection becomes infected). We assume the same vaccine effectiveness for the first two SARS-CoV-2 variants considered in the model (pre-Alpha B.1.1.7 and Alpha B.1.1.7), and separate specific vaccine effectiveness estimates for the Delta B.1.617.2 and Omicron B.1.1.529 variants, shown in **Table S2**. Throughout, we assume identical vaccine effectiveness across all age groups in the model. These may be subject to change in future work, as new evidence emerges. We treat individuals who have been and will be vaccinated with Moderna vaccines the same as individuals receiving Pfizer vaccines. We model individuals who have received different vaccine products (e.g. viral vector AstraZeneca (Va compartments in **Figure 1**) and mRNA Pfizer/Moderna (Vb compartments in **Figure 1**)) and one or two vaccine doses separately, assuming separate efficacy estimates for each category (**Table S2**). We additionally consider individuals who have received two vaccine doses but no booster dose as having reduced levels of protection in the vaccinated and waned state (**Figure 1, Table S2**).

We base our vaccine effectiveness assumptions for the pre-Alpha, Alpha and Delta variants on a number of studies considering vaccine efficacy and effectiveness (**Tables S2, S6, S7**). For the Omicron B.1.1.529 variant (comprising both BA.1 and BA.2 sublineages), we assume a 5.5-fold reduction in neutralisation between Delta and Omicron^44^, resulting in approximately 45% lower vaccine protection against infection. We use the resulting vaccine protection against infection with Omicron to generate assumptions for vaccine protection against Omicron disease, hospitalisation and mortality. For protection against disease, we assume the same conditional protection against disease given infection as for the Delta B.1.617.2 variant. For protection against hospitalisation and mortality, we use Khoury et al.’s modelled relationship between efficacy against any infection and efficacy against severe infection^61^ to scale up our assumptions for protection against infection with Omicron to higher levels of protection against these two severe outcomes. For protection against onward transmission, we use the same assumptions as for the Delta B.1.617.2 variant (**Table S2**).

Our assumptions about the levels of vaccine protection for individuals who have received two vaccine doses but no booster dose, and their protection has waned, are shown in **Table S2**. We have based our assumptions for vaccine protection against infection, disease and onward transmission on scaling down assumed levels of protection for dose-two vaccinated individuals using vaccine product-specific measured percentage reductions in vaccine protection over time from Andrews et al.^19^. For protection against infection and disease, we referred to the measured product-specific percentage reductions in protection against symptomatic infection for individuals aged 16 years and older after 20+ weeks, compared to measured protection in week 1. For reductions in protection against onward transmission, we calculate the average product-specific measured percentage reduction in protection across the measured percentage changes in protection against symptomatic infection and hospitalisation measured at 20+ weeks compared to week 1 and the percentage change in protection against death measured at 20+ weeks compared to weeks 2-9. Since we model an individual’s average duration in the second dose compartment as 29.3 weeks (**Table S5B**), before they are either boosted and remain in the two-dose vaccinated compartment or are not boosted and wane, we have chosen to further reduce our scaled vaccine effectiveness estimates against infection and onward transmission resulting from using Andrews et al.^19^ 20+ week estimates by an additional 25% reduction on top of the measured percentage reductions (**Table S2**). To balance this assumption, we have not included an additional 25% reduction in protection for the assumed level of vaccine protection against disease for vaccinated and waned individuals (**Table S2**). To arrive at assumptions for vaccine protection against severe outcomes (hospitalisation and mortality) for individuals in the vaccinated and waned state, we use Khoury et al.’s modelled relationship between efficacy against any infection and efficacy against severe infection^61^ to increase our assumed levels of protection against infection up to higher levels (**Table S2**).

#### Booster vaccinations

We model the effect of booster vaccinations by boosting individuals in two categories: those who remain in the second-dose vaccinated compartment following receiving a booster as well as individuals who are in the recovered disease state but would also have received a primary course of COVID-19 vaccination followed by a booster vaccination. Boosted individuals remain in their current disease state, but we model an additional increase in their level of vaccine protection and cross protection against reinfection following their first booster vaccine in late 2021/early 2022 as follows. We firstly assume that the Pfizer-BioNTech and Moderna booster vaccinations being administered in England result in a 2.5-fold increase in neutralisation titres^62^, and we use Khoury et al.’s modelled relationship between neutralisation titres and protective efficacy^61^ to increase our two-dose vaccine effectiveness and cross protection assumptions against infection to boosted levels (boosted vaccine effectiveness assumptions are shown in **Table S2**). For the boosted levels of cross-protection against infection in the recovered disease state, we use the same increase in protection but this is applied only to the proportion of recovered individuals who would have received a primary vaccination course multiplied by the age-specific probability of receiving a booster vaccine (see **Table S5B**). We scale up the boosted protection against infection to equivalent levels of protection against hospitalisation and mortality using Khoury et al.’s modelled relationship between efficacy against any infection and efficacy against severe infection^61^.

We refer to data on the number of COVID-19 booster vaccinations being delivered over time on the UK Government’s COVID-19 dashboard^2^ to inform the time at which the additional booster effect is applied to each 5-year age group, from individuals aged 75 years and above to individuals aged 15 years and above, in sequentially younger age groups over time. Between the start of the COVID-19 booster vaccination rollout (September 2021) and the 15th of December 2021, we assume a daily supply of booster vaccines of 229,000. We assume that booster supply increases to 1 million doses per day from the 15th of December 2021, following the JCVI announcement of an acceleration and widening of the booster vaccination programme^14^. Using these daily supply levels allows us to approximate the effects of the booster vaccination campaign by introducing the assumed booster effect on vaccine protection and cross protection at specific times for each 5-year age group in the model, from the oldest to the youngest eligible group. We assume that this additional booster effect lasts for 180 days, before vaccine protection returns to assumed two-dose levels (**Table S2**) and cross protection returns to previous levels (**Table S3**).

#### Waning immunity

We model waning immune protection against SARS-CoV-2 developed from a prior SARS-CoV-2 infection and/or vaccination. For waning immune protection following a previous infection, we assume identical rates of waning for all virus variants and for all age groups (**Table S4**). Once individuals who have recovered from a prior infection wane, they return to a susceptible disease state; thus we model waning of so-called natural immunity against different endpoints (infection, disease, hospitalisation, deaths and onward transmission) at the same rate. For our central waning assumptions, we do not allow individuals in the two-dose vaccinated compartments to wane directly back to being susceptible. To account for booster vaccinations, upon reaching the end of the assumed duration within the second dose state (**Table S5B**), individuals either receive a booster vaccine and return to the start of the second-dose compartment (with additional levels of protection afforded for the first booster vaccination in late 2021/early 2022, given an assumed booster duration of 180 days, see description above), or move into a third state with reduced levels of vaccine protection across different outcomes (see **Table S2**). This third state corresponds to individuals who have received two vaccine doses and no booster dose, leading to waning of their vaccine protection (see *Vaccine effectiveness* above). Once individuals have moved into this waned state with reduced levels of vaccine protection, they are also allowed to wane back to being susceptible, with different rates considered for each vaccine product (**Table S4**). The assumed percentage loss in reduction for the central waning scenario is based on measured percentage changes in vaccine protection against hospitalisation after 20+ weeks for each vaccine product in Andrews et al.^19^

### Mobility and vaccination schedules

To produce forward projections, the model requires information about future contact rates and vaccination rates. We base our assumptions on how social contact rates might be expected to change by referring to historical mobility data^21^ and making assumptions about future mobility until December 2022 (**Fig. 3**). Mobility levels have generally been gradually increasing since March 2022 but have still not returned to pre-pandemic levels. Hence we consider four scenarios: we project current levels of mobility forwards (i.e. no change), and we consider a return to pre-pandemic baseline levels of mobility within periods of 3 weeks, 3 months and 6 months. These assumed future changes in mobility are combined with future school term schedules to generate an overall schedule for future contact rates (**Fig. 3**).

The time-varying transmission multiplier also needs to be projected forwards for model projections. This is done by using the R package *forecast*^63^ to fit a second-order moving-average model to the fitted transmission multiplier function, then using random forecasts of the resulting moving-average model for each new projection. This process of using different realisations of the projected transmission multiplier is what produces most of the variability among different runs of each scenario (**Fig. 4c**). For forward projections, we combine the assumed future contact rates schedule with the projected transmission multiplier (**Fig. 3**) to produce an “overall” time-varying transmission potential until the end of the simulation period.

For future first-dose vaccinations, we generate vaccine schedules according to assumed future vaccine supply (i.e. number of doses available per week) and uptake limits per 5-year age group, in combination with the historic first-dose vaccine schedules generated using PHE/UKHSA data on vaccines delivered up to the 2nd of May 2022. Future first-dose vaccine supply is assumed to be 150,000 doses per week for England throughout the remainder of the projection period (i.e. until December 2022). We assume that first-dose vaccination uptake limits in individuals aged 15 years and above have already been reached, so no future first doses are delivered to these age groups. Our basecase scenario assumes that uptake is limited at 80% for individuals aged 5-14, delivering future first doses first to the 10-14 age group up to the uptake limit, followed by 5-9 year olds. This assumption of 80% uptake for 5-14 year olds is chosen based on the first-dose uptake measured by age across England; uptake is lower for younger age groups, with uptake reaching at least 80% as of May 2022 for the youngest eligible adults, who have been eligible to receive their first COVID-19 vaccinations in England since the summer of 2021. We also consider the impact of an alternative vaccination schedule generated assuming lower uptake levels for children aged 5-14 years (**Fig. S7**).

The assumed future supply of first doses per week is firstly divided into daily supply levels. Supply levels for each day are distributed into the seven NHS England regions according to the population size of each region. The allocated number of future first doses per day in each region are initially allocated to the oldest age groups which have not yet reached their age-specific uptake limits (i.e. since we assume maximum uptake has been reached for individuals aged 15 and above, 10-14 year olds are prioritised first up to their 80% uptake limit, followed by 5-9 year olds up to their uptake limit). The allocated number of first doses per day, per region and per age group are divided into specified proportions of vaccine products relevant to each age group (75% Pfizer and 25% Moderna for <40 year olds, and 60% AstraZeneca, 30% Pfizer and 10% Moderna for 40+ year olds; since May 2021 the JCVI has advised a preference for individuals under 40 years of age to receive an alternative to the viral-vector Oxford-AstraZeneca vaccine^64^). Any doses remaining after this process are carried over to the next age group down (up to the relevant uptake limit and reallocated according to our assumptions on vaccine product mix), the next NHS England region, or the next day, or are left unallocated in the schedule and recorded as leftover doses.

## Data Availability

Analysis code and data will be made available upon publication at: https://github.com/rosannaclairebarnard/newcovid3

## Contributions

RCB and NGD accessed and verified the data and conducted the analyses. All authors contributed to study design and drafting of the manuscript.

## Declaration of interests

RCB, NGD, MJ and WJE are participants of the UK’s Scientific Pandemic Influenza Group on Modelling. WJE attends the UK’s Scientific Advisory Group for Emergencies. All authors declare no competing interests.

See Supplementary material for working group authors and acknowledgments.

## Statement on data availability

Data sources used for these analyses are either publicly available, or were provided to the named authors as members of the UK’s Scientific Pandemic Influenza Modelling (SPI-M) group, which provides expert advice to the UK’s Department of Health and Social Care and wider UK government on scientific matters. Specific permissions have been sought to use SPI-M data for this publication. The SPI-M datasets used for model fitting are unpublished and not publicly available, but are closely aligned with the UK Government’s COVID-19 dashboard^2^ and other publicly available sources such as the Wellcome Sanger Institute’s COVID-19 genomic surveillance data^65^. Where publicly available datasets were used in our analysis, we have included references indicating where the data can be accessed.

## Statement on code availability

Analysis and code will be made available upon publication at: https://github.com/rosannaclairebarnard/newcovid3

## Funding statement

The following funding sources are acknowledged as providing funding for the named authors. This project has received funding from the European Union’s Horizon 2020 research and innovation programme - project EpiPose (Grant agreement number 101003688: RCB, MJ, WJE). This work reflects only the authors’ view. The European Commission is not responsible for any use that may be made of the information it contains.

This project has also received funding from the UK Medical Research Council (MC_PC_19065: NGD, WJE), was partly funded by the National Institute for Health Research (NIHR) (Health Protection Research Unit for Immunisation NIHR200929: NGD, MJ; Health Protection Research Unit in Modelling and Health Economics NIHR200908: MJ, WJE; PR-OD-1017-20002: WJE). Funding statements for CMMID COVID-19 working group members are listed in the Supplementary material.

## Ethics

Ethical approval for this research was given by the London School of Hygiene & Tropical Medicine Ethics Committee, project ID: 22828.

## Supplementary material

**Figure S1A.**
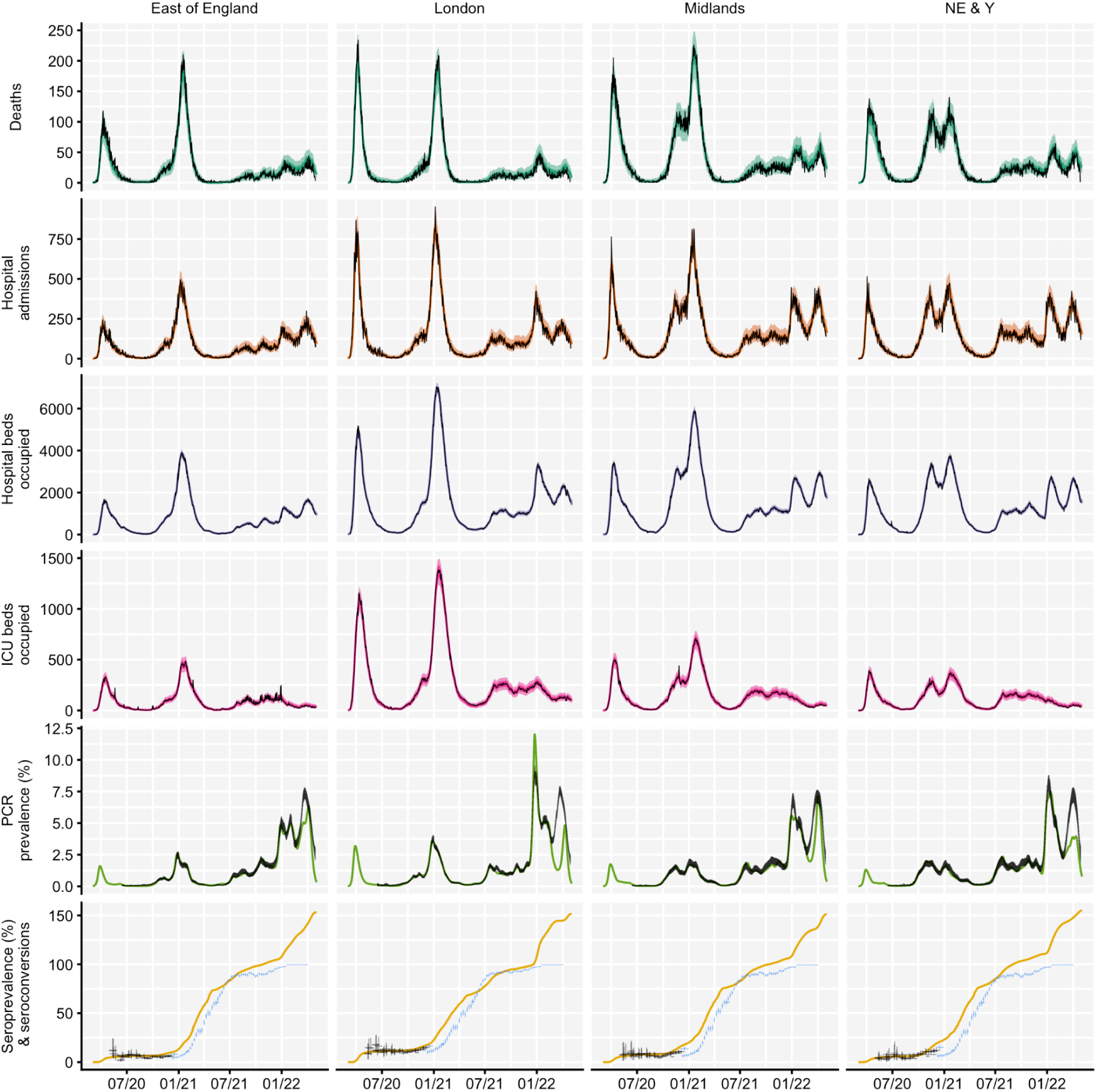
Regional model fit using epidemiological data from NHS England regions between March 2020 and May 2022. Four out of seven NHS England regions are shown here: East of England, London, Midlands and North East & Yorkshire (NE & Y); see Figure S1B for the remaining three NHS England regions. Black lines show reported data, with vertical black lines showing 95% confidence intervals for PCR prevalence and seroprevalence estimates. Seroprevalence estimates from 1st December 2020 onwards are not used for model fitting and are plotted in blue on top of the modelled cumulative number of seroconversions over time. Coloured lines and shaded areas show medians, 50% and 90% interquantile ranges from the fitted model. COVID-19 deaths data was provided by the UK Health Security Agency (UKHSA) and hospital admissions, hospital and ICU bed occupancy data was provided by NHS England. These data sources are unpublished and not publicly available, but are closely aligned with the UK Government’s COVID-19 dashboard^2^. PCR prevalence data was obtained from the Office for National Statistics’ COVID-19 Infection Survey (ONS-CIS)^66^. Seroprevalence data was obtained from the UK Biobank^52^, REACT-2 study^53^ and from the ONS-CIS^54, 66^. ICU = intensive care unit. NHS = National Health Service.

**Figure S1B.**
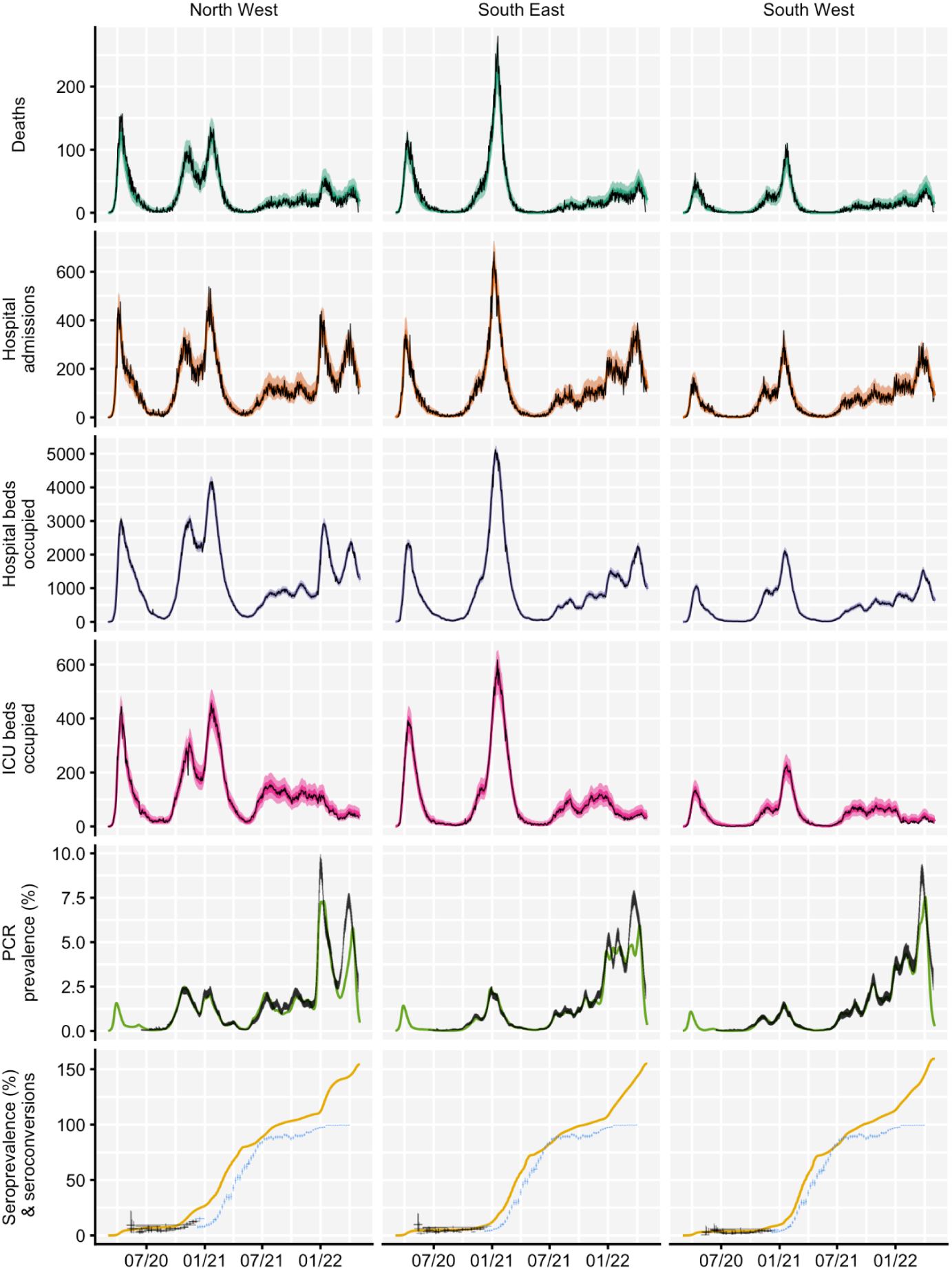
Regional model fit using epidemiological data from NHS England regions between March 2020 and May 2022. Three out of seven NHS England regions are shown here: North West, South East and South West; see Figure S1A for the remaining four NHS England regions. Black lines show reported data, with vertical black lines showing 95% confidence intervals for PCR prevalence and seroprevalence estimates. Seroprevalence estimates from 1st December 2020 onwards are not used for model fitting and are plotted in blue on top of the modelled cumulative number of seroconversions over time. Coloured lines and shaded areas show medians, 50% and 90% interquantile ranges from the fitted model. COVID-19 deaths data was provided by the UK Health Security Agency (UKHSA) and hospital admissions, hospital and ICU bed occupancy data was provided by NHS England. These data sources are unpublished and not publicly available, but are closely aligned with the UK Government’s COVID-19 dashboard^2^. PCR prevalence data was obtained from the Office for National Statistics’ COVID-19 Infection Survey (ONS-CIS)^27^. Seroprevalence data was obtained from the UK Biobank^52^, REACT-2 study^53^ and from the ONS-CIS^54, 66^,. ICU = intensive care unit. NHS = National Health Service.

**Figure S1C.**
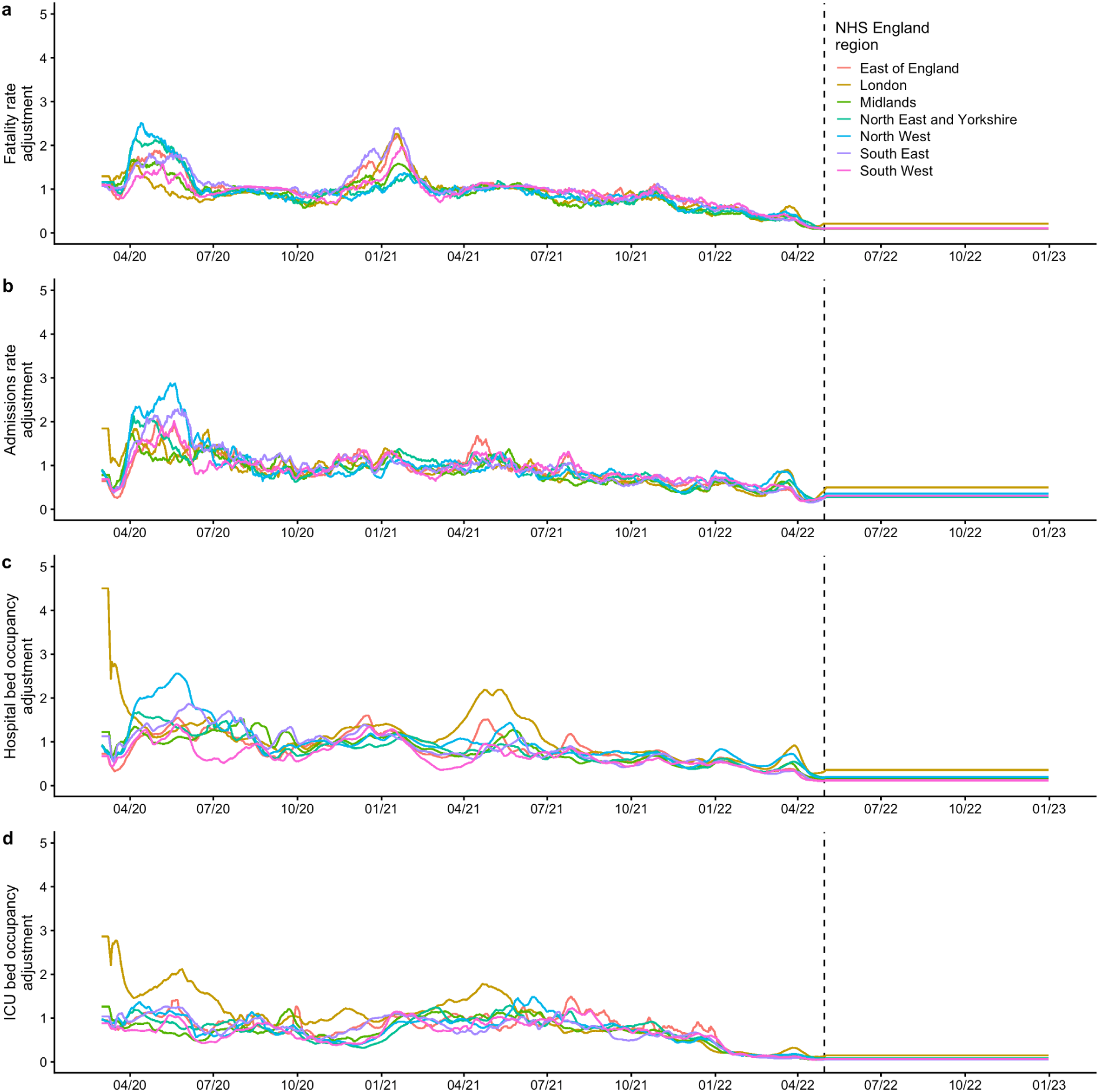
Modelled time-varying adjustments to rates of severe COVID-19 outcomes for the basecase scenario between March 2020 and December 2022. Vertical dashed lines show the final date of data used for model fitting, after which the final measured adjustment value is carried forward for the remainder of each simulation. **(a)** Modelled adjustments to the infection fatality rate over time, plotted for all seven NHS England regions. **(b)** Modelled adjustments to the hospital admission rate over time. **(c)** Modelled adjustments to the hospital bed occupancy rate over time. **(d)** Modelled adjustments to the intensive care unit (ICU) bed occupancy rate over time. NHS = National Health Service.

**Figure S2.**
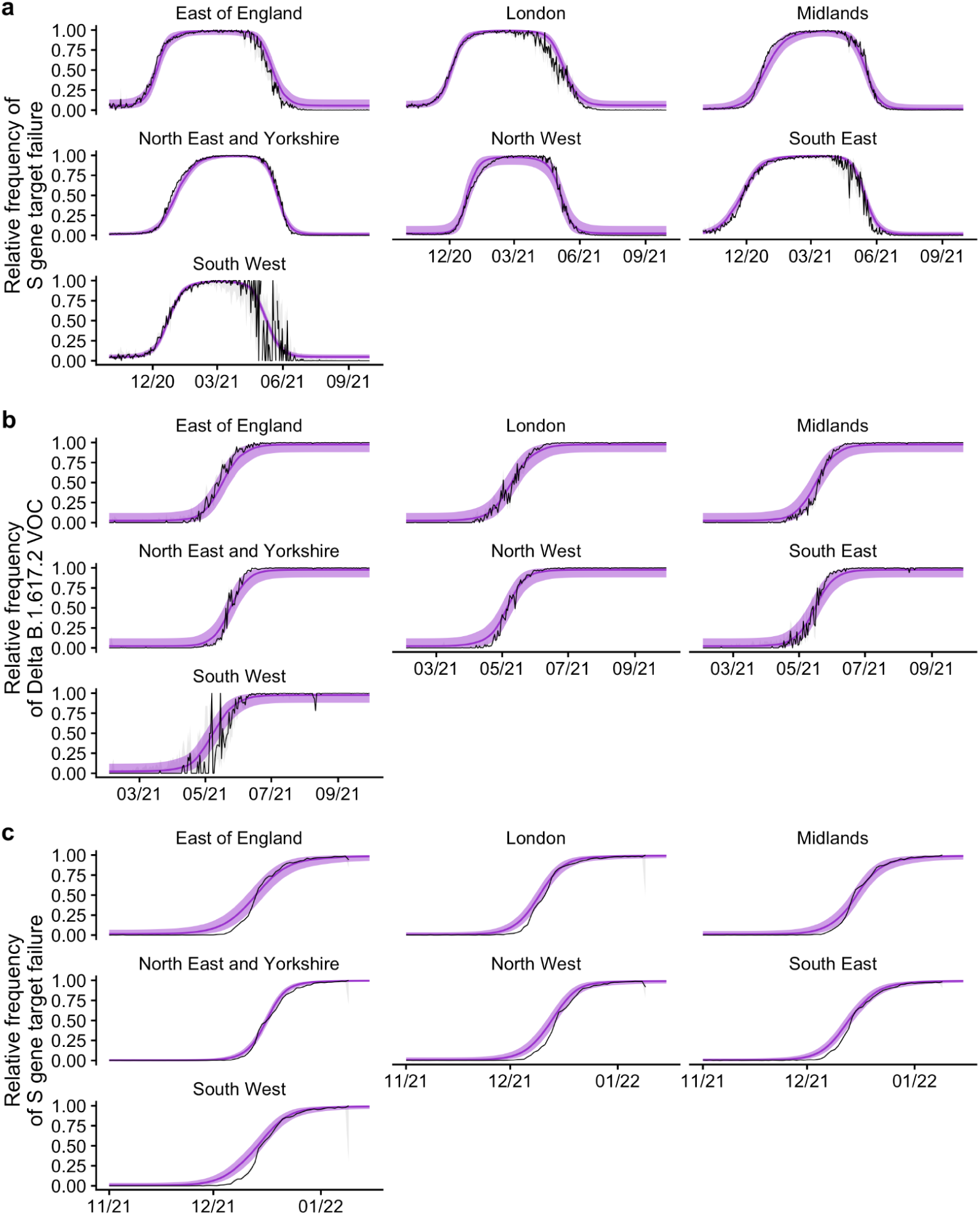
Regional model fits to the B.1.1.7 Alpha, B.1.617.2 Delta and B.1.1.529 Omicron variants of concern (VOCs) using S-gene target failure data and genomic sequencing data from NHS England regions between October 2020 and January 2022. **(a)** For the Alpha B.1.1.7 VOC, we use the frequency of S-gene target failure between October 2020 and September 2021 as a proxy for the proportion of infections attributable to Alpha over time. **(b)** For the Delta B.1.617.2 VOC, we use genomic sequencing data measuring the proportion of sequenced Pillar 2 cases attributable to Delta between February and September 2021. **(c)** For the Omicron B.1.1.529 VOC, we use the frequency of S-gene target failure between November 2021 and January 2022 as a proxy for the proportion of infections attributable to Omicron over time. In all three panels, black lines show reported data, with grey shaded regions showing 95% confidence intervals for the relative frequency of S-gene target failure in Pillar 2 PCR confirmed cases of SARS-CoV-2 (panels **a** and **c**) and the relative frequency of the Delta B.1.617.2 VOC in sequenced Pillar 2 PCR confirmed cases (panel **b**). Coloured lines and shaded areas show medians and 95% interquantile ranges from the fitted model. S-gene target failure and sequencing data was provided by the UK Health Security Agency (UKHSA). These data sources are unpublished and not publicly available. NHS = National Health Service.

**Figure S3.**
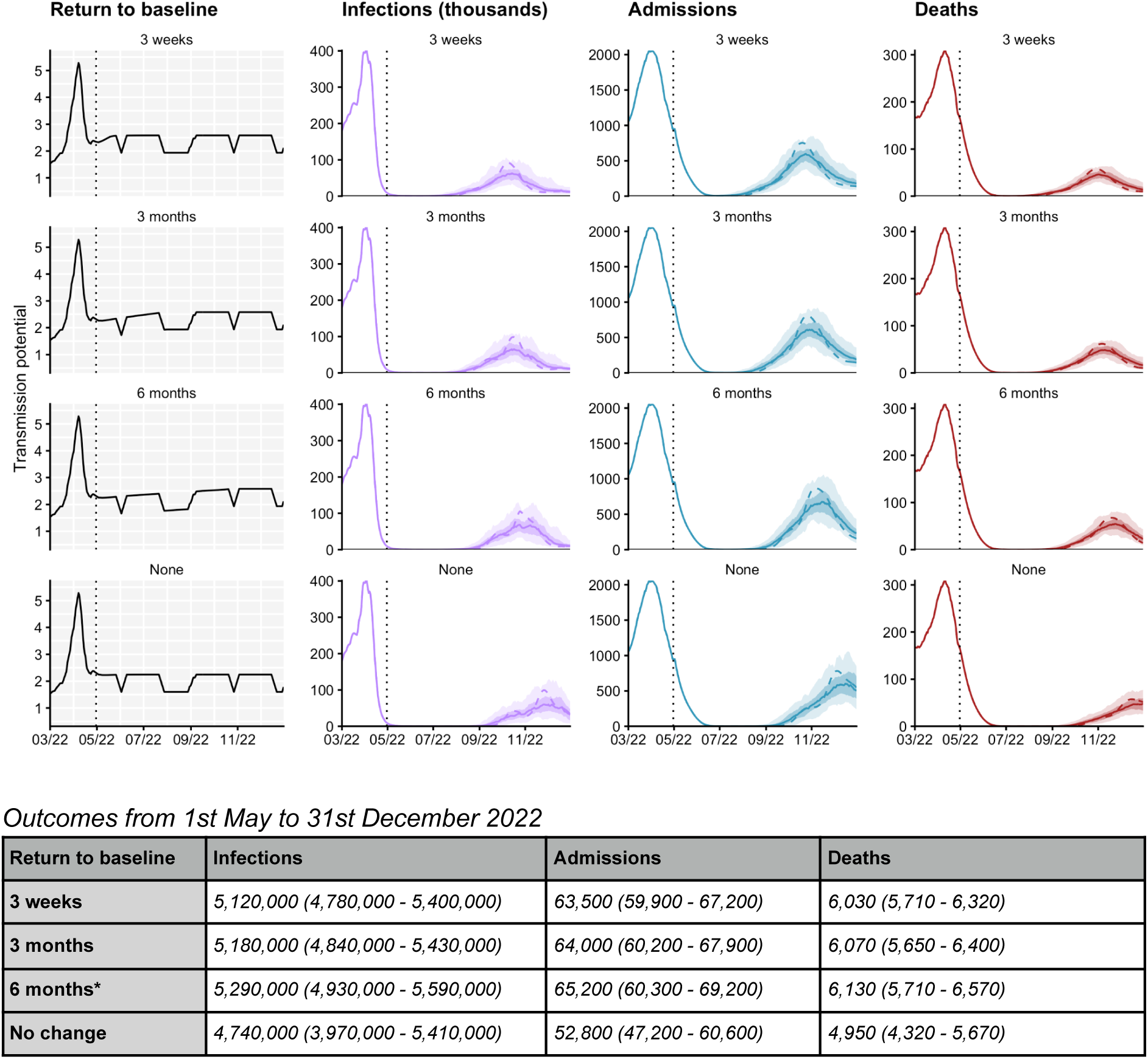
Impact of behaviour change on projected dynamics of SARS-CoV-2 transmission in England from March to December 2022. Top left: Overall transmission potential over time, incorporating mobility data, transmission adjustments and school term dates. Top right: Possible trajectories for COVID-19 infections (thousands), hospital admissions, and deaths are simulated for different rates of return to pre-pandemic baseline levels. The shaded areas and solid lines show the 50% and 90% interquantile ranges and the median for each time point, while the dashed line shows one sample trajectory. The vertical dotted lines denote the end of model fitting and the beginning of model projections. All scenarios assume central waning of vaccine protection (see Table S4), measured booster vaccination uptake relative to second dose uptake as of April 2022^24^, and 20% seasonality introduced from 1st April 2021. Full details about the assumptions for each scenario are given in Table 1. Table: the total number of COVID-19 infections, hospital admissions, and deaths, between 1st May and 31st December 2022, shown to 3 significant figures. The 6-month scenario is marked with an asterisk (*) and corresponds to the basecase scenario.

**Figure S4A.**
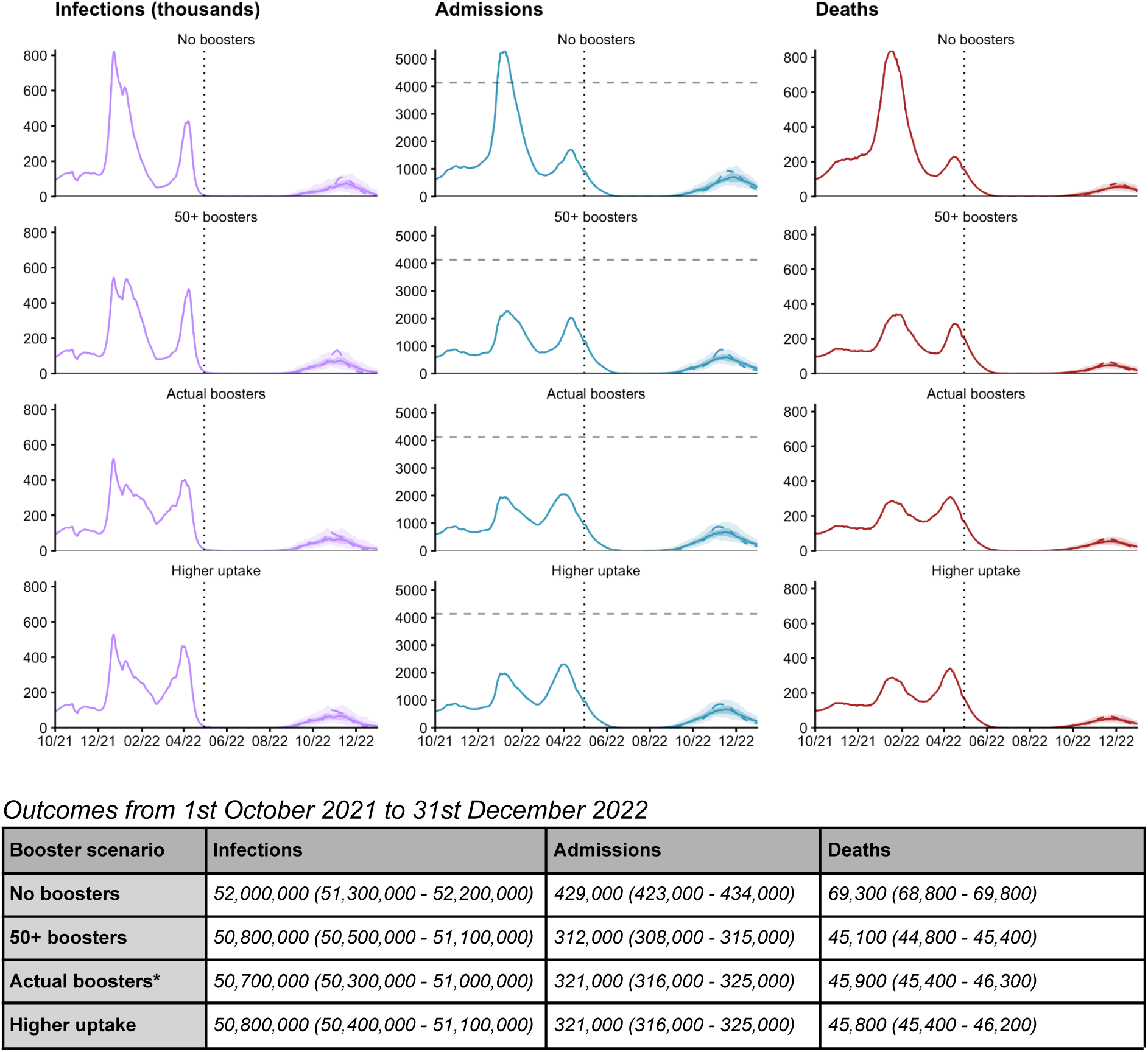
Impact of booster vaccination uptake on projected dynamics of SARS-CoV-2 transmission in England from October 2021 to December 2022. Top: Possible trajectories for COVID-19 infections (thousands), hospital admissions and deaths are simulated until December 2022, with different assumptions used for COVID-19 booster vaccination uptake. From top to bottom, we consider: a counterfactual scenario where booster vaccines were not available, a scenario where booster vaccinations were only offered to individuals aged 50 and above (at 95% uptake), the basecase scenario which is matched to measured uptake of booster vaccines relative to second dose uptake as of April 2022, and an additional counterfactual scenario where higher booster vaccination uptake was achieved (90% for individuals aged 15-49 and 98% for individuals aged 50 and over). Shaded areas and solid lines show the 50% and 90% interquantile ranges and the median for each time point, while the dashed line shows a single sample trajectory. The vertical dotted lines denote the end of model fitting and the beginning of model projections. The horizontal dashed line denotes the maximum number of daily recorded COVID-19 hospitalisations in England^2^. Full details about the assumptions for each scenario are given in Table 1. Tables: the total number of COVID-19 infections, hospital admissions, and deaths, between 1st October 2021 and 31st December 2022 and between January and December 2022, shown to 3 significant figures. The actual boosters scenario is marked with an asterisk (*) and corresponds to the basecase scenario. **Figure S4B. Impact of booster vaccination scenarios on modelled proportions of the population in England in different disease states from October 2021 to December 2022.** The modelled proportion of individuals across all ages in England in different disease states (from top to bottom: susceptible, infected, immune (natural protection), and vaccinated with 2-dose levels of protection) between October 2021 and December 2022, shown for one model run from each scenario. Booster vaccination rollout in England started in September 2021, initially targeted to at-risk individuals and individuals aged 50 years and above, 6 months after their previous COVID-19 vaccination. In December 2021 following increasing numbers of Omicron cases, the booster vaccination rollout was accelerated and extended to all individuals aged 18 years and above, with the minimum recommended gap between the previous vaccination and the booster dose shortened to 3 months^14^. Later in December 2022, booster vaccinations were also recommended for individuals aged 16 and 17 years old, at least 3 months following completion of their primary COVID-19 vaccination course^16^. The counterfactual scenario without any booster vaccination uptake (red solid line) is projected to have resulted in a much larger wave of infections (mostly as a result of breakthrough infections in vaccinated individuals) in late 2021 to early 2022.

**Figure S4B.**
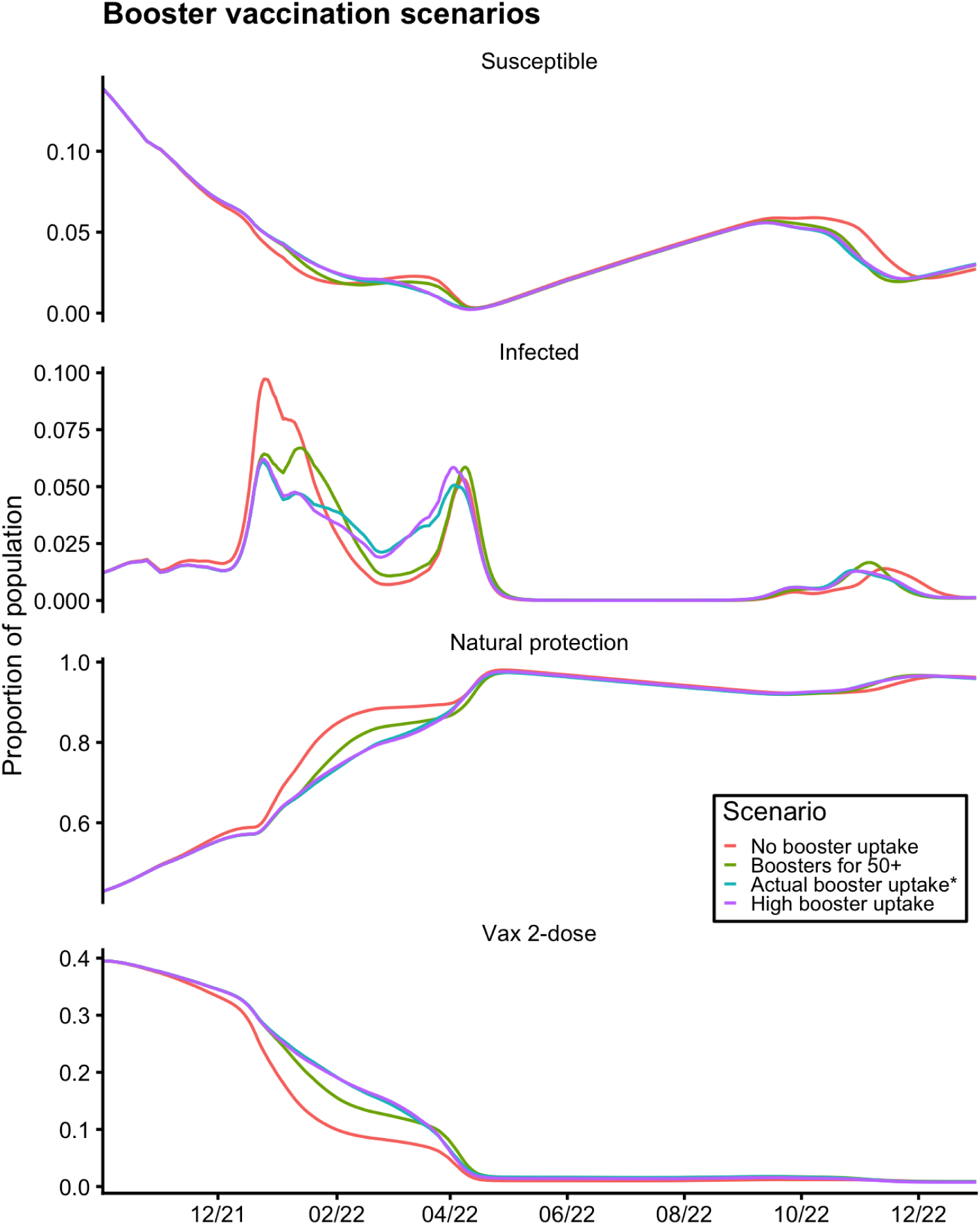
Impact of booster vaccination scenarios on modelled proportions of the population in England in different disease states from October 2021 to December 2022. The modelled proportion of individuals across all ages in England in different disease states (from top to bottom: susceptible, infected, immune (natural protection), and vaccinated with 2-dose levels of protection) between October 2021 and December 2022, shown for one model run from each scenario. Booster vaccination rollout in England started in September 2021, initially targeted to at-risk individuals and individuals aged 50 years and above, 6 months after their previous COVID-19 vaccination. In December 2021 following increasing numbers of Omicron cases, the booster vaccination rollout was accelerated and extended to all individuals aged 18 years and above, with the minimum recommended gap between the previous vaccination and the booster dose shortened to 3 months^14^. Later in December 2022, booster vaccinations were also recommended for individuals aged 16 and 17 years old, at least 3 months following completion of their primary COVID-19 vaccination course^16^. The counterfactual scenario without any booster vaccination uptake (red solid line) is projected to have resulted in a much larger wave of infections (mostly as a result of breakthrough infections in vaccinated individuals) in late 2021 to early 2022.

**Figure S5.**
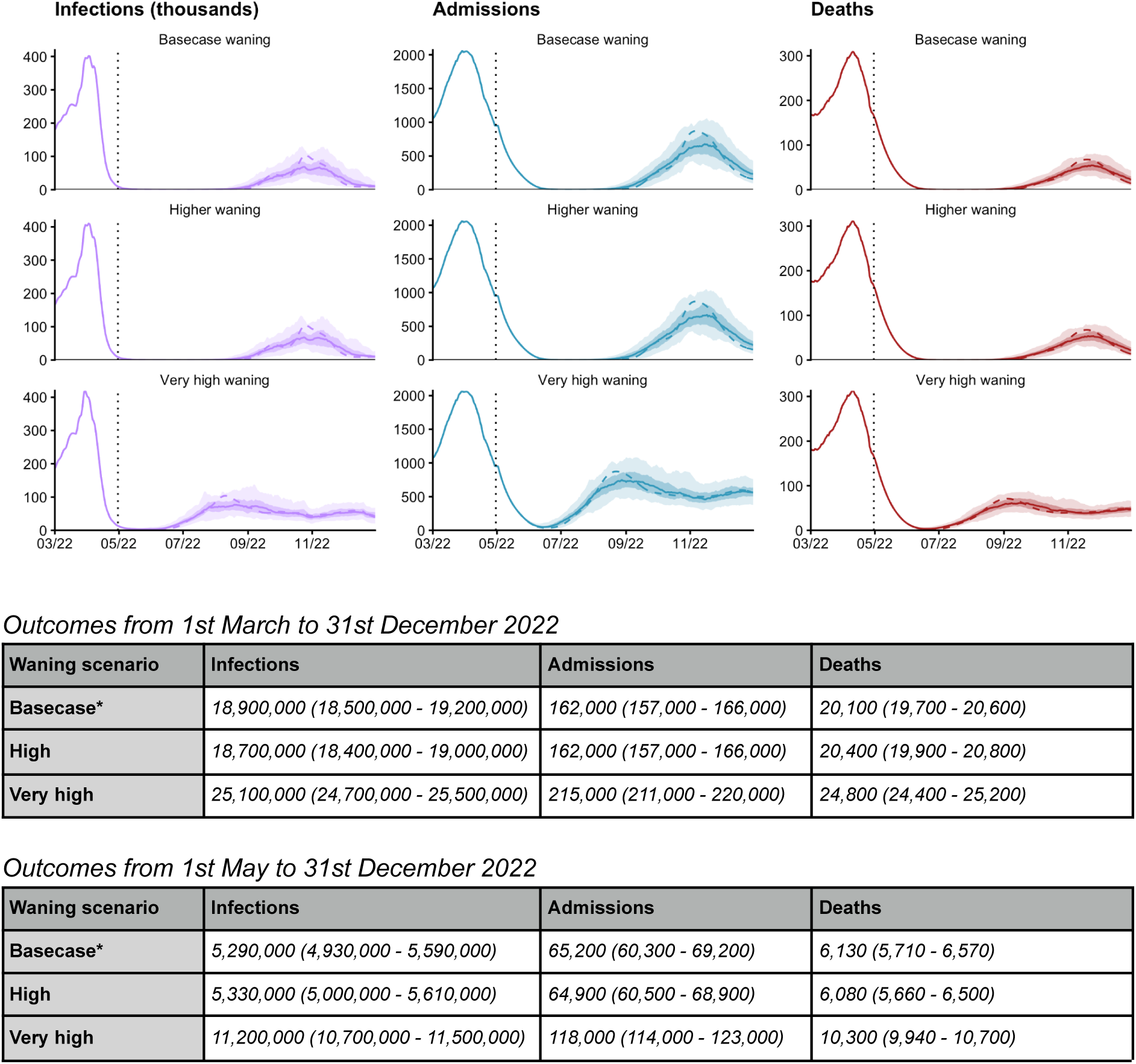
Impact of waning immunity on projected dynamics of SARS-CoV-2 transmission in England from March to December 2022. Top: Possible trajectories for COVID-19 infections (thousands), hospital admissions and deaths are simulated until December 2022, with different assumptions used for the rate that immunity (conferred from vaccination and following a prior infection) wanes (see Table S4). The shaded areas and solid lines show the 50% and 90% interquantile ranges, and the median for each time point, while the dashed line shows a single sample trajectory. The vertical dotted lines denote the end of model fitting and the beginning of model projections. Full details about the assumptions for each scenario are given in Table 1. Tables: the total number of COVID-19 infections, hospital admissions, and deaths, between 1st May and 31st December 2022, and between 1st January and 31st December 2022, shown to 3 significant figures.

**Figure S6.**
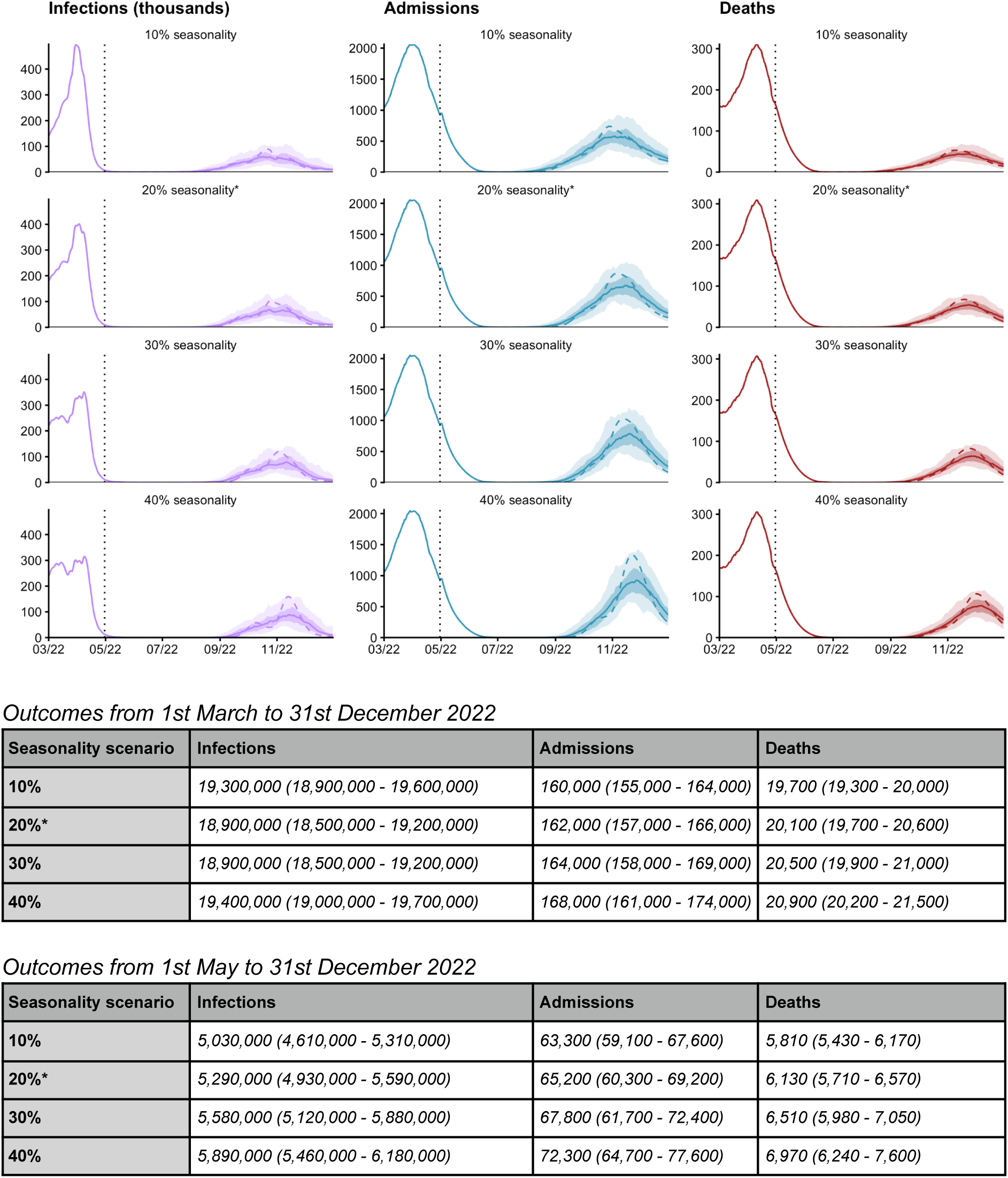
Impact of seasonality on projected dynamics of SARS-CoV-2 transmission in England from March to December 2022. Top: Possible trajectories for COVID-19 infections (thousands), hospital admissions and deaths are simulated until December 2022, with different assumptions made for the extent of seasonality in transmission. From top to bottom: 10%, 20%, 30% and 40% seasonality is introduced from 1st April 2021. The shaded areas and solid lines show the 50% and 90% interquantile ranges, and the median for each time point, while the dashed line shows a single sample trajectory. The vertical dotted lines denote the end of model fitting and the beginning of model projections. Full details about the assumptions for each scenario are given in Table 1. Tables: the total number of COVID-19 infections, hospital admissions, and deaths, between 1st March and 31st December 2022 and between 1st May and 31st December 2022, shown to 3 significant figures.

**Figure S7.**
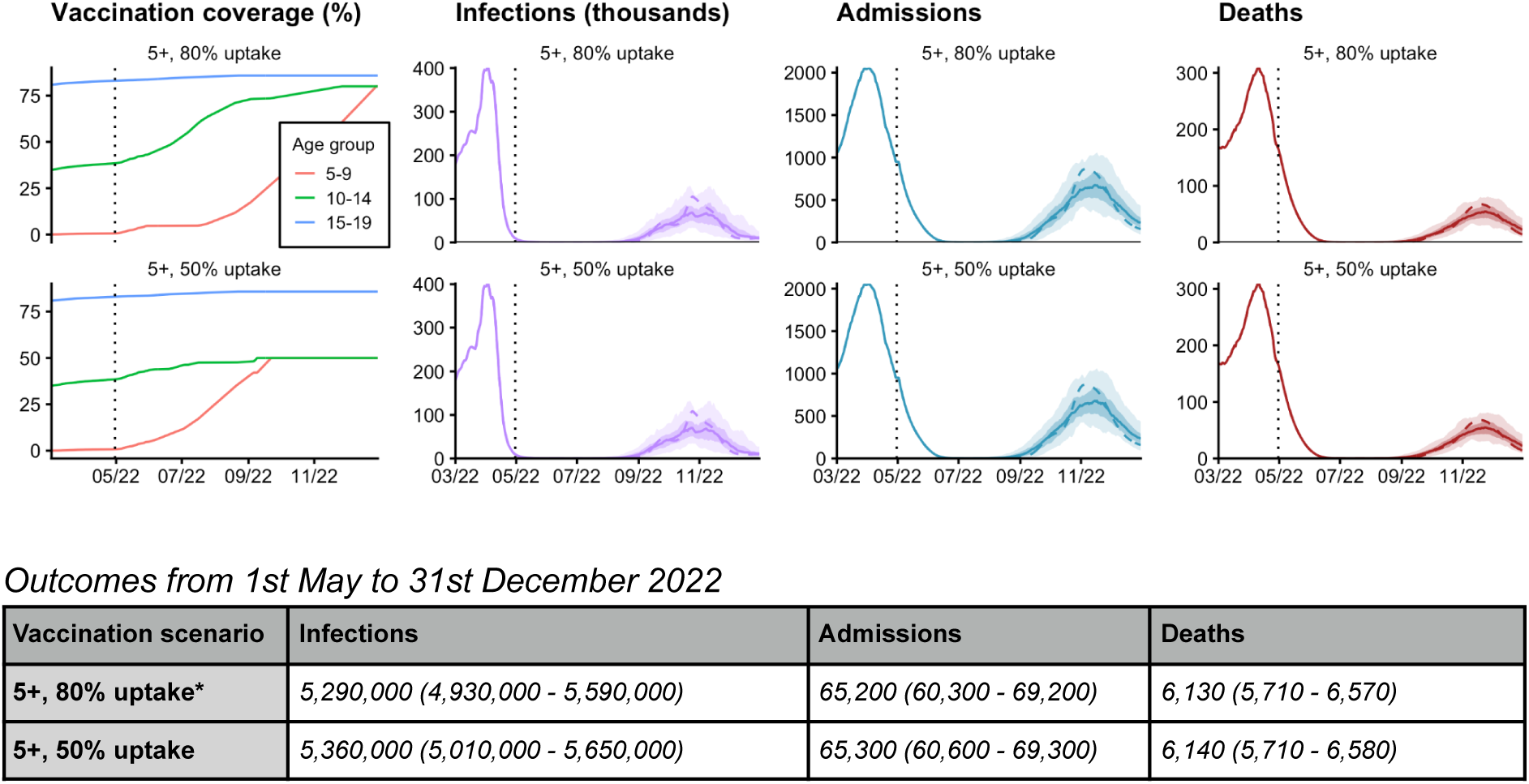
Impact of vaccinating adolescents and children on projected dynamics of SARS-CoV-2 transmission in England from March to December 2022. Top left: Vaccination coverage by age group over time for the two scenarios considered, shown for age groups 5-9, 10-14 and 15-19 only. From top to bottom: vaccinating children aged 5 and above at 80% uptake and at 50% uptake. Top right: Possible trajectories for COVID-19 infections (thousands), hospital admissions and deaths are simulated until December 2022, with different assumptions made for the levels of vaccination coverage in children aged 5 years and above. The shaded areas and solid lines show the 50% and 90% interquantile ranges, and the median for each time point, while the dashed line shows a single sample trajectory. The vertical dotted lines denote the end of model fitting and the beginning of model projections. Full details about the assumptions for each scenario are given in Table 1. Tables: the total number of COVID-19 infections, hospital admissions, and deaths, between 1st May and 31st December 2022, shown to 3 significant figures.

**Figure S8.**
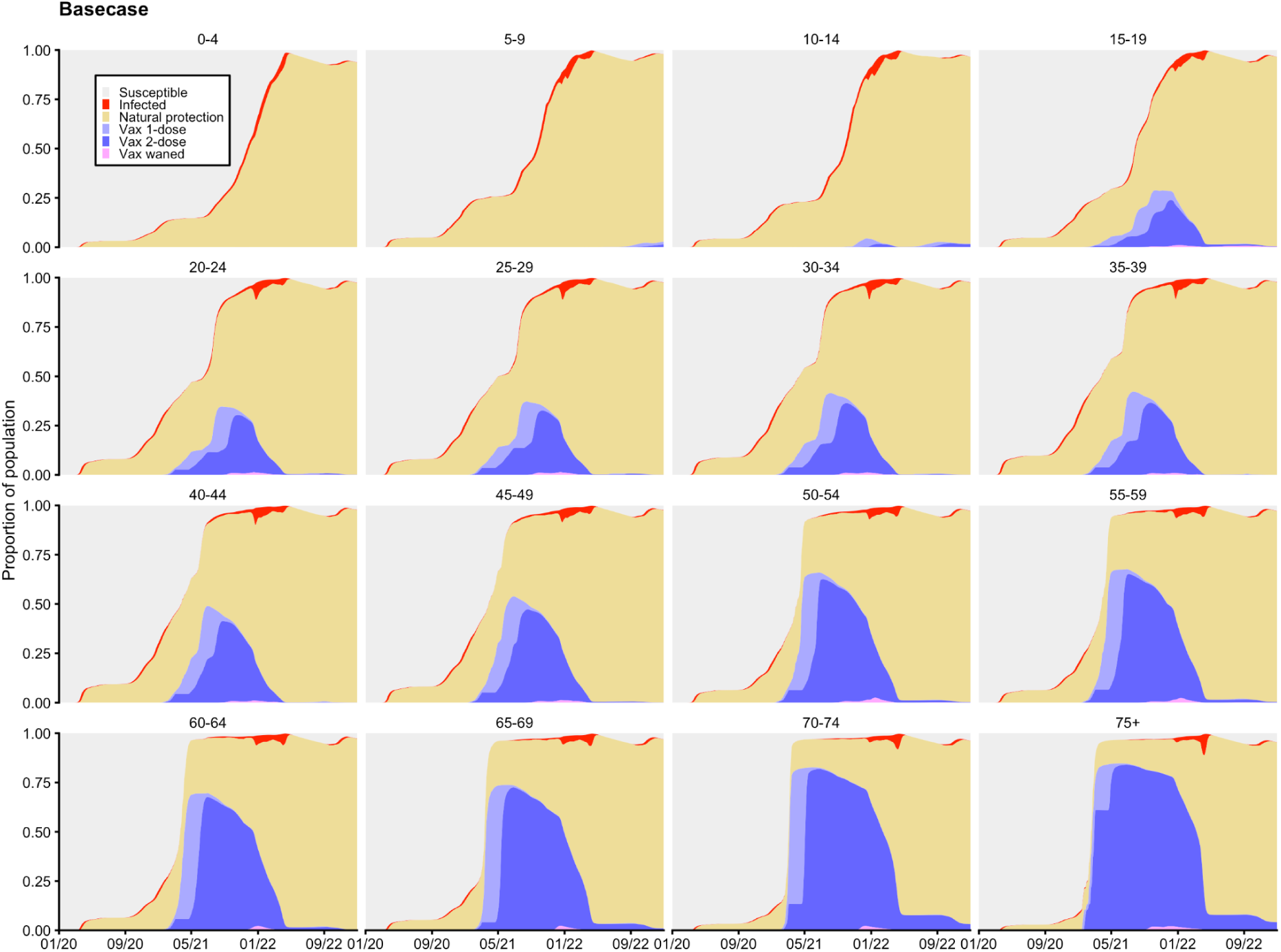
Modelled distribution of disease states over time for each 5-year age group, amalgamated across England from January 2020 to December 2022. The basecase scenario is shown here, which includes the model fitting period (up to 13th May 2022) and the basecase model projection until December 2022. Each panel shows the proportion of individuals in each age group who are: currently susceptible (grey), currently infectious (red), naturally protected (including individuals who have been vaccinated and have natural protection due to infection prior to or after their vaccination) (yellow), vaccine protected with 1 dose (light purple), vaccine protected with 2 doses (including individuals who have received booster doses) (dark purple), and partially vaccine protected with waned vaccine protection approximately six months after the second dose (and having not received any booster vaccination) (pink). Note that, due to waning of both natural and vaccine protection back to the fully-susceptible state, the susceptible proportion does not represent the fraction of each age group that has never been infected or vaccinated, as it includes some people who have been previously infected and/or vaccinated but have completely lost their protection.

**Table S1A.** Fitted model estimates for the relative transmissibility of the Alpha B.1.1.7 variant (compared to pre-existing SARS-CoV-2 variants), Delta B.1.617.2 variant (compared to Alpha), and the Omicron B.1.1.529 variant’s BA.1 sublineage (compared to Delta).

| **NHS England region** | **Alpha B.1.1.7 transmissibility (relative to pre-existing variants)** | **Delta B.1.617.2 transmissibility (relative to Alpha B.1.1.7)** | **Omicron B.1.1.529 sublineage BA.1 transmissibility (relative to Delta B.1.617.2)** |
| --- | --- | --- | --- |
| East of England | 1.862965 | 1.627675 | 1.1351497 |
| London | 1.835242 | 1.583826 | 1.5852839 |
| Midlands | 1.513771 | 1.588105 | 1.1198891 |
| North East & Yorkshire | 1.591191 | 1.740032 | 1.3439809 |
| North West | 2.139368 | 1.657270 | 0.9028017 |
| South East | 1.466895 | 1.657890 | 1.5161782 |
| South West | 1.694188 | 1.527031 | 1.2348388 |

**Table S1B.** Fitted model estimates for the relative transmissibility of the Delta B.1.617.2 variant and the Omicron B.1.1.529 BA.1 sublineage compared to pre-existing SARS-CoV-2 variants (i.e. wild type and D614G). The third column shows the overall relative transmissibility of the Omicron BA.2 sublineage, given the fitted values for previously circulating variants and Omicron sublineage BA.1, and our assumption that BA.2 is 50% more transmissible than BA.1.

| **NHS England region** | **Delta relative to wild type / D614G variants** | **Omicron BA.1 relative to wild type / D614G variants** | **Omicron BA.2 relative to wild type / D614G variants** |
| --- | --- | --- | --- |
| East of England | 3.032302 | 3.442117 | 5.163176 |
| London | 2.906704 | 4.607951 | 6.911926 |
| Midlands | 2.404027 | 2.692244 | 4.038366 |
| North East & Yorkshire | 2.768723 | 3.721111 | 5.581666 |
| North West | 3.545509 | 3.200892 | 4.801338 |
| South East | 2.431951 | 3.687271 | 5.530907 |
| South West | 2.587078 | 3.194625 | 4.791937 |

**Table S2.**
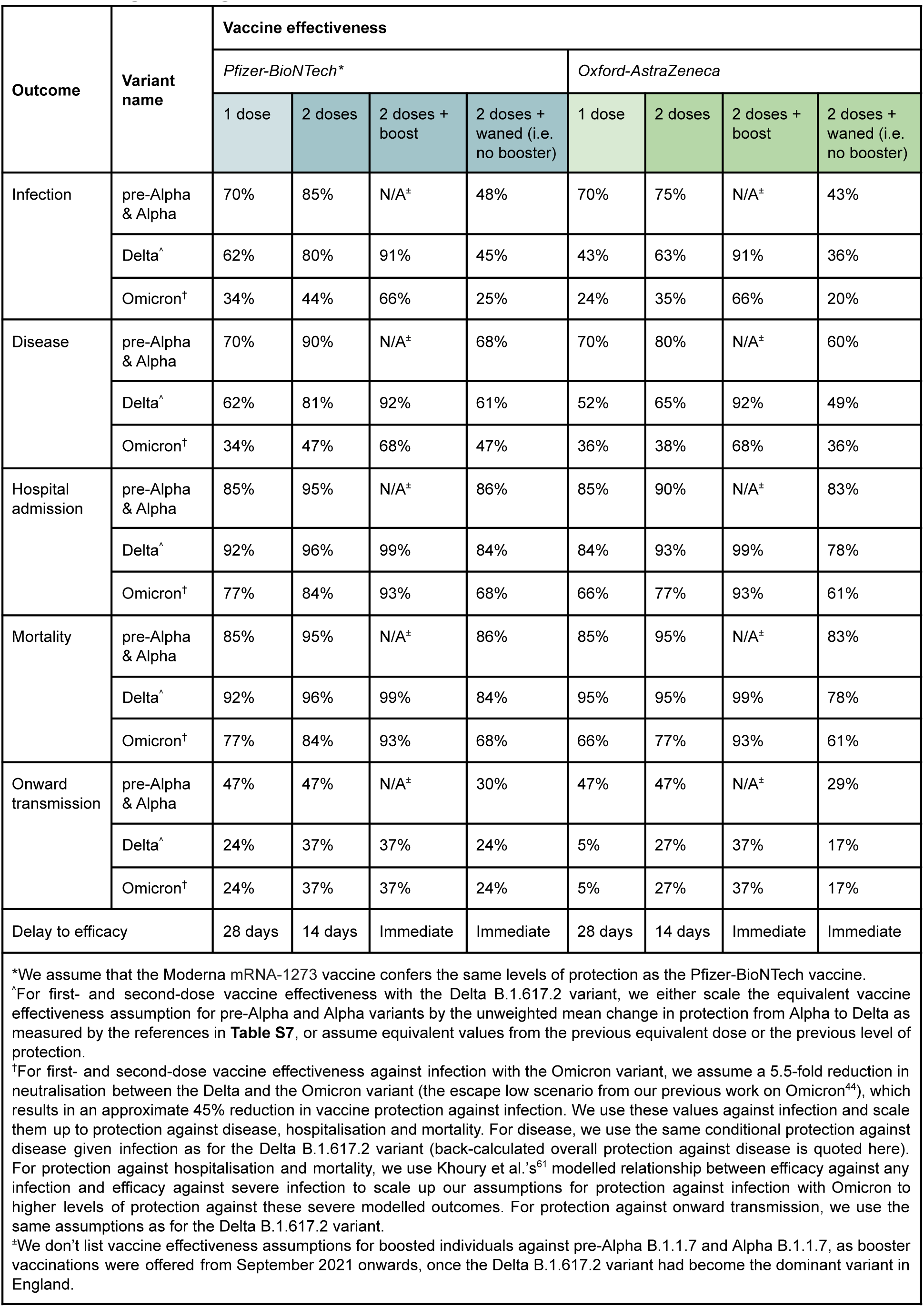
Assumptions for overall vaccine effectiveness against all SARS-CoV-2 outcomes, percentages rounded to 2 significant figures.

**Table S3.** Assumptions for cross protection against SARS-CoV-2 infection and sequential disease outcomes given immunity from a prior infection with SARS-CoV-2

| Outcome | Variant name | Cross protection given immunity from infection with wildtype / pre-Alpha | Cross protection given immunity from infection with Alpha | Cross protection given immunity from infection with Delta | Cross protection given immunity from infection with Omicron |
| --- | --- | --- | --- | --- | --- |
| Infection | pre-Alpha & Alpha | 100% | 100% | 100% | 100% |
|  | Delta | 100% | 100% | 100% | 100% |
|  | Omicron<sup>†</sup> | 55% | 55% | 55% | 100% |
| Disease | pre-Alpha & Alpha | 100% | 100% | 100% | 100% |
|  | Delta | 100% | 100% | 100% | 100% |
|  | Omicron<sup>†</sup> | 57% | 57% | 57% | 100% |
| Hospitalisation | pre-Alpha & Alpha | 100% | 100% | 100% | 100% |
|  | Delta | 100% | 100% | 100% | 100% |
|  | Omicron<sup>†</sup> | 89% | 89% | 89% | 100% |
| Mortality | pre-Alpha & Alpha | 100% | 100% | 100% | 100% |
|  | Delta | 100% | 100% | 100% | 100% |
|  | Omicron<sup>†</sup> | 89% | 89% | 89% | 100% |
| Onward transmission | pre-Alpha & Alpha | 47% (100%)<sup>+</sup> | 47% (100%)<sup>+</sup> | 47% (100%)<sup>+</sup> | 47% (100%)<sup>+</sup> |
|  | Delta | 37% (100%)<sup>+</sup> | 37% (100%)<sup>+</sup> | 37% (100%)<sup>+</sup> | 37% (100%)<sup>+</sup> |
|  | Omicron<sup>†</sup> | 37% | 37% | 37% | 47% (100%)<sup>+</sup> |

\*Where protection against infection is 100%, effective protection against sequential SARS-CoV-2 outcomes (disease, hospital admission, mortality and onward transmission) is also 100%, even if model parameters are set to lower levels of protection.
<sup>†</sup>For cross protection against infection with the Omicron B.1.1.529 variant, we assume a 5.5-fold reduction in neutralisation between the Delta and the Omicron variant (the escape low scenario from our previous work<sup>44</sup>), which results in an approximate 45% reduction in cross protection against infection with Delta B.1.617.2. We use these values against infection and scale them up to protection against disease, hospitalisation and mortality. For disease, we use the same conditional protection against disease given infection as for the Delta B.1.617.2 variant for individuals who were vaccinated with two doses of Pfizer (see **Table S2**) (back-calculated overall protection against disease is quoted here). For cross protection against hospitalisation and mortality, we use Khoury et al.'s<sup>61</sup> modelled relationship between efficacy against any infection and efficacy against severe infection to scale up our assumptions for cross protection against infection with Omicron to higher levels of protection against these severe modelled outcomes. For protection against onward transmission, we use the same assumptions as for the Delta B.1.617.2 variant for individuals who were vaccinated with two doses of Pfizer (see **Table S2**).

**Table S4.** Waning immunity scenarios. Modelling assumptions for the rates of waning immunity. All rates shown here correspond to the rates at which individuals with some form of immunity (either from vaccination or from a prior infection) lose their immunity and return to a fully susceptible disease state. Default waning values are used for the majority of scenarios, including the basecase (see Table 1). The high waning scenario assumes a non-zero rate of waning for individuals with second-dose / second-dose + boosted levels of protection (wva2, wvb2), whereas the central scenario assumes no waning for these categories of individuals. The very high waning scenario assumes the same loss of protection as the high waning scenario but in half the amount of time.

| Parameter name(s) | Description | Default values (central waning) | High waning | Very high waning |
| --- | --- | --- | --- | --- |
| wn, wn2, wn3 | Rate of waning out of recovered compartment and back to susceptible, for all modelled SARS-CoV-2 strains | $\log(0.85)/-365$ corresponding to exponential waning with a 15% loss of protection after 1 year<sup>a</sup> | $\log(0.85)/-365$ corresponding to exponential waning with a 15% loss of protection after 1 year | $\log(0.85)/-182.5$ corresponding to exponential waning with a 15% loss of protection after 6 months |
| wva1 | Rate of waning out of first-dose AstraZeneca vaccinated compartment | 0 | 0 | 0 |
| wva2 | Rate of waning out of second-dose (and second-dose + boosted) AstraZeneca vaccinated compartment | 0 | $\log(0.851)/-140$ | $\log(0.851)/-70$ |
| wva3 | Rate of waning out of second-dose + waned AstraZeneca vaccinated compartment | $\log(0.851)/-140^*$ | $\log(0.851)/-140$ | $\log(0.851)/-70$ |
| wvb1 | Rate of waning out of first-dose mRNA vaccinated compartment | 0 | 0 | 0 |
| wvb2 | Rate of waning out of second-dose (and second-dose + boosted) mRNA vaccinated compartment | 0 | $\log(0.923)/-140$ | $\log(0.923)/-70$ |
| wvb3 | Rate of waning out of second-dose + waned mRNA vaccinated compartment | $\log(0.923)/-140^*$ | $\log(0.923)/-140$ | $\log(0.923)/-70$ |

<sup>a</sup>We referred to a number of studies looking at the level of protection against reinfection with SARS-CoV-2<sup>43</sup>. An unweighted mean across these studies finds approximately 85.74% protection against reinfection with SARS-CoV-2 after 27.76 weeks. When individuals in the model wane, they lose all remaining protection against SARS-CoV-2 outcomes of all types. Our central assumptions therefore assume that the rate at which individuals lose all their protection is slower than that at which reinfections might occur.
\*We used vaccine-specific measured reductions in protection against hospitalisation over 20 weeks from Andrews et al.<sup>19</sup>

**Table S5A.** Details of fitted model parameters. The initial DE-MCMC fitting was done independently for each NHS England region, with 12500 burn-in iterations and 1250 final iterations (13750 iterations total for each region).

| Parameter | Description | Prior distribution or assumed value / distribution | Notes |
| --- | --- | --- | --- |
| tS | Start date of wild-type SARS-CoV-2 epidemic in days after 1 January 2020 | $\sim \text{uniform}(0, 60)$ (i.e. 1st January - 1st March 2020) | Determines the date at which seeding begins in a region; starting on this date, one random individual per day contracts SARS-CoV-2, repeated for 28 days |
| v2_when | Start date of Alpha B.1.1.7 SARS-CoV-2 variant epidemic in days after 1 January 2020 | $\sim \text{uniform}(144, 365)$ (i.e. 24th May - 31st December 2020) | Determines the date at which the novel SARS-CoV-2 variant is introduced into a region; on this date, ten random individuals contract the Alpha B.1.1.7 SARS-CoV-2 variant |
| v3_when | Start date of Delta B.1.617.2 SARS-CoV-2 variant epidemic in days after 1 January 2020 | $\sim \text{uniform}(366, 486)$ (i.e. 1st January - 1st May 2021) | Determines the date at which the novel SARS-CoV-2 variant is introduced into a region; on this date, ten random individuals contract the Delta B.1.617.2 SARS-CoV-2 variant |
| v4_when | Start date of Omicron B.1.1.529 SARS-CoV-2 variant epidemic in days after 1 January 2020 | $\sim \text{normal}(685, 7), \geq 670 \text{ and } \leq 700$ (i.e. 1st November 2021 - 1st December 2021) | Determines the date at which the novel SARS-CoV-2 variant is introduced into a region; on this date, ten random individuals contract the Omicron B.1.1.529 SARS-CoV-2 variant, repeated for 14 days |
| u | Basic susceptibility to infection | $\sim \text{normal}(0.09, 0.02), \geq 0.04 \text{ and } \leq 0.2$ | Determines basic reproduction number $R_0$ |
| v2_relu | Relative transmissibility of Alpha B.1.1.7 variant, compared to pre-existing variants | $\sim \text{lognormal}(0, 0.4), \geq 0.25 \text{ and } \leq 4$ | Determines the transmission advantage that the second SARS-CoV-2 variant in the model, parameterised for the Alpha B.1.1.7 variant, has over pre-existing variants. |
| v3_relu | Relative transmissibility of Delta B.1.617.2 variant, compared to Alpha B.1.1.7 | $\sim \text{lognormal}(0, 0.4), \geq 0.25 \text{ and } \leq 4$ | Determines the transmission advantage that the third SARS-CoV-2 variant in the model, parameterised for the Delta B.1.617.2 variant, has over the Alpha B.1.1.7 variant. |
| v4_relu | Relative transmissibility of Omicron B.1.1.529 variant, compared to Delta B.1.617.2 | $\sim \text{lognormal}(0.4, 0.1), \geq 0.25 \text{ and } \leq 4$ | Determines the transmission advantage that the fourth SARS-CoV-2 variant in the model, parameterised for the Omicron B.1.1.529 variant, has over the Delta B.1.617.2 variant. N.B. although the compartmental model accounts for three SARS-CoV-2 variants in total (see **Figure 1**); we repurpose the first modelled variant (which originally described wildtype and other SARS-CoV-2 variants circulating prior to the Alpha B.1.1.7 variant) for the new Omicron variant on 22nd September 2021. |
| v2_hosp_rlo<br>v2_icu_rlo<br>v2_cfr_rlo | Relative log-odds of hospitalisation, ICU admission and death for the Alpha B.1.1.7 variant, compared to pre-existing variants | $\sim \text{normal}(0, 0.1), \geq -4 \text{ and } \leq 4$ | Vague priors |
| v2_sgtf0 | The proportion of wild-type SARS-CoV-2 that produce S gene target failure (i.e. the “false positive” rate of identifying Alpha B.1.1.7 by SGTF frequency) | $\sim \text{beta}(1.5, 15)$ | Vague prior |
| v2_disp | Controls variance in the distribution for fitting model-predicted Alpha B.1.1.7 frequency to SGTF data | $\sim \text{exponential}(10, 10), \geq 0 \text{ and } \leq 0.25$ | Vague prior |
| v4_sgtf0 | The proportion of non-Omicron SARS-CoV-2 that produce S gene target failure (i.e. the “false positive” rate of identifying Omicron B.1.1.529 by SGTF frequency) | $\sim \text{beta}(1.5, 15)$ | Vague prior |
| v4_disp | Controls variance in the distribution for fitting model-predicted Omicron B.1.1.529 frequency to SGTF data | $\sim \text{exponential}(10, 10), \geq 0 \text{ and } \leq 0.25$ | Vague prior |
| death_mean | Mean delay in days from start of infectious period to death | $\sim \text{normal}(15, 2), \geq 5 \text{ and } \leq 30$ | The delay itself is assumed to follow a gamma distribution with shape parameter 2.2, and mean `death_mean`. Prior and shape of distribution informed by analysis of CO-CIN data<sup>22</sup>. |
| hosp_admission | Mean delay in days from start of infectious period to hospital admission | $\sim \text{normal}(8, 1), \geq 4 \text{ and } \leq 20$ | Delay is assumed to follow a gamma distribution with shape parameter 0.71 and mean `hosp_admission`. Prior and shape of distribution informed by analysis of CO-CIN data<sup>22</sup>. |
| icu_admission | Mean delay in days from start of infectious period to ICU admission | $\sim \text{normal}(12.5, 1), \geq 8 \text{ and } \leq 14$ | Delay is assumed to follow a gamma distribution with shape parameter 1.91 and mean `icu_admission`. Prior and shape of distribution informed by analysis of CO-CIN data<sup>22</sup>. |
| f102, f144, f186, f228, f270, f312, f354, f396, f438, f480, f522, f564, f606, f648, f690, f732, f774, f816 | Contact multiplier for 6-week consecutive periods, with the first 6-week period starting from 12th April 2020 (f102) and the last 6-week period starting from 27th March 2022 (f816). | $\sim \text{lognormal}(0, 0.1), \geq 0.5 \text{ and } \leq 2$ | Vague prior |

**Table S5B.** Model parameters not subject to fitting.

| Parameter | Description | Value | Reference |
| --- | --- | --- | --- |
| $d_E$ | Latent period (E to $I_P$ and E to $I_S$; days) | $\sim \text{gamma}(\mu = 2.5, k = 2.5)$ | Set to 2.5 so that incubation period (latent period plus period of preclinical infectiousness) is 5 days<sup>67</sup> |
| $d_P$ | Duration of preclinical infectiousness ($I_P$ to $I_C$; days) | $\sim \text{gamma}(\mu = 2.5, k = 4)$ | Assumed to be half the duration of total infectiousness in clinically-infected individuals<sup>68</sup> |
| $d_C$ | Duration of clinical infectiousness ($I_C$ to R; days) | $\sim \text{gamma}(\mu = 2.5, k = 4)$ | Infectious period set to 5 days, to result in a serial interval of approximately 6 days<sup>69-71</sup> |
| $d_S$ | Duration of subclinical infectiousness ($I_S$ to R; days) | $\sim \text{gamma}(\mu = 5.0, k = 4)$ | Assumed to be the same duration as total infectious period for clinical cases, including preclinical transmission |
| $y_i$ | Probability of clinical symptoms given infection for age group $i$ | Estimated from case distributions across 6 countries | <sup>29</sup> |
| $f$ | Relative infectiousness of subclinical cases | 50% | Assumed<sup>29,72</sup> |
| $c_{ij}$ | Number of age-$j$ individuals contacted by an age-$i$ individual per day, prior to changes in mobility | UK-specific contact matrix | <sup>58</sup> |
| $N_i$ | Number of age-$i$ individuals | From demographic data | <sup>73</sup> |
| $\Delta t$ | Time step for discrete-time simulation | 0.25 days | |
| $P(ICU)_i$ | Proportion of hospitalised cases that require critical care for age group $i$ | Estimated from CO-CIN data | <sup>74</sup> |
| dVa1 / dVb1 | The duration that individuals have first-dose levels of vaccine protection before transitioning to second-dose levels. Note that we assume a 28-day delay between individuals receiving their first-dose and moving into first-dose levels of protection, and an equivalent 14-day delay before individuals reach second-dose efficacy. These duration assumptions take those delays into account. | Initially,<br>$\sim \text{gamma}(\mu = 7.5, k = 1000)$<br>$7.5 = 21.5 - 28 + 14$<br>Then,<br>$\sim \text{gamma}(\mu = 57.8, k = 1000)$<br>$57.8 = 71.8 - 28 + 14$ | The average delay between first and second vaccine doses was 21.5 days prior to 26th January 2021 when the JCVI updated their guidance<sup>75</sup> on dosing schedules, extending the maximum recommended dosing gap. Following this, the average delay was measured as more than 71 days, using data from October 2021. |
| dVa2 / dVb2 | The duration that individuals have second-dose levels of vaccine protection before receiving a booster vaccine or transitioning to a second-dose + waned state. Note that we assume a 14-day delay before individuals reach second-dose efficacy. These duration assumptions take those delays into account. | $\sim \text{gamma}(\mu = 205, k = 2000)$<br>290 days between 8th December 2020 and 24th September 2021<br>$28 + (71 - 14) = 85 = \text{average } d1:d2 \text{ delay}$<br>$290 - 85 = 205 \text{ days}$ | The JCVI initially recommended a 6-month delay between second doses and booster vaccinations. Vaccine rollout began on 8th December 2020 and booster rollout began in September 2021. |
| $P(\text{boost})$ | The age-specific probability of receiving a COVID-19 booster vaccination, given primary vaccination with either a viral-vector or an mRNA | $P_{\text{BOOST}} = c(0, 0, 0, 0.4, 0.544, 0.597, 0.643, 0.698, 0.757, 0.809, 0.861, 0.892, 0.917, 0.946, 0.964, 0.967)$ | These probabilities are chosen by referring to NHS England data on monthly COVID-19 vaccinations, published on |
|  | vaccine. Probabilities are given in order for each 5-year age group in the model, from 0-4 years up to 70-74 years, and finally for individuals aged 75 years and above. |  | 14th April 2022<sup>24</sup>. We refer to the number of individuals who have received a booster or third vaccine divided by the number of individuals who have received a second vaccine, rounding to 3 decimal places, to inform our assumptions. For the 75+ model age group, we take the average of measured uptake for age groups 75-79 and 80+ in the NHS England data. We assume that individuals aged 16 years and above receive booster vaccinations and that by April 2022, maximum uptake levels have been reached for individuals aged 20 years and above (since these age groups have been eligible to receive boosters since December 2021). For individuals aged 16-19 in the 15-19 years age group, we assume booster uptake will reach 40% across the whole age group, referencing the different levels of uptake already achieved for older age groups (e.g. we calculate 20-24 year olds as having approximately 54% of booster dose uptake in eligible individuals, the lowest level of uptake across all age groups in the NHS England data). |

**Table S6.**
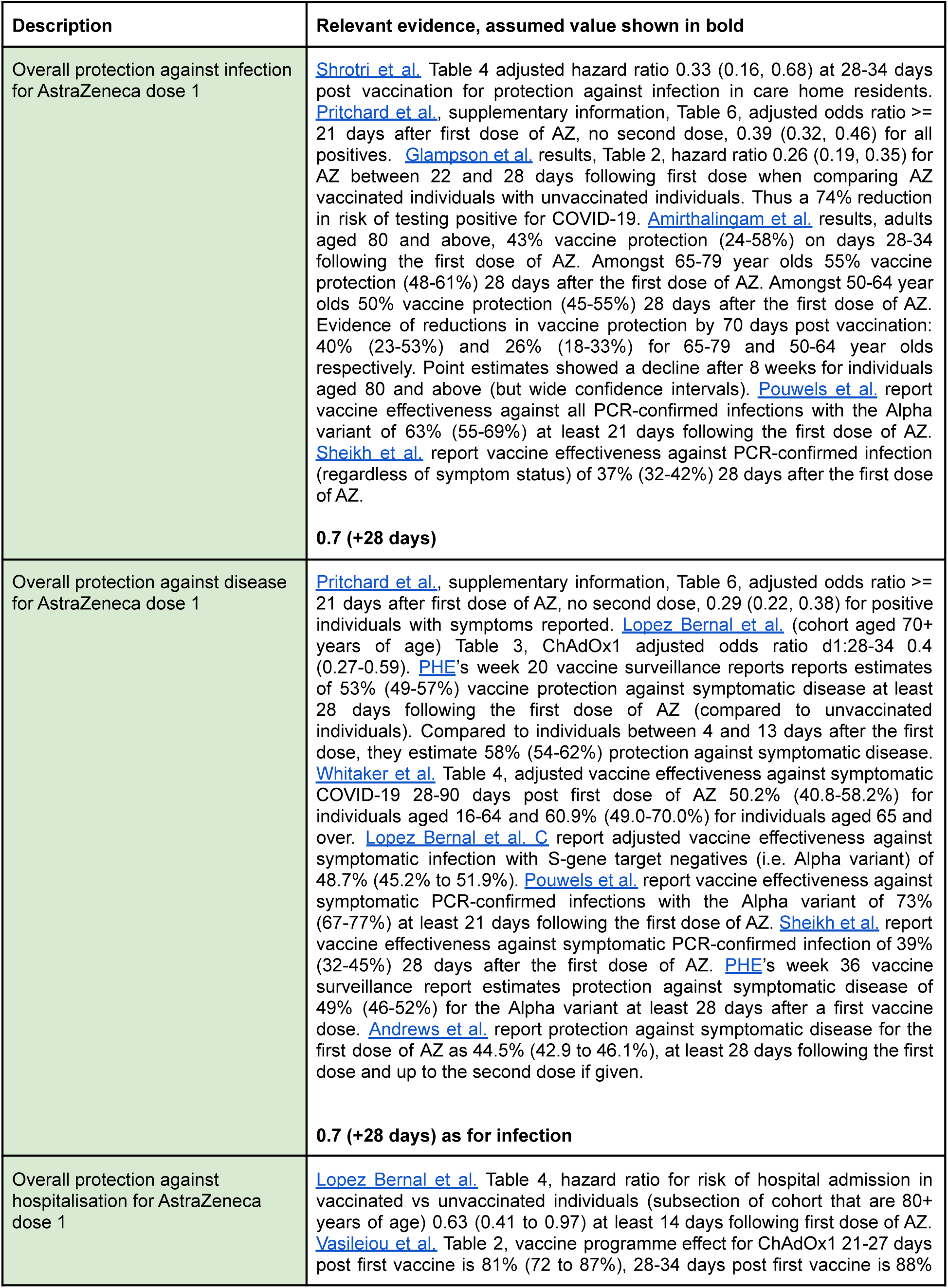
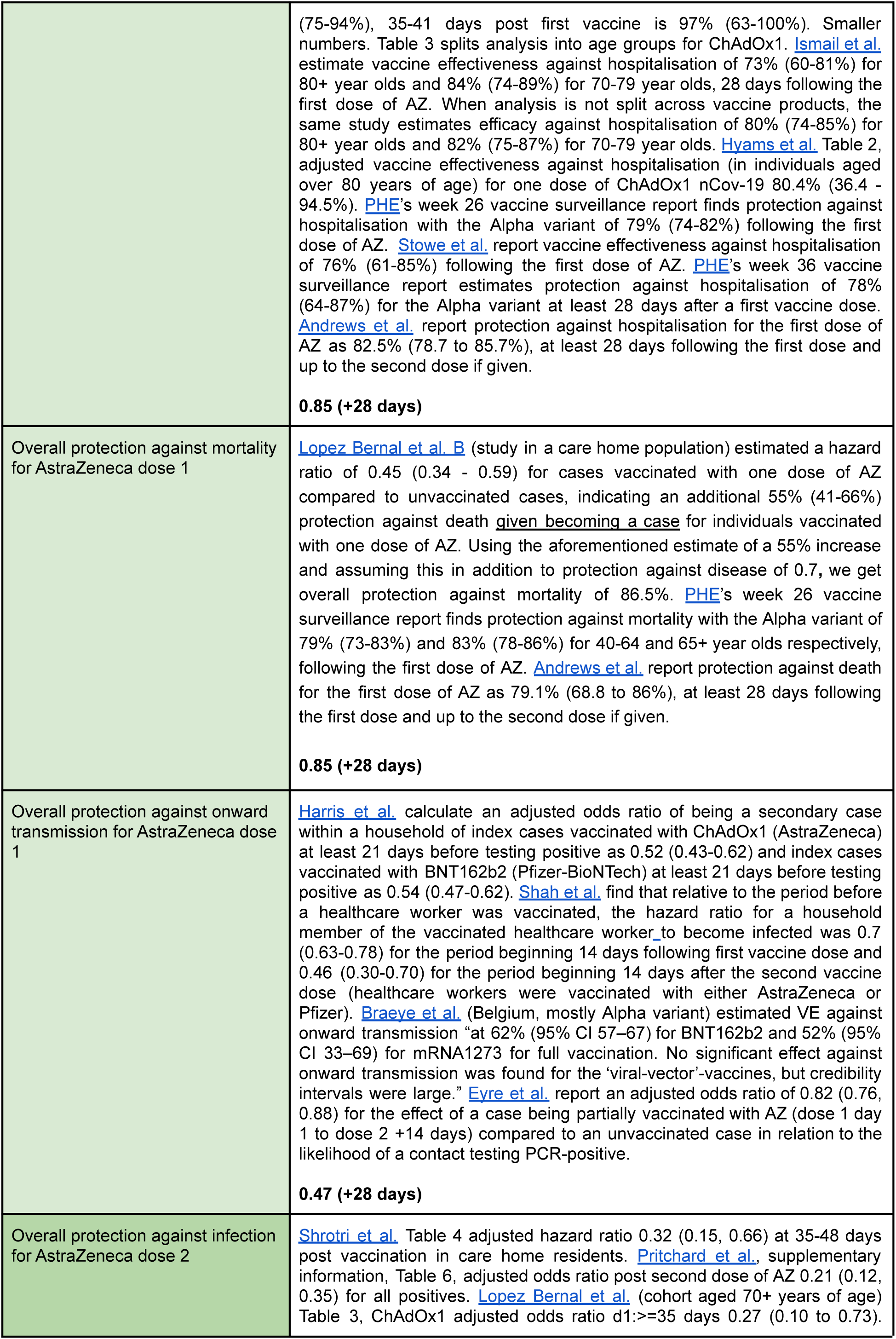
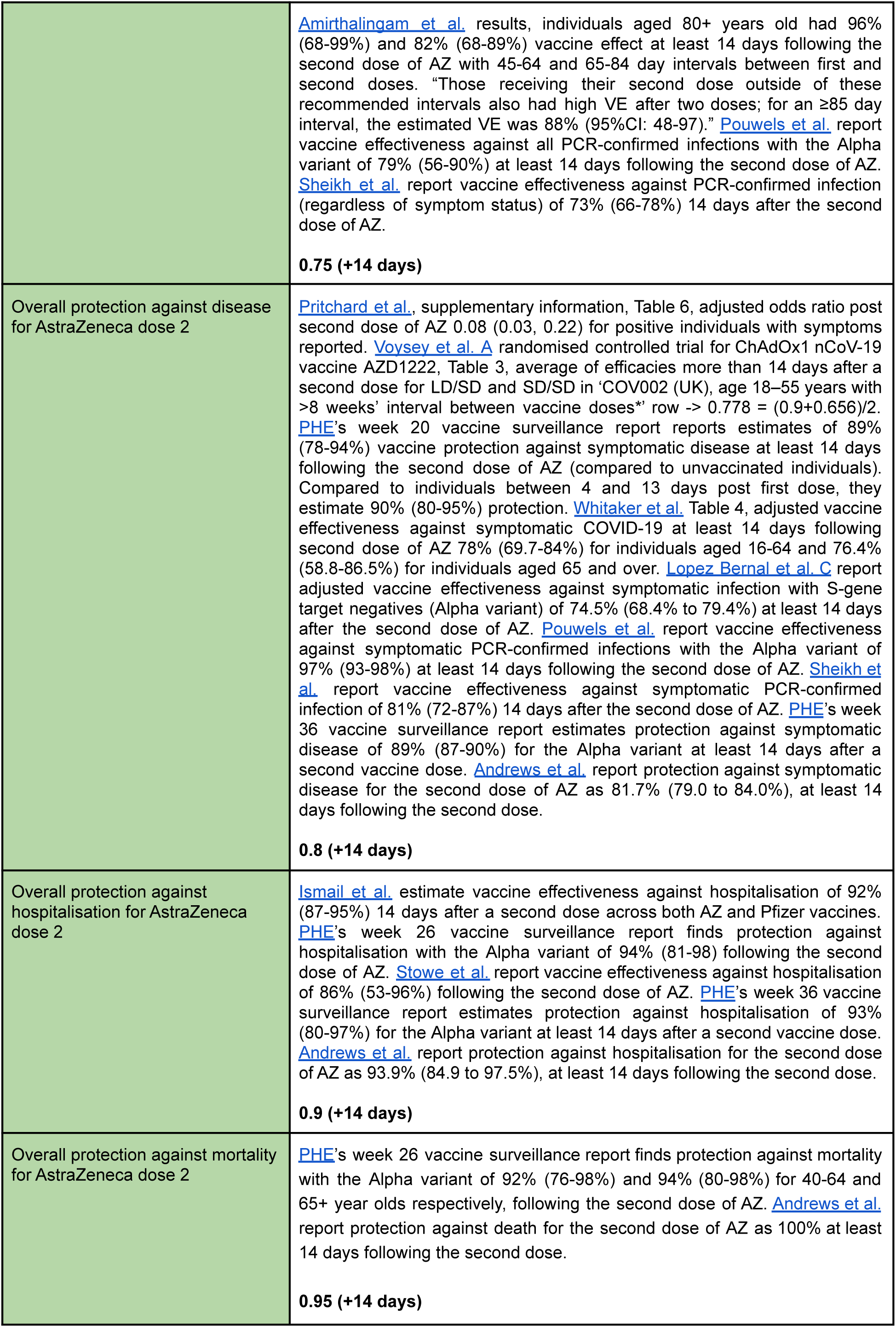
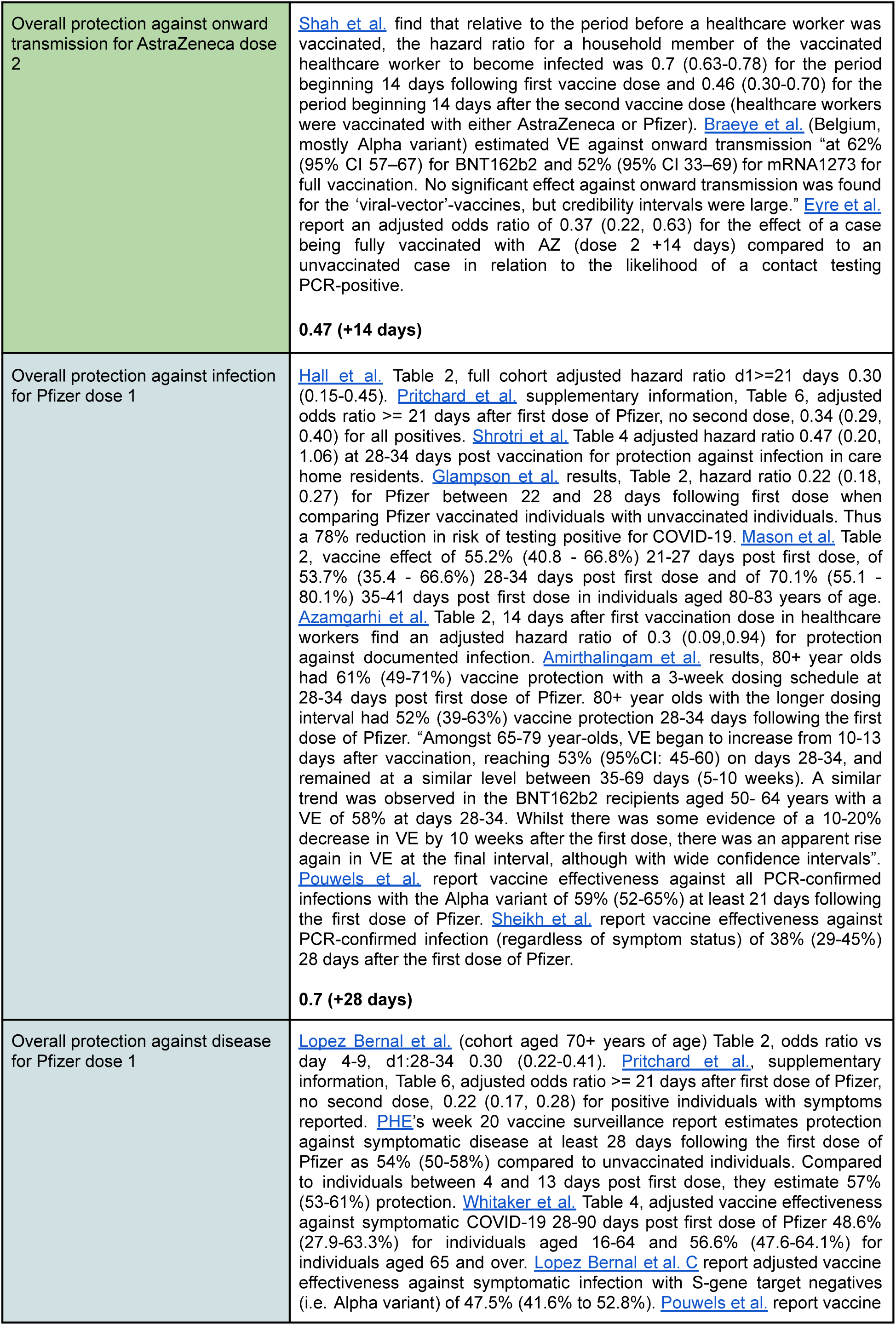
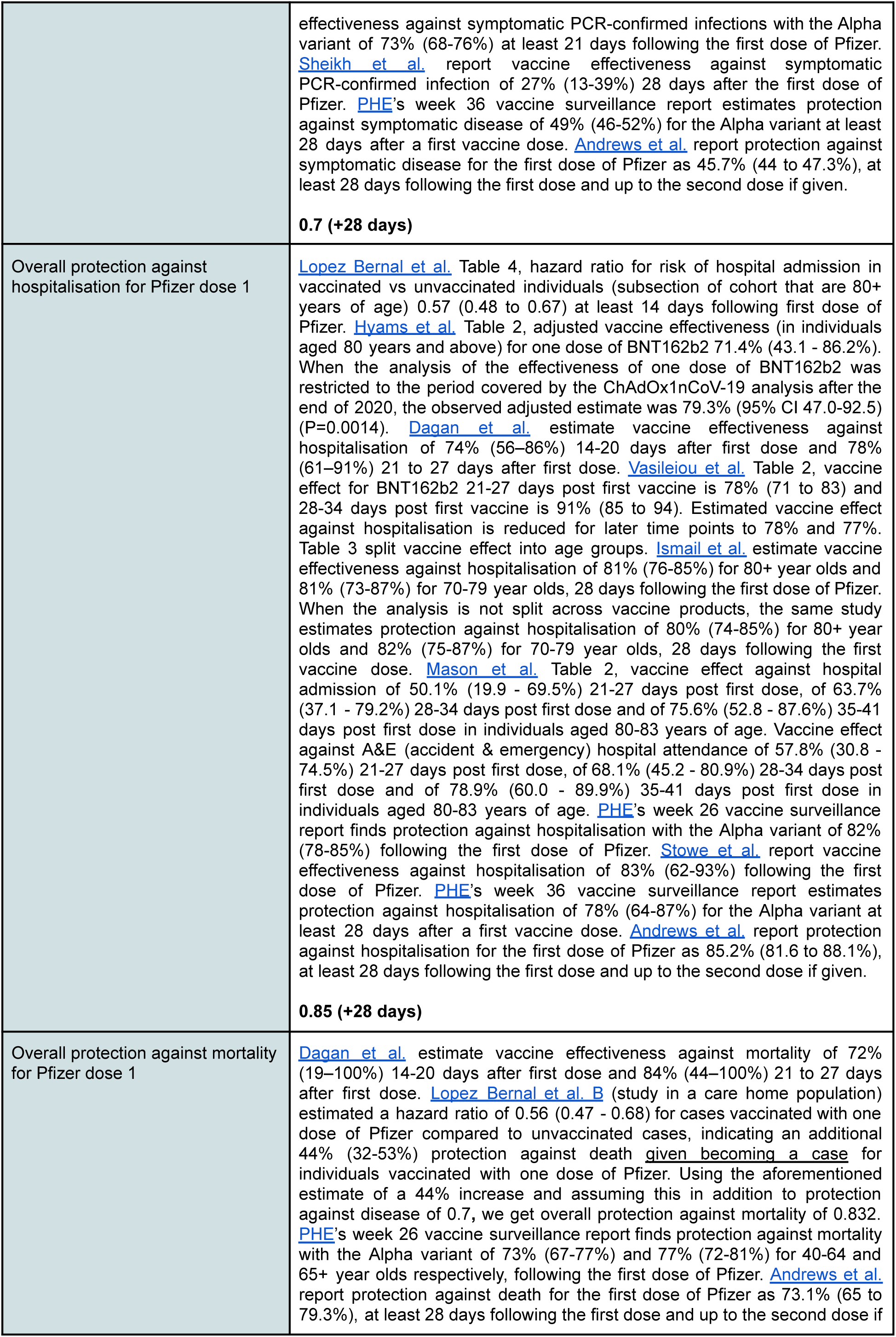
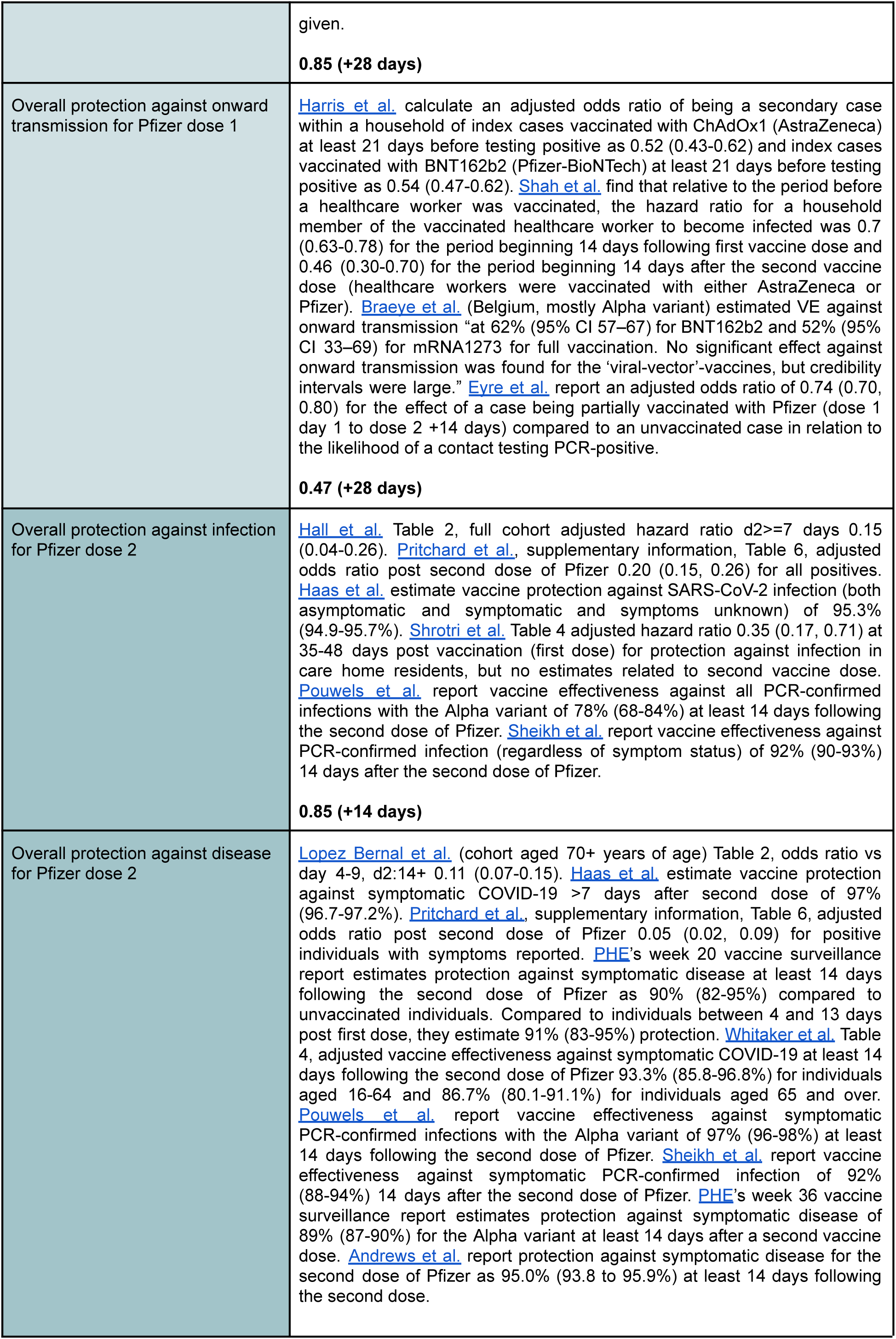
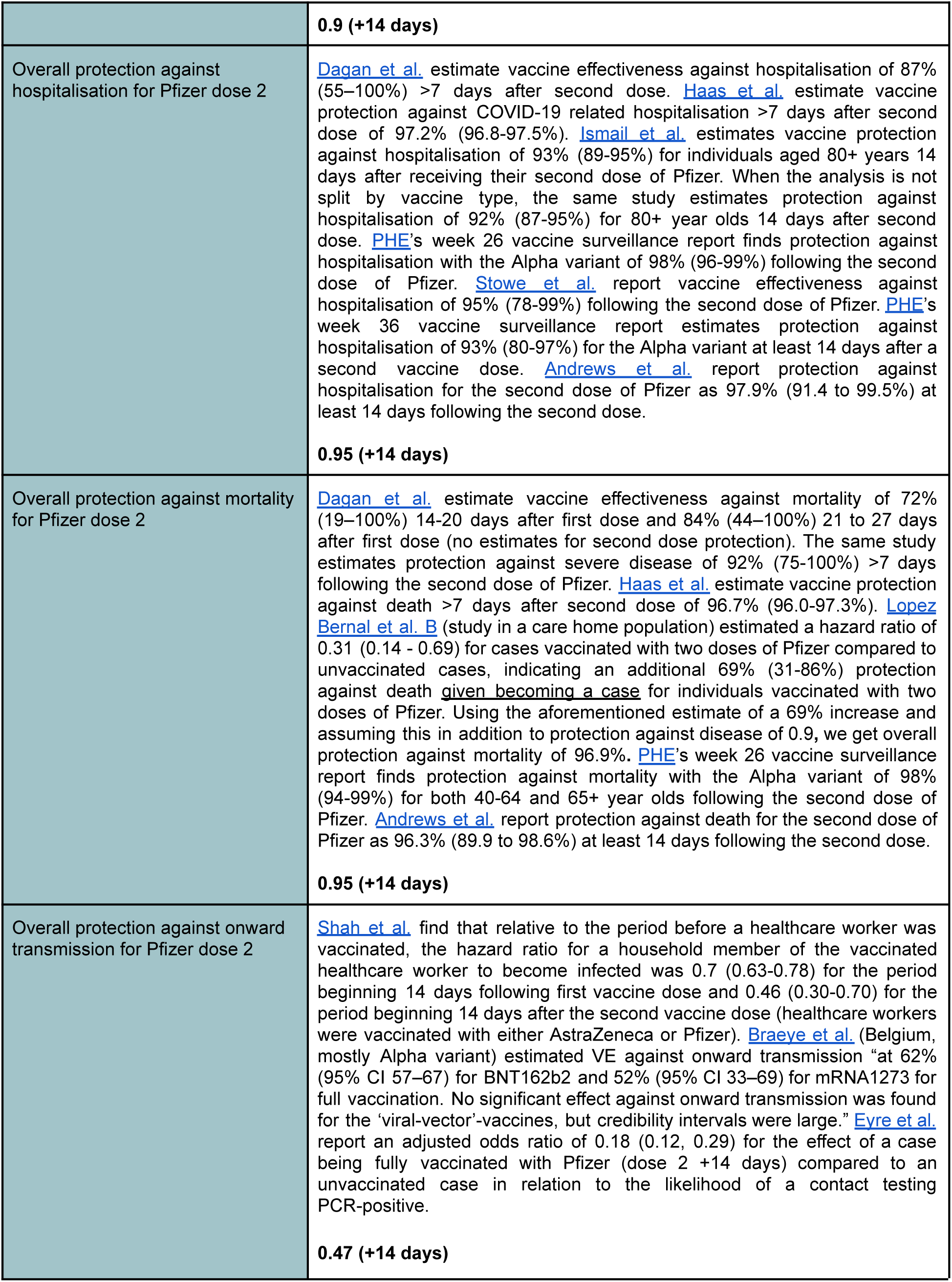
Vaccine effectiveness against pre-B.1.1.7 and Alpha B.1.1.7 variants - relevant evidence and basecase model assumptions

| Description | Relevant evidence, assumed value shown in bold |
| --- | --- |
| Overall protection against infection for AstraZeneca dose 1 | Shrotri et al. Table 4 adjusted hazard ratio 0.33 (0.16, 0.68) at 28-34 days post vaccination for protection against infection in care home residents. Pritchard et al., supplementary information, Table 6, adjusted odds ratio ≥ 21 days after first dose of AZ, no second dose, 0.39 (0.32, 0.46) for all positives. Glampson et al. results, Table 2, hazard ratio 0.26 (0.19, 0.35) for AZ between 22 and 28 days following first dose when comparing AZ vaccinated individuals with unvaccinated individuals. Thus a 74% reduction in risk of testing positive for COVID-19. Amirthalingam et al. results, adults aged 80 and above, 43% vaccine protection (24-58%) on days 28-34 following the first dose of AZ. Amongst 65-79 year olds 55% vaccine protection (48-61%) 28 days after the first dose of AZ. Amongst 50-64 year olds 50% vaccine protection (45-55%) 28 days after the first dose of AZ. Evidence of reductions in vaccine protection by 70 days post vaccination: 40% (23-53%) and 26% (18-33%) for 65-79 and 50-64 year olds respectively. Point estimates showed a decline after 8 weeks for individuals aged 80 and above (but wide confidence intervals). Pouwels et al. report vaccine effectiveness against all PCR-confirmed infections with the Alpha variant of 63% (55-69%) at least 21 days following the first dose of AZ. Sheikh et al. report vaccine effectiveness against PCR-confirmed infection (regardless of symptom status) of 37% (32-42%) 28 days after the first dose of AZ.<br>**0.7 (+28 days)** |
| Overall protection against disease for AstraZeneca dose 1 | Pritchard et al., supplementary information, Table 6, adjusted odds ratio ≥ 21 days after first dose of AZ, no second dose, 0.29 (0.22, 0.38) for positive individuals with symptoms reported. Lopez Bernal et al. (cohort aged 70+ years of age) Table 3, ChAdOx1 adjusted odds ratio d1:28-34 0.4 (0.27-0.59). PHE's week 20 vaccine surveillance reports reports estimates of 53% (49-57%) vaccine protection against symptomatic disease at least 28 days following the first dose of AZ (compared to unvaccinated individuals). Compared to individuals between 4 and 13 days after the first dose, they estimate 58% (54-62%) protection against symptomatic disease. Whitaker et al. Table 4, adjusted vaccine effectiveness against symptomatic COVID-19 28-90 days post first dose of AZ 50.2% (40.8-58.2%) for individuals aged 16-64 and 60.9% (49.0-70.0%) for individuals aged 65 and over. Lopez Bernal et al. C report adjusted vaccine effectiveness against symptomatic infection with S-gene target negatives (i.e. Alpha variant) of 48.7% (45.2% to 51.9%). Pouwels et al. report vaccine effectiveness against symptomatic PCR-confirmed infections with the Alpha variant of 73% (67-77%) at least 21 days following the first dose of AZ. Sheikh et al. report vaccine effectiveness against symptomatic PCR-confirmed infection of 39% (32-45%) 28 days after the first dose of AZ. PHE's week 36 vaccine surveillance report estimates protection against symptomatic disease of 49% (46-52%) for the Alpha variant at least 28 days after a first vaccine dose. Andrews et al. report protection against symptomatic disease for the first dose of AZ as 44.5% (42.9 to 46.1%), at least 28 days following the first dose and up to the second dose if given.<br>**0.7 (+28 days) as for infection** |
| Overall protection against hospitalisation for AstraZeneca dose 1 | Lopez Bernal et al. Table 4, hazard ratio for risk of hospital admission in vaccinated vs unvaccinated individuals (subsection of cohort that are 80+ years of age) 0.63 (0.41 to 0.97) at least 14 days following first dose of AZ. Vasileiou et al. Table 2, vaccine programme effect for ChAdOx1 21-27 days post first vaccine is 81% (72 to 87%), 28-34 days post first vaccine is 88% |
|  | (75-94%), 35-41 days post first vaccine is 97% (63-100%). Smaller numbers. Table 3 splits analysis into age groups for ChAdOx1. Ismail et al. estimate vaccine effectiveness against hospitalisation of 73% (60-81%) for 80+ year olds and 84% (74-89%) for 70-79 year olds, 28 days following the first dose of AZ. When analysis is not split across vaccine products, the same study estimates efficacy against hospitalisation of 80% (74-85%) for 80+ year olds and 82% (75-87%) for 70-79 year olds. Hyams et al. Table 2, adjusted vaccine effectiveness against hospitalisation (in individuals aged over 80 years of age) for one dose of ChAdOx1 nCov-19 80.4% (36.4 - 94.5%). PHE's week 26 vaccine surveillance report finds protection against hospitalisation with the Alpha variant of 79% (74-82%) following the first dose of AZ. Stowe et al. report vaccine effectiveness against hospitalisation of 76% (61-85%) following the first dose of AZ. PHE's week 36 vaccine surveillance report estimates protection against hospitalisation of 78% (64-87%) for the Alpha variant at least 28 days after a first vaccine dose. Andrews et al. report protection against hospitalisation for the first dose of AZ as 82.5% (78.7 to 85.7%), at least 28 days following the first dose and up to the second dose if given.<br>**0.85 (+28 days)** |
| Overall protection against mortality for AstraZeneca dose 1 | Lopez Bernal et al. B (study in a care home population) estimated a hazard ratio of 0.45 (0.34 - 0.59) for cases vaccinated with one dose of AZ compared to unvaccinated cases, indicating an additional 55% (41-66%) protection against death <u>given becoming a case</u> for individuals vaccinated with one dose of AZ. Using the aforementioned estimate of a 55% increase and assuming this in addition to protection against disease of 0.7, we get overall protection against mortality of 86.5%. PHE's week 26 vaccine surveillance report finds protection against mortality with the Alpha variant of 79% (73-83%) and 83% (78-86%) for 40-64 and 65+ year olds respectively, following the first dose of AZ. Andrews et al. report protection against death for the first dose of AZ as 79.1% (68.8 to 86%), at least 28 days following the first dose and up to the second dose if given.<br>**0.85 (+28 days)** |
| Overall protection against onward transmission for AstraZeneca dose 1 | Harris et al. calculate an adjusted odds ratio of being a secondary case within a household of index cases vaccinated with ChAdOx1 (AstraZeneca) at least 21 days before testing positive as 0.52 (0.43-0.62) and index cases vaccinated with BNT162b2 (Pfizer-BioNTech) at least 21 days before testing positive as 0.54 (0.47-0.62). Shah et al. find that relative to the period before a healthcare worker was vaccinated, the hazard ratio for a household member of the vaccinated healthcare worker <u>to become infected</u> was 0.7 (0.63-0.78) for the period beginning 14 days following first vaccine dose and 0.46 (0.30-0.70) for the period beginning 14 days after the second vaccine dose (healthcare workers were vaccinated with either AstraZeneca or Pfizer). Braeye et al. (Belgium, mostly Alpha variant) estimated VE against onward transmission "at 62% (95% CI 57-67) for BNT162b2 and 52% (95% CI 33-69) for mRNA1273 for full vaccination. No significant effect against onward transmission was found for the 'viral-vector'-vaccines, but credibility intervals were large." Eyre et al. report an adjusted odds ratio of 0.82 (0.76, 0.88) for the effect of a case being partially vaccinated with AZ (dose 1 day 1 to dose 2 +14 days) compared to an unvaccinated case in relation to the likelihood of a contact testing PCR-positive.<br>**0.47 (+28 days)** |
| Overall protection against infection for AstraZeneca dose 2 | Shrotri et al. Table 4 adjusted hazard ratio 0.32 (0.15, 0.66) at 35-48 days post vaccination in care home residents. Pritchard et al., supplementary information, Table 6, adjusted odds ratio post second dose of AZ 0.21 (0.12, 0.35) for all positives. Lopez Bernal et al. (cohort aged 70+ years of age) Table 3, ChAdOx1 adjusted odds ratio d1:>=35 days 0.27 (0.10 to 0.73). |
|  | Amirthalingam et al. results, individuals aged 80+ years old had 96% (68-99%) and 82% (68-89%) vaccine effect at least 14 days following the second dose of AZ with 45-64 and 65-84 day intervals between first and second doses. "Those receiving their second dose outside of these recommended intervals also had high VE after two doses; for an $\geq 85$ day interval, the estimated VE was 88% (95%CI: 48-97)." Pouwels et al. report vaccine effectiveness against all PCR-confirmed infections with the Alpha variant of 79% (56-90%) at least 14 days following the second dose of AZ. Sheikh et al. report vaccine effectiveness against PCR-confirmed infection (regardless of symptom status) of 73% (66-78%) 14 days after the second dose of AZ.<br>**0.75 (+14 days)** |
| Overall protection against disease for AstraZeneca dose 2 | Pritchard et al., supplementary information, Table 6, adjusted odds ratio post second dose of AZ 0.08 (0.03, 0.22) for positive individuals with symptoms reported. Voysey et al. A randomised controlled trial for ChAdOx1 nCoV-19 vaccine AZD1222, Table 3, average of efficacies more than 14 days after a second dose for LD/SD and SD/SD in 'COV002 (UK), age 18–55 years with >8 weeks' interval between vaccine doses*' row $\rightarrow 0.778 = (0.9+0.656)/2$. PHE's week 20 vaccine surveillance report reports estimates of 89% (78-94%) vaccine protection against symptomatic disease at least 14 days following the second dose of AZ (compared to unvaccinated individuals). Compared to individuals between 4 and 13 days post first dose, they estimate 90% (80-95%) protection. Whitaker et al. Table 4, adjusted vaccine effectiveness against symptomatic COVID-19 at least 14 days following second dose of AZ 78% (69.7-84%) for individuals aged 16-64 and 76.4% (58.8-86.5%) for individuals aged 65 and over. Lopez Bernal et al. C report adjusted vaccine effectiveness against symptomatic infection with S-gene target negatives (Alpha variant) of 74.5% (68.4% to 79.4%) at least 14 days after the second dose of AZ. Pouwels et al. report vaccine effectiveness against symptomatic PCR-confirmed infections with the Alpha variant of 97% (93-98%) at least 14 days following the second dose of AZ. Sheikh et al. report vaccine effectiveness against symptomatic PCR-confirmed infection of 81% (72-87%) 14 days after the second dose of AZ. PHE's week 36 vaccine surveillance report estimates protection against symptomatic disease of 89% (87-90%) for the Alpha variant at least 14 days after a second vaccine dose. Andrews et al. report protection against symptomatic disease for the second dose of AZ as 81.7% (79.0 to 84.0%), at least 14 days following the second dose.<br>**0.8 (+14 days)** |
| Overall protection against hospitalisation for AstraZeneca dose 2 | Ismail et al. estimate vaccine effectiveness against hospitalisation of 92% (87-95%) 14 days after a second dose across both AZ and Pfizer vaccines. PHE's week 26 vaccine surveillance report finds protection against hospitalisation with the Alpha variant of 94% (81-98) following the second dose of AZ. Stowe et al. report vaccine effectiveness against hospitalisation of 86% (53-96%) following the second dose of AZ. PHE's week 36 vaccine surveillance report estimates protection against hospitalisation of 93% (80-97%) for the Alpha variant at least 14 days after a second vaccine dose. Andrews et al. report protection against hospitalisation for the second dose of AZ as 93.9% (84.9 to 97.5%), at least 14 days following the second dose.<br>**0.9 (+14 days)** |
| Overall protection against mortality for AstraZeneca dose 2 | PHE's week 26 vaccine surveillance report finds protection against mortality with the Alpha variant of 92% (76-98%) and 94% (80-98%) for 40-64 and 65+ year olds respectively, following the second dose of AZ. Andrews et al. report protection against death for the second dose of AZ as 100% at least 14 days following the second dose.<br>**0.95 (+14 days)** |
| Overall protection against onward transmission for AstraZeneca dose 2 | Shah et al. find that relative to the period before a healthcare worker was vaccinated, the hazard ratio for a household member of the vaccinated healthcare worker to become infected was 0.7 (0.63-0.78) for the period beginning 14 days following first vaccine dose and 0.46 (0.30-0.70) for the period beginning 14 days after the second vaccine dose (healthcare workers were vaccinated with either AstraZeneca or Pfizer). Braeye et al. (Belgium, mostly Alpha variant) estimated VE against onward transmission “at 62% (95% CI 57–67) for BNT162b2 and 52% (95% CI 33–69) for mRNA1273 for full vaccination. No significant effect against onward transmission was found for the ‘viral-vector’-vaccines, but credibility intervals were large.” Eyre et al. report an adjusted odds ratio of 0.37 (0.22, 0.63) for the effect of a case being fully vaccinated with AZ (dose 2 +14 days) compared to an unvaccinated case in relation to the likelihood of a contact testing PCR-positive.<br>**0.47 (+14 days)** |
| Overall protection against infection for Pfizer dose 1 | Hall et al. Table 2, full cohort adjusted hazard ratio d1>=21 days 0.30 (0.15-0.45). Pritchard et al. supplementary information, Table 6, adjusted odds ratio >= 21 days after first dose of Pfizer, no second dose, 0.34 (0.29, 0.40) for all positives. Shrotri et al. Table 4 adjusted hazard ratio 0.47 (0.20, 1.06) at 28-34 days post vaccination for protection against infection in care home residents. Glampson et al. results, Table 2, hazard ratio 0.22 (0.18, 0.27) for Pfizer between 22 and 28 days following first dose when comparing Pfizer vaccinated individuals with unvaccinated individuals. Thus a 78% reduction in risk of testing positive for COVID-19. Mason et al. Table 2, vaccine effect of 55.2% (40.8 - 66.8%) 21-27 days post first dose, of 53.7% (35.4 - 66.6%) 28-34 days post first dose and of 70.1% (55.1 - 80.1%) 35-41 days post first dose in individuals aged 80-83 years of age. Azamgarhi et al. Table 2, 14 days after first vaccination dose in healthcare workers find an adjusted hazard ratio of 0.3 (0.09,0.94) for protection against documented infection. Amirthalingam et al. results, 80+ year olds had 61% (49-71%) vaccine protection with a 3-week dosing schedule at 28-34 days post first dose of Pfizer. 80+ year olds with the longer dosing interval had 52% (39-63%) vaccine protection 28-34 days following the first dose of Pfizer. “Amongst 65-79 year-olds, VE began to increase from 10-13 days after vaccination, reaching 53% (95%CI: 45-60) on days 28-34, and remained at a similar level between 35-69 days (5-10 weeks). A similar trend was observed in the BNT162b2 recipients aged 50- 64 years with a VE of 58% at days 28-34. Whilst there was some evidence of a 10-20% decrease in VE by 10 weeks after the first dose, there was an apparent rise again in VE at the final interval, although with wide confidence intervals”. Pouwels et al. report vaccine effectiveness against all PCR-confirmed infections with the Alpha variant of 59% (52-65%) at least 21 days following the first dose of Pfizer. Sheikh et al. report vaccine effectiveness against PCR-confirmed infection (regardless of symptom status) of 38% (29-45%) 28 days after the first dose of Pfizer.<br>**0.7 (+28 days)** |
| Overall protection against disease for Pfizer dose 1 | Lopez Bernal et al. (cohort aged 70+ years of age) Table 2, odds ratio vs day 4-9, d1:28-34 0.30 (0.22-0.41). Pritchard et al. supplementary information, Table 6, adjusted odds ratio >= 21 days after first dose of Pfizer, no second dose, 0.22 (0.17, 0.28) for positive individuals with symptoms reported. PHE's week 20 vaccine surveillance report estimates protection against symptomatic disease at least 28 days following the first dose of Pfizer as 54% (50-58%) compared to unvaccinated individuals. Compared to individuals between 4 and 13 days post first dose, they estimate 57% (53-61%) protection. Whitaker et al. Table 4, adjusted vaccine effectiveness against symptomatic COVID-19 28-90 days post first dose of Pfizer 48.6% (27.9-63.3%) for individuals aged 16-64 and 56.6% (47.6-64.1%) for individuals aged 65 and over. Lopez Bernal et al. C report adjusted vaccine effectiveness against symptomatic infection with S-gene target negatives (i.e. Alpha variant) of 47.5% (41.6% to 52.8%). Pouwels et al. report vaccine |
|  | effectiveness against symptomatic PCR-confirmed infections with the Alpha variant of 73% (68-76%) at least 21 days following the first dose of Pfizer. Sheikh et al. report vaccine effectiveness against symptomatic PCR-confirmed infection of 27% (13-39%) 28 days after the first dose of Pfizer. PHE's week 36 vaccine surveillance report estimates protection against symptomatic disease of 49% (46-52%) for the Alpha variant at least 28 days after a first vaccine dose. Andrews et al. report protection against symptomatic disease for the first dose of Pfizer as 45.7% (44 to 47.3%), at least 28 days following the first dose and up to the second dose if given.<br>**0.7 (+28 days)** |
| Overall protection against hospitalisation for Pfizer dose 1 | Lopez Bernal et al. Table 4, hazard ratio for risk of hospital admission in vaccinated vs unvaccinated individuals (subsection of cohort that are 80+ years of age) 0.57 (0.48 to 0.67) at least 14 days following first dose of Pfizer. Hyams et al. Table 2, adjusted vaccine effectiveness (in individuals aged 80 years and above) for one dose of BNT162b2 71.4% (43.1 - 86.2%). When the analysis of the effectiveness of one dose of BNT162b2 was restricted to the period covered by the ChAdOx1nCoV-19 analysis after the end of 2020, the observed adjusted estimate was 79.3% (95% CI 47.0-92.5) (P=0.0014). Dagan et al. estimate vaccine effectiveness against hospitalisation of 74% (56–86%) 14-20 days after first dose and 78% (61–91%) 21 to 27 days after first dose. Vasileiou et al. Table 2, vaccine effect for BNT162b2 21-27 days post first vaccine is 78% (71 to 83) and 28-34 days post first vaccine is 91% (85 to 94). Estimated vaccine effect against hospitalisation is reduced for later time points to 78% and 77%. Table 3 split vaccine effect into age groups. Ismail et al. estimate vaccine effectiveness against hospitalisation of 81% (76-85%) for 80+ year olds and 81% (73-87%) for 70-79 year olds, 28 days following the first dose of Pfizer. When the analysis is not split across vaccine products, the same study estimates protection against hospitalisation of 80% (74-85%) for 80+ year olds and 82% (75-87%) for 70-79 year olds, 28 days following the first vaccine dose. Mason et al. Table 2, vaccine effect against hospital admission of 50.1% (19.9 - 69.5%) 21-27 days post first dose, of 63.7% (37.1 - 79.2%) 28-34 days post first dose and of 75.6% (52.8 - 87.6%) 35-41 days post first dose in individuals aged 80-83 years of age. Vaccine effect against A&E (accident & emergency) hospital attendance of 57.8% (30.8 - 74.5%) 21-27 days post first dose, of 68.1% (45.2 - 80.9%) 28-34 days post first dose and of 78.9% (60.0 - 89.9%) 35-41 days post first dose in individuals aged 80-83 years of age. PHE's week 26 vaccine surveillance report finds protection against hospitalisation with the Alpha variant of 82% (78-85%) following the first dose of Pfizer. Stowe et al. report vaccine effectiveness against hospitalisation of 83% (62-93%) following the first dose of Pfizer. PHE's week 36 vaccine surveillance report estimates protection against hospitalisation of 78% (64-87%) for the Alpha variant at least 28 days after a first vaccine dose. Andrews et al. report protection against hospitalisation for the first dose of Pfizer as 85.2% (81.6 to 88.1%), at least 28 days following the first dose and up to the second dose if given.<br>**0.85 (+28 days)** |
| Overall protection against mortality for Pfizer dose 1 | Dagan et al. estimate vaccine effectiveness against mortality of 72% (19–100%) 14-20 days after first dose and 84% (44–100%) 21 to 27 days after first dose. Lopez Bernal et al. B (study in a care home population) estimated a hazard ratio of 0.56 (0.47 - 0.68) for cases vaccinated with one dose of Pfizer compared to unvaccinated cases, indicating an additional 44% (32-53%) protection against death <u>given becoming a case</u> for individuals vaccinated with one dose of Pfizer. Using the aforementioned estimate of a 44% increase and assuming this in addition to protection against disease of 0.7, we get overall protection against mortality of 0.832. PHE's week 26 vaccine surveillance report finds protection against mortality with the Alpha variant of 73% (67-77%) and 77% (72-81%) for 40-64 and 65+ year olds respectively, following the first dose of Pfizer. Andrews et al. report protection against death for the first dose of Pfizer as 73.1% (65 to 79.3%), at least 28 days following the first dose and up to the second dose if |
|  | given.<br>**0.85 (+28 days)** |
| Overall protection against onward transmission for Pfizer dose 1 | Harris et al. calculate an adjusted odds ratio of being a secondary case within a household of index cases vaccinated with ChAdOx1 (AstraZeneca) at least 21 days before testing positive as 0.52 (0.43-0.62) and index cases vaccinated with BNT162b2 (Pfizer-BioNTech) at least 21 days before testing positive as 0.54 (0.47-0.62). Shah et al. find that relative to the period before a healthcare worker was vaccinated, the hazard ratio for a household member of the vaccinated healthcare worker to become infected was 0.7 (0.63-0.78) for the period beginning 14 days following first vaccine dose and 0.46 (0.30-0.70) for the period beginning 14 days after the second vaccine dose (healthcare workers were vaccinated with either AstraZeneca or Pfizer). Braeye et al. (Belgium, mostly Alpha variant) estimated VE against onward transmission "at 62% (95% CI 57–67) for BNT162b2 and 52% (95% CI 33–69) for mRNA1273 for full vaccination. No significant effect against onward transmission was found for the 'viral-vector'-vaccines, but credibility intervals were large." Eyre et al. report an adjusted odds ratio of 0.74 (0.70, 0.80) for the effect of a case being partially vaccinated with Pfizer (dose 1 day 1 to dose 2 +14 days) compared to an unvaccinated case in relation to the likelihood of a contact testing PCR-positive.<br>**0.47 (+28 days)** |
| Overall protection against infection for Pfizer dose 2 | Hall et al. Table 2, full cohort adjusted hazard ratio d2>=7 days 0.15 (0.04-0.26). Pritchard et al., supplementary information, Table 6, adjusted odds ratio post second dose of Pfizer 0.20 (0.15, 0.26) for all positives. Haas et al. estimate vaccine protection against SARS-CoV-2 infection (both asymptomatic and symptomatic and symptoms unknown) of 95.3% (94.9-95.7%). Shrotri et al. Table 4 adjusted hazard ratio 0.35 (0.17, 0.71) at 35-48 days post vaccination (first dose) for protection against infection in care home residents, but no estimates related to second vaccine dose. Pouwels et al. report vaccine effectiveness against all PCR-confirmed infections with the Alpha variant of 78% (68-84%) at least 14 days following the second dose of Pfizer. Sheikh et al. report vaccine effectiveness against PCR-confirmed infection (regardless of symptom status) of 92% (90-93%) 14 days after the second dose of Pfizer.<br>**0.85 (+14 days)** |
| Overall protection against disease for Pfizer dose 2 | Lopez Bernal et al. (cohort aged 70+ years of age) Table 2, odds ratio vs day 4-9, d2:14+ 0.11 (0.07-0.15). Haas et al. estimate vaccine protection against symptomatic COVID-19 >7 days after second dose of 97% (96.7-97.2%). Pritchard et al., supplementary information, Table 6, adjusted odds ratio post second dose of Pfizer 0.05 (0.02, 0.09) for positive individuals with symptoms reported. PHE's week 20 vaccine surveillance report estimates protection against symptomatic disease at least 14 days following the second dose of Pfizer as 90% (82-95%) compared to unvaccinated individuals. Compared to individuals between 4 and 13 days post first dose, they estimate 91% (83-95%) protection. Whitaker et al. Table 4, adjusted vaccine effectiveness against symptomatic COVID-19 at least 14 days following the second dose of Pfizer 93.3% (85.8-96.8%) for individuals aged 16-64 and 86.7% (80.1-91.1%) for individuals aged 65 and over. Pouwels et al. report vaccine effectiveness against symptomatic PCR-confirmed infections with the Alpha variant of 97% (96-98%) at least 14 days following the second dose of Pfizer. Sheikh et al. report vaccine effectiveness against symptomatic PCR-confirmed infection of 92% (88-94%) 14 days after the second dose of Pfizer. PHE's week 36 vaccine surveillance report estimates protection against symptomatic disease of 89% (87-90%) for the Alpha variant at least 14 days after a second vaccine dose. Andrews et al. report protection against symptomatic disease for the second dose of Pfizer as 95.0% (93.8 to 95.9%) at least 14 days following the second dose. |
|  | **0.9 (+14 days)** |
| Overall protection against hospitalisation for Pfizer dose 2 | Dagan et al. estimate vaccine effectiveness against hospitalisation of 87% (55–100%) >7 days after second dose. Haas et al. estimate vaccine protection against COVID-19 related hospitalisation >7 days after second dose of 97.2% (96.8-97.5%). Ismail et al. estimates vaccine protection against hospitalisation of 93% (89-95%) for individuals aged 80+ years 14 days after receiving their second dose of Pfizer. When the analysis is not split by vaccine type, the same study estimates protection against hospitalisation of 92% (87-95%) for 80+ year olds 14 days after second dose. PHE's week 26 vaccine surveillance report finds protection against hospitalisation with the Alpha variant of 98% (96-99%) following the second dose of Pfizer. Stowe et al. report vaccine effectiveness against hospitalisation of 95% (78-99%) following the second dose of Pfizer. PHE's week 36 vaccine surveillance report estimates protection against hospitalisation of 93% (80-97%) for the Alpha variant at least 14 days after a second vaccine dose. Andrews et al. report protection against hospitalisation for the second dose of Pfizer as 97.9% (91.4 to 99.5%) at least 14 days following the second dose.<br>**0.95 (+14 days)** |
| Overall protection against mortality for Pfizer dose 2 | Dagan et al. estimate vaccine effectiveness against mortality of 72% (19–100%) 14–20 days after first dose and 84% (44–100%) 21 to 27 days after first dose (no estimates for second dose protection). The same study estimates protection against severe disease of 92% (75-100%) >7 days following the second dose of Pfizer. Haas et al. estimate vaccine protection against death >7 days after second dose of 96.7% (96.0-97.3%). Lopez Bernal et al. B (study in a care home population) estimated a hazard ratio of 0.31 (0.14 - 0.69) for cases vaccinated with two doses of Pfizer compared to unvaccinated cases, indicating an additional 69% (31-86%) protection against death <u>given becoming a case</u> for individuals vaccinated with two doses of Pfizer. Using the aforementioned estimate of a 69% increase and assuming this in addition to protection against disease of 0.9, we get overall protection against mortality of 96.9%. PHE's week 26 vaccine surveillance report finds protection against mortality with the Alpha variant of 98% (94-99%) for both 40-64 and 65+ year olds following the second dose of Pfizer. Andrews et al. report protection against death for the second dose of Pfizer as 96.3% (89.9 to 98.6%) at least 14 days following the second dose.<br>**0.95 (+14 days)** |
| Overall protection against onward transmission for Pfizer dose 2 | Shah et al. find that relative to the period before a healthcare worker was vaccinated, the hazard ratio for a household member of the vaccinated healthcare worker to become infected was 0.7 (0.63-0.78) for the period beginning 14 days following first vaccine dose and 0.46 (0.30-0.70) for the period beginning 14 days after the second vaccine dose (healthcare workers were vaccinated with either AstraZeneca or Pfizer). Braeye et al. (Belgium, mostly Alpha variant) estimated VE against onward transmission “at 62% (95% CI 57–67) for BNT162b2 and 52% (95% CI 33–69) for mRNA1273 for full vaccination. No significant effect against onward transmission was found for the ‘viral-vector’-vaccines, but credibility intervals were large.” Eyre et al. report an adjusted odds ratio of 0.18 (0.12, 0.29) for the effect of a case being fully vaccinated with Pfizer (dose 2 +14 days) compared to an unvaccinated case in relation to the likelihood of a contact testing PCR-positive.<br>**0.47 (+14 days)** |

**Table S7.** Vaccine effectiveness against B.1.617.2 Delta variant - relevant evidence and baseline model assumptions

| Description | Relevant evidence, assumed value shown in bold |
| --- | --- |
| Overall protection against infection for AstraZeneca dose 1 | Pouwels et al. report vaccine effectiveness against all PCR-confirmed infections with the Delta variant of 46% (35-55%) at least 21 days following the first dose of AZ. This is a **26.98% reduction** on their equivalent estimate for the Alpha variant. Sheikh et al. report vaccine effectiveness against PCR-confirmed infection (regardless of symptom status) of 18% (9-25%) 28 days after the first dose of AZ. This is a **51.35% reduction** on their equivalent estimate for the Alpha variant.<br>**Alpha assumption 0.7**<br>**0.43 = 0.7 \* (1 - 0.39) (+28 days)** |
| Overall protection against disease for AstraZeneca dose 1 | Lopez Bernal et al. C report adjusted vaccine effectiveness against symptomatic infection with S-gene target positives (Delta variant) of 30.0% (24.3-35.3%) at least 21 days after the first dose of AZ. A **38.4% reduction** on their estimate of equivalent vaccine protection against the Alpha variant. Pouwels et al. report vaccine effectiveness against symptomatic PCR-confirmed infections with the Delta variant of 40% (28-50%) at least 21 days following the first dose of AZ. This is a **45.21% reduction** on their equivalent estimate for the Alpha variant. Sheikh et al. report vaccine effectiveness against symptomatic PCR-confirmed infection of 33% (23-41%) 28 days after the first dose of AZ. This is a **15.38% reduction** on their equivalent estimate for the Alpha variant. PHE's week 36 vaccine surveillance report estimates protection against symptomatic disease of 35% (32-38%) for the Delta variant at least 28 days after a first vaccine dose. This is a **28.57% reduction** compared to their equivalent estimate for Alpha. Andrews et al. report protection against symptomatic disease for the first dose of AZ as 43.3% (42.3 to 44.2%), at least 28 days following the first dose and up to the second dose if given. This is a **2.7% reduction** compared to their equivalent estimate for Alpha.<br>**Alpha assumption 0.7**<br>**0.52 = 0.7 \* (1-0.26) (+28 days)** |
| Overall protection against hospitalisation for AstraZeneca dose 1 | Stowe et al. report vaccine effectiveness against hospitalisation of 71% (51-83%) following the first dose of AZ. This is a **6.58% reduction** on their equivalent estimate for the Alpha variant. PHE's week 36 vaccine surveillance report estimates protection against hospitalisation of 80% (69-88%) for the Delta variant at least 28 days after a first vaccine dose. This is a **2.56% increase** compared to their equivalent estimate for Alpha. Andrews et al. report protection against hospitalisation for the first dose of AZ as 81.4% (78.7 to 83.7%), at least 28 days following the first dose and up to the second dose if given. This is a **1.3% reduction** compared to their equivalent estimate for Alpha.<br>**Alpha assumption 0.85**<br>**0.84 = 0.85 \* (1-0.017) (+28 days)** |
| Overall protection against mortality for AstraZeneca dose 1 | Andrews et al. report protection against death for the first dose of AZ as 88.4% (78.2 to 93.8%), at least 28 days following the first dose and up to the second dose if given. This is a **11.8% increase** compared to their equivalent |
|  | estimate for Alpha.<br>**Alpha assumption 0.85**<br>**$0.95 = 0.85 * (1 + 0.118) (+28 \text{ days})$** |
| Overall protection against onward transmission for AstraZeneca dose 1 | Eyre et al. report an adjusted odds ratio of 0.98 (0.90, 1.06) for the effect of a case being partially vaccinated with AZ (dose 1 day 1 to dose 2 +14 days) compared to an unvaccinated case in relation to the likelihood of a contact testing PCR-positive (N.B. this estimate has a non-significant p-value), equivalent to vaccine protection of 2%. Their equivalent estimate for Alpha is 0.82 (0.76, 0.88), therefore vaccine protection of 18%. This is a **88.9% overall reduction** in vaccine effect from Alpha to Delta.<br>**Alpha assumption 0.47**<br>**$0.05 = 0.47 * (1 - 0.889) (+28 \text{ days})$** |
| Overall protection against infection for AstraZeneca dose 2 | Pouwels et al. report vaccine effectiveness against all PCR-confirmed infections with the Delta variant of 67% (62-71%) at least 14 days following the second dose of AZ. This is a **15.19% reduction** on their equivalent estimate for the Alpha variant. Sheikh et al. report vaccine effectiveness against PCR-confirmed infection (regardless of symptom status) of 60% (53-66%) 14 days after the second dose of AZ. This is a **17.81% reduction** on their equivalent estimate for the Alpha variant.<br>**Alpha assumption 0.75**<br>**$0.63 = 0.75 * (1 - 0.165) (+14 \text{ days})$** |
| Overall protection against disease for AstraZeneca dose 2 | Lopez Bernal et al. C report adjusted vaccine effectiveness against symptomatic infection with S-gene target positives (Delta variant) of 67.0% (61.3% to 71.8%) at least 14 days after the second dose of AZ. A **10.07% reduction** on their estimate of equivalent vaccine protection against the Alpha variant. Pouwels et al. report vaccine effectiveness against symptomatic PCR-confirmed infections with the Delta variant of 71% (66-74%) at least 14 days following the second dose of AZ. This is a **26.8% reduction** on their equivalent estimate for the Alpha variant. Sheikh et al. report vaccine effectiveness against symptomatic PCR-confirmed infection of 61% (51-70%) 14 days after the second dose of AZ. This is a **24.69% reduction** on their equivalent estimate for the Alpha variant. PHE's week 36 vaccine surveillance report estimates protection against symptomatic disease of 79% (78-80%) for the Delta variant at least 14 days after a second vaccine dose. This is an **11.24% reduction** compared to their equivalent estimate for Alpha. Andrews et al. report protection against symptomatic disease for the second dose of AZ as 65.2% (64.9 to 65.6%), at least 14 days following the second dose. This is a **20.2% reduction** compared to their equivalent estimate for Alpha.<br>**Alpha assumption 0.8**<br>**$0.65 = 0.8 * (1 - 0.186) (+14 \text{ days})$** |
| Overall protection against hospitalisation for AstraZeneca dose 2 | Stowe et al. report vaccine effectiveness against hospitalisation of 92% (75-97%) following the second dose of AZ. This is a **6.98% increase** on their equivalent estimate for the Alpha variant. PHE's week 36 vaccine surveillance report estimates protection against hospitalisation of 96% (91-98%) for the Delta variant at least 14 days after a second vaccine dose. |
|  | This is a **3.23% increase** compared to their equivalent estimate for Alpha. Andrews et al. report protection against hospitalisation for the second dose of AZ as 93.0% (92.4 to 93.5%), at least 14 days following the second dose. This is a **1% reduction** compared to their equivalent estimate for Alpha.<br>**Alpha assumption 0.9**<br>**0.93 = 0.9 \* (1 + 0.0307) (+14 days)** |
| Overall protection against mortality for AstraZeneca dose 2 | Andrews et al. report protection against death for the second dose of AZ as 92.7% (90.7 to 94.3%), at least 14 days following the second dose. This is a **7.3% reduction** compared to their equivalent estimate for Alpha.<br>**Alpha assumption 0.95**<br>**0.95 (+14 days), as for dose 1 protection against mortality** |
| Overall protection against onward transmission for AstraZeneca dose 2 | Eyre et al. report an adjusted odds ratio of 0.64 (0.57, 0.72) for the effect of a case being fully vaccinated with AZ (dose 2 +14 days) compared to an unvaccinated case in relation to the likelihood of a contact testing PCR-positive, equivalent to vaccine protection of 36%. Their equivalent estimate for Alpha is 0.37 (0.22, 0.63), therefore vaccine protection of 63%. This is a **42.9% overall reduction** in vaccine effect from Alpha to Delta.<br>**Alpha assumption 0.47**<br>**0.27 = 0.47 \* (1 - 0.429) (+14 days)** |
| Overall protection against infection for Pfizer dose 1 | Pouwels et al. report vaccine effectiveness against all PCR-confirmed infections with the Delta variant of 57% (50-63%) at least 21 days following the first dose of Pfizer. This is a **3.39% reduction** on their equivalent estimate for the Alpha variant. Sheikh et al. report vaccine effectiveness against PCR-confirmed infection (regardless of symptom status) of 30% (17-41%) 28 days after the first dose of Pfizer. This is a **21.05% reduction** on their equivalent estimate for the Alpha variant.<br>**Alpha assumption 0.7**<br>**0.62 = 0.7 \* (1-0.12) (+28 days)** |
| Overall protection against disease for Pfizer dose 1 | Lopez Bernal et al. C report adjusted vaccine effectiveness against symptomatic infection with S-gene target positives (Delta variant) of 35.6% (22.7% to 46.4%) at least 21 days after the first dose of Pfizer. A **25.05% reduction** on their estimate of equivalent vaccine protection against the Alpha variant. Pouwels et al. report vaccine effectiveness against symptomatic PCR-confirmed infections with the Delta variant of 58% (51-64%) at least 21 days following the first dose of Pfizer. This is a **20.55% reduction** on their equivalent estimate for the Alpha variant. Sheikh et al. report vaccine effectiveness against symptomatic PCR-confirmed infection of 33% (15-47%) 28 days after the first dose of Pfizer. This is a **22.22% reduction** on their equivalent estimate for the Alpha variant. PHE's week 36 vaccine surveillance report estimates protection against symptomatic disease of 35% (32-38%) for the Delta variant at least 28 days after a first vaccine dose. This is a **28.57% reduction** compared to their equivalent estimate for Alpha. Andrews et al. report protection against symptomatic disease for the first dose of Pfizer as 51.9% (51.4 to 52.4%), at least 28 days following the first dose and up to the second dose if given. This is a **13.6% increase** compared to their equivalent estimate for Alpha. |
|  | **Alpha assumption 0.7**<br>**0.62 (+28 days) as for infection, otherwise would be 0.58** |
| Overall protection against hospitalisation for Pfizer dose 1 | Stowe et al. report vaccine effectiveness against hospitalisation of 94% (46-99%) following the first dose of Pfizer. This is a **13.25% increase** on their equivalent estimate for the Alpha variant. PHE's week 36 vaccine surveillance report estimates protection against hospitalisation of 80% (69-88%) for the Delta variant at least 28 days after a first vaccine dose. This is a **2.56% increase** compared to their equivalent estimate for Alpha. Andrews et al. report protection against hospitalisation for the first dose of Pfizer as 91.8% (90.4 to 93%), at least 28 days following the first dose and up to the second dose if given. This is a **7.7% increase** compared to their equivalent estimate for Alpha.<br>**Alpha assumption 0.85**<br>**0.92 = 0.85 \* (1+0.078) (+28 days)** |
| Overall protection against mortality for Pfizer dose 1 | Andrews et al. report protection against death for the first dose of Pfizer as 88.6% (77.3 to 94.3%), at least 28 days following the first dose and up to the second dose if given. This is a **21.2% increase** compared to their equivalent estimate for Alpha.<br>**Alpha assumption 0.85**<br>**0.92 (+28 days) as for hospitalisation** |
| Overall protection against onward transmission for Pfizer dose 1 | Eyre et al. report an adjusted odds ratio of 0.87 (0.81, 0.94) for the effect of a case being partially vaccinated with Pfizer (dose 1 day 1 to dose 2 +14 days) compared to an unvaccinated case in relation to the likelihood of a contact testing PCR-positive, equivalent to vaccine protection of 13%. Their equivalent estimate for Alpha is 0.74 (0.70, 0.80), therefore vaccine protection of 26%. This is a **50% overall reduction** in vaccine effect from Alpha to Delta.<br>**Alpha assumption 0.47**<br>**0.24 = 0.47 \* (1 - 0.5) (+28 days)** |
| Overall protection against infection for Pfizer dose 2 | Pouwels et al. report vaccine effectiveness against all PCR-confirmed infections with the Delta variant of 80% (77-83%) at least 14 days following the second dose of Pfizer. This is a **2.56% increase** on their equivalent estimate for the Alpha variant. Sheikh et al. report vaccine effectiveness against PCR-confirmed infection (regardless of symptom status) of 79% (75-82%) 14 days after the second dose of Pfizer. This is a **14.13% reduction** on their equivalent estimate for the Alpha variant.<br>**Alpha assumption 0.85**<br>**0.8 = 0.85 \* (1-0.057) (+14 days)** |
| Overall protection against disease for Pfizer dose 2 | Lopez Bernal et al. C report adjusted vaccine effectiveness against symptomatic infection with S-gene target positives (Delta variant) of 88.0% (85.3% to 90.1%) at least 14 days after the second dose of Pfizer. A **6.08% reduction** on their estimate of equivalent vaccine protection against the Alpha variant. Pouwels et al. report vaccine effectiveness against symptomatic PCR-confirmed infections with the Delta variant of 84% (82-86%) at least 14 days following the second dose of Pfizer. This is a **13.4% reduction** on their equivalent estimate for the Alpha variant. Sheikh et al. report vaccine effectiveness against symptomatic PCR-confirmed infection of 83% (78-87%) 14 days after the second dose of Pfizer. This is a **9.78% reduction** on their equivalent estimate for the Alpha variant. PHE's week 36 vaccine surveillance report estimates protection against symptomatic disease of 79% (78-80%) for the Delta variant at least 14 days after a second vaccine dose. This is an **11.24% reduction** compared to their equivalent estimate for Alpha. Andrews et al. report protection against symptomatic disease for the second dose of Pfizer as 83.5% (83.3 to 83.6%), at least 14 days following the second dose. This is a **12.1% reduction** compared to their equivalent estimate for Alpha.<br>**Alpha assumption 0.9**<br>**$0.81 = 0.9 * (1 - 0.105) (+14 \text{ days})$** |
| Overall protection against hospitalisation for Pfizer dose 2 | Stowe et al. report vaccine effectiveness against hospitalisation of 96% (86-99%) following the second dose of Pfizer. This is a **1.05% increase** on their equivalent estimate for the Alpha variant. PHE's week 36 vaccine surveillance report estimates protection against hospitalisation of 96% (91-98%) for the Delta variant at least 14 days after a second vaccine dose. This is a **3.23% increase** compared to their equivalent estimate for Alpha. Andrews et al. report protection against hospitalisation for the second dose of Pfizer as 96.7% (96.3 to 97%), at least 14 days following the second dose. This is a **1.2% reduction** compared to their equivalent estimate for Alpha.<br>**Alpha assumption 0.95**<br>**$0.96 = 0.95 * (1 + 0.0103) (+14 \text{ days})$** |
| Overall protection against mortality for Pfizer dose 2 | Andrews et al. report protection against death for the second dose of Pfizer as 95.2% (93.7 to 96.4%), at least 14 days following the second dose. This is a **1.1% reduction** compared to their equivalent estimate for Alpha.<br>**Alpha assumption 0.95**<br>**0.96 (+14 days) as for hospitalisation** |
| Overall protection against onward transmission for Pfizer dose 2 | Eyre et al. report an adjusted odds ratio of 0.35 (0.26, 0.48) for the effect of a case being fully vaccinated with Pfizer (dose 2 +14 days) compared to an unvaccinated case in relation to the likelihood of a contact testing PCR-positive, equivalent to vaccine protection of 65%. Their equivalent estimate for Alpha is 0.18 (0.12, 0.29), therefore vaccine protection of 82%. This is a **20.7% overall reduction** in vaccine effect from Alpha to Delta.<br>**Alpha assumption 0.47**<br>**$0.37 = 0.47 * (1 - 0.207) (+14 \text{ days})$** |

### Working group authors and acknowledgements

#### CMMID COVID-19 working group

The following authors were part of the Centre for Mathematical Modelling of Infectious Disease COVID-19 working group. Each contributed in processing, cleaning and interpretation of data, interpreted findings, contributed to the manuscript, and approved the work for publication: James D Munday, Rachel Lowe, Gwenan M Knight, Quentin J Leclerc, Damien C Tully, David Hodgson, Rachael Pung, Joel Hellewell, Mihaly Koltai, David Simons, Kaja Abbas, Adam J Kucharski, Simon R Procter, Frank G Sandmann, Carl A B Pearson, Kiesha Prem, Alicia Showering, Sophie R Meakin, Kathleen O’Reilly, Ciara V McCarthy, Matthew Quaife, Kerry LM Wong, Yalda Jafari, Arminder K Deol, Rein M G J Houben, Charlie Diamond, Thibaut Jombart, C Julian Villabona-Arenas, William Waites, Rosalind M Eggo, Akira Endo, Hamish P Gibbs, Petra Klepac, Jack Williams, Billy J Quilty, Oliver Brady, Jon C Emery, Katherine E Atkins, Lloyd A C Chapman, Katharine Sherratt, Sam Abbott, Nikos I Bosse, Paul Mee, Sebastian Funk, Jiayao Lei, Yang Liu, Stefan Flasche, James W Rudge, Fiona Yueqian Sun, Graham Medley, Timothy W Russell, Amy Gimma, Stéphane Hué, Christopher I Jarvis, Emilie Finch and Samuel Clifford.

#### CMMID COVID-19 working group funding statements

PK (Royal Society: RP\EA\180004, European Commission: 101003688), TJ (Global Challenges Research Fund: ES/P010873/1, UK Public Health Rapid Support Team, NIHR: Health Protection Research Unit for Modelling Methodology HPRU-2012-10096, UK MRC: MC_PC_19065), AG (European Commission: 101003688), SFlasche (Wellcome Trust: 208812/Z/17/Z), RL (Royal Society: Dorothy Hodgkin Fellowship), DH (NIHR: 1R01AI141534-01A1), AJK (Wellcome Trust: 206250/Z/17/Z, NIHR: NIHR200908), HPG (EDCTP2: RIA2020EF-2983-CSIGN, UK DHSC/UK Aid/NIHR: PR-OD-1017-20001), SFunk (Wellcome Trust: 210758/Z/18/Z), GMK (UK MRC: MR/P014658/1), WW (MRC: MR/V027956/1), RME (HDR UK: MR/S003975/1, UK MRC: MC_PC_19065, NIHR: NIHR200908), MK (Wellcome Trust: 221303/Z/20/Z), LACC (NIHR: NIHR200908), YL (B&MGF: INV-003174, NIHR: 16/137/109, European Commission: 101003688, UK MRC: MC_PC_19065), KA (BMGF: INV-016832; OPP1157270), KO’R (B&MGF: OPP1191821), KEA (ERC: SG 757688), FYS (NIHR: 16/137/109), FGS (NIHR: NIHR200929), RP (Singapore Ministry of Health), PM CADDE (MR/S0195/1 & FAPESP 18/14389-0), SC (Wellcome Trust: 208812/Z/17/Z, UK MRC: MC_PC_19065), SA (Wellcome Trust: 210758/Z/18/Z), CIJ (Global Challenges Research Fund: ES/P010873/1), CVM (NIHR: NIHR200929), NIB (HPRU: NIHR200908), SRP (B&MGF: INV-016832), KLM (European Commission: 101003688), JH (Wellcome Trust: 210758/Z/18/Z), YJ (UKRI: MR/V028456/1), MQ (ERC Starting Grant: #757699, B&MGF: INV-001754), OJB (Wellcome Trust: 206471/Z/17/Z), JDM (Wellcome Trust: 210758/Z/18/Z), TWR (Wellcome Trust: 206250/Z/17/Z), KP (B&MGF: INV-003174, European Commission: 101003688), BJQ (NIHR: 16/137/109, NIHR: 16/136/46, B&MGF: OPP1139859), SRM (Wellcome Trust: 210758/Z/18/Z), GFM (B&MGF: NTD Modelling Consortium OPP1184344), EF (MRC: MR/N013638/1), CJVA (ERC: SG 757688), CABP (B&MGF: NTD Modelling Consortium OPP1184344, FCDO/Wellcome Trust: Epidemic Preparedness Coronavirus research programme 221303/Z/20/Z), AE (Nakajima Foundation), JCE (ERC Starting Grant: #757699), QJL (UK MRC: LID DTP MR/N013638/1), RMGJH (ERC Starting Grant: #757699), CD (NIHR: 16/137/109), JYL (B&MGF: INV-003174), KS (Wellcome Trust: 210758/Z/18/Z), DS (BBSRC LIDP: BB/M009513/1), JWR (DTRA: HDTRA1-18-1-0051).

## Acknowledgements

The authors would like to thank Stephen J. Rivers, Lloyd A. C. Chapman and Ciara V. McCarthy for helpful discussions which have contributed towards completion of this work.

## Notes

### Summary of Updates

The manuscript and analysis has been updated following the emergence and spread of the Omicron B.1.1.529 SARS-CoV-2 variant in England.

## References

1. COVID-19 Data Explorer. Our World in Data https://ourworldindata.org/coronavirus-data-explorer.

2. UK Summary. https://coronavirus.data.gov.uk.

3. Tracking SARS-CoV-2 variants. https://www.who.int/activities/tracking-SARS-CoV-2-variants.

4. Website. https://www.instituteforgovernment.org.uk/charts/uk-government-coronavirus-lockdowns.

5. Prime Minister’s Office,. PM statement on living with COVID: 21 February 2022. GOV.UK https://www.gov.uk/government/speeches/pm-statement-on-living-with-covid-21-february-2022 (2022).

6. Medicines and Healthcare products Regulatory Agency. Valneva COVID-19 vaccine approved by MHRA. GOV.UK https://www.gov.uk/government/news/valneva-covid-19-vaccine-approved-by-mhra (2022).

7. Joint Committee on Vaccination and Immunisation: advice on priority groups for COVID-19 vaccination, 30 December 2020. https://www.gov.uk/government/publications/priority-groups-for-coronavirus-covid-19-vaccination-advice-from-the-jcvi-30-december-2020/joint-committee-on-vaccination-and-immunisation-advice-on-priority-groups-for-covid-19-vaccination-30-december-2020.

8. Department of Health and Social Care. All young people aged 16 and 17 in England to be offered vaccine by next week. GOV.UK https://www.gov.uk/government/news/all-young-people-aged-16-and-17-in-england-to-be-offered-vaccine-by-next-week (2021).

9. England, N. H. S. NHS England » NHS rolls out COVID vaccine to five million 5 to 11 year olds. https://www.england.nhs.uk/2022/04/nhs-rolls-out-covid-vaccine-to-five-million-5-to-11-year-olds/.

10. UK Health Security Agency. JCVI issues advice on COVID-19 booster vaccines for those aged 40 to 49 and second doses for 16 to 17 year olds. GOV.UK https://www.gov.uk/government/news/jcvi-issues-advice-on-covid-19-booster-vaccines-for-those-aged-40-to-49-and-second-doses-for-16-to-17-year-olds (2021).

11. Classification of Omicron (B.1.1.529): SARS-CoV-2 Variant of Concern. https://www.who.int/news/item/26-11-2021-classification-of-omicron-(b.1.1.529)-sars-cov-2-variant-of-concern.

12. Department of Health and Social Care. First UK cases of Omicron variant identified. GOV.UK https://www.gov.uk/government/news/first-uk-cases-of-omicron-variant-identified (2021).

13. Andrews, N. et al. Covid-19 Vaccine Effectiveness against the Omicron (B.1.1.529) Variant. N. Engl. J. Med. 386, (2022).

14. Department of Health and Social Care. JCVI advice on the UK vaccine response to the Omicron variant. GOV.UK https://www.gov.uk/government/publications/uk-vaccine-response-to-the-omicron-variant-jcvi-advice/jcvi-advice-on-the-uk-vaccine-response-to-the-omicron-variant (2021).

15. JCVI statement on the adult COVID-19 booster vaccination programme and the Omicron variant: 7 January 2022. GOV.UK https://www.gov.uk/government/publications/jcvi-statement-on-the-adult-covid-19-booster-vaccination-programme-and-the-omicron-variant/jcvi-statement-on-the-adult-covid-19-booster-vaccination-programme-and-the-omicron-variant-7-january-2022.

16. JCVI statement on COVID-19 vaccination of children and young people: 22 December 2021. GOV.UK https://www.gov.uk/government/publications/jcvi-update-on-advice-for-covid-19-vaccination-of-children-and-young-people/jcvi-statement-on-covid-19-vaccination-of-children-and-young-people-22-december-2021.

17. England, N. H. S. NHS England » NHS expands COVID vaccinations to the most vulnerable 5 to 11 year olds. https://www.england.nhs.uk/2022/01/nhs-expands-covid-vaccinations-to-the-most-vulnerable-5-to-11-year-olds/.

18. JCVI statement on vaccination of children aged 5 to 11 years old. GOV.UK https://www.gov.uk/government/publications/jcvi-update-on-advice-for-covid-19-vaccination-of-children-aged-5-to-11/jcvi-statement-on-vaccination-of-children-aged-5-to-11-years-old.

19. Andrews, N. et al. Duration of Protection against Mild and Severe Disease by Covid-19 Vaccines. N. Engl. J. Med. 386, (2022).

20. UK Health Security Agency. COVID-19 vaccine weekly surveillance reports (weeks 39 to 42). https://www.gov.uk/government/publications/covid-19-vaccine-weekly-surveillance-reports (2021).

21. Google. COVID-19 Community Mobility Reports.

22. Davies, N. G. et al. Association of tiered restrictions and a second lockdown with COVID-19 deaths and hospital admissions in England: a modelling study. The Lancet Infectious Diseases vol. 21 482–492 (2021).

23. Attendance in education and early years settings during the coronavirus (COVID-19) pandemic. https://explore-education-statistics.service.gov.uk/find-statistics/attendance-in-education-and-early-years-settings-during-the-coronavirus-covid-19-outbreak/2022-week-16.

24. Statistics. Statistics » COVID-19 Vaccinations. https://www.england.nhs.uk/statistics/statistical-work-areas/covid-19-vaccinations/.

25. England, N. H. S. NHS England » Regional teams. https://www.england.nhs.uk/about/regional-area-teams/.

26. Website. https://doi.org/10.1016/S0140-6736(21)01694-9 doi:10.1016/S0140-6736(21)01694-9.

27. Yapp, R. & Willis, Z. Coronavirus (COVID-19) Infection Survey: England. (2022).

28. Department of Health and Social Care. England to return to Plan A following the success of the booster programme. *GOV.UK* https://www.gov.uk/government/news/england-to-return-to-plan-a-following-the-success-of-the-booster-programme (2022).

29. Davies, N. G. et al. Age-dependent effects in the transmission and control of COVID-19 epidemics. Nat. Med. 26, 1205–1211 (2020).

30. Davies, N. G. et al. Increased mortality in community-tested cases of SARS-CoV-2 lineage B.1.1.7. Nature 593, 270–274 (2021).

31. Public Health England. Investigation of SARS-CoV-2 variants of concern: technical briefings. https://www.gov.uk/government/publications/investigation-of-novel-sars-cov-2-variant-variant-of-concern-20201201 (2020).

32. Nyberg, T. et al. Comparative analysis of the risks of hospitalisation and death associated with SARS-CoV-2 omicron (B.1.1.529) and delta (B.1.617.2) variants in England: a cohort study. Lancet 399, (2022).

33. Sigal, A., Milo, R. & Jassat, W. Estimating disease severity of Omicron and Delta SARS-CoV-2 infections. Nat. Rev. Immunol. 22, 267–269 (2022).

34. Teslya, A. et al. Impact of self-imposed prevention measures and short-term government-imposed social distancing on mitigating and delaying a COVID-19 epidemic: A modelling study. PLoS Med. 17, e1003166 (2020).

35. Weitz, J. S., Park, S. W., Eksin, C. & Dushoff, J. Awareness-driven behavior changes can shift the shape of epidemics away from peaks and toward plateaus, shoulders, and oscillations. Proc. Natl. Acad. Sci. U. S. A. 117, 32764–32771 (2020).

36. UK Health Security Agency. Investigation of SARS-CoV-2 variants: technical briefings. GOV.UK https://www.gov.uk/government/publications/investigation-of-sars-cov-2-variants-technical-briefings (2021).

37. Bonnie Lewis, Ed Pyle, Matt Dennes, Tim Vizard. Coronavirus and the social impacts on Great Britain - Office for National Statistics. https://www.ons.gov.uk/peoplepopulationandcommunity/healthandsocialcare/healthandwellbeing/bulletins/coronavirusandthesocialimpactsongreatbritain/1april2022 (2022).

38. UK Health Security Agency. JCVI advises a spring COVID-19 vaccine dose for the most vulnerable. GOV.UK https://www.gov.uk/government/news/jcvi-advises-a-spring-covid-19-vaccine-dose-for-the-most-vulnerable (2022).

39. Lyngse, F. P., et al. Transmission of SARS-CoV-2 Omicron VOC subvariants BA.1 and BA.2: Evidence from Danish Households. medRxiv 2022.01.28.22270044 (2022).

40. Stegger, M., et al. Occurrence and significance of Omicron BA.1 infection followed by BA.2 reinfection. medRxiv 2022.02.19.22271112 (2022).

41. Davies, N. G. et al. Estimated transmissibility and impact of SARS-CoV-2 lineage B.1.1.7 in England. Science 372, (2021).

42. Scientific Advisory Group for Emergencies. LSHTM: Updated roadmap assessment – prior to delayed Step 4, 7 July 2021. https://www.gov.uk/government/publications/lshtm-updated-roadmap-assessment-prior-to-delayed-step-4-7-july-2021 (2021).

43. Scientific Advisory Group for Emergencies. LSHTM: Interim roadmap assessment – prior to steps 3 and 4, 5 May 2021. GOV.UK https://www.gov.uk/government/publications/lshtm-interim-roadmap-assessment-prior-to-steps-3-and-4-5-may-2021 (2021).

44. Barnard, R. C., Davies, N. G., Pearson, C. A. B., Jit, M. & John Edmunds, W. Projected epidemiological consequences of the Omicron SARS-CoV-2 variant in England, December 2021 to April 2022. medRxiv 2021.12.15.21267858 (2021).

45. Scientific Advisory Group for Emergencies. LSHTM: autumn and winter scenarios 2021 to 2022, 13 October 2021. GOV.UK https://www.gov.uk/government/publications/lshtm-autumn-and-winter-scenarios-2021-to-2022-13-october-2021 (2021).

46. COVID-19 Genomic Surveillance – Wellcome Sanger Institute. https://covid19.sanger.ac.uk/lineages/raw.

47. Ito, K., Piantham, C. & Nishiura, H. Estimating relative generation times and relative reproduction numbers of Omicron BA.1 and BA.2 with respect to Delta in Denmark. medRxiv 2022.03.02.22271767 (2022).

48. Yamasoba, D. et al. Virological characteristics of SARS-CoV-2 BA.2 variant. bioRxiv 2022.02.14.480335 (2022) doi:10.1101/2022.02.14.480335.

49. Lentini, A., Pereira, A., Winqvist, O. & Reinius, B. Monitoring of the SARS-CoV-2 Omicron BA.1/BA.2 variant transition in the Swedish population reveals higher viral quantity in BA.2 cases. medRxiv 2022.03.26.22272984 (2022).

50. Statement on Omicron sublineage BA.2. https://www.who.int/news/item/22-02-2022-statement-on-omicron-sublineage-ba.2.

51. Kirsebom, F. C. M., et al. COVID-19 Vaccine Effectiveness against the Omicron BA.2 variant in England. medRxiv 2022.03.22.22272691 (2022).

52. COVID-19 hub. https://www.ukbiobank.ac.uk/learn-more-about-uk-biobank/covid-19-hub.

53. Real-time Assessment of Community Transmission findings. https://www.imperial.ac.uk/medicine/research-and-impact/groups/react-study/real-time-assessment-of-community-transmission-findings/.

54. Hill, R. Y. A. Coronavirus (COVID-19) Infection Survey, antibody and vaccination data, UK. (2021).

55. Levin, A. T. et al. Assessing the Age Specificity of Infection Fatality Rates for COVID-19: Systematic Review, Meta-Analysis, and Public Policy Implications. medRxiv 2020.07.23.20160895 (2020).

56. Scientific Advisory Group for Emergencies. Imperial College London: Omicron severity and vaccine effectiveness, 5 January 2022. GOV.UK https://www.gov.uk/government/publications/imperial-college-london-omicron-severity-and-vaccine-effectiveness-5-january-2022 (2022).

57. Report 50 - Hospitalisation risk for Omicron cases in England. Imperial College London http://www.imperial.ac.uk/medicine/departments/school-public-health/infectious-disease-epidemiology/mrc-global-infectious-disease-analysis/covid-19/report-50-severity-omicron/.

58. Mossong, J. et al. Social contacts and mixing patterns relevant to the spread of infectious diseases. PLoS Med. 5, e74 (2008).

59. Braak, C. J. F. T. A Markov Chain Monte Carlo version of the genetic algorithm Differential Evolution: easy Bayesian computing for real parameter spaces. Stat. Comput. 16, 239–249 (2006).

60. Endo, A., van Leeuwen, E. & Baguelin, M. Introduction to particle Markov-chain Monte Carlo for disease dynamics modellers. Epidemics 29, 100363 (2019).

61. Khoury, D. S. et al. Neutralizing antibody levels are highly predictive of immune protection from symptomatic SARS-CoV-2 infection. Nat. Med. 27, 1205–1211 (2021).

62. Cromer, D. et al. Neutralising antibody titres as predictors of protection against SARS-CoV-2 variants and the impact of boosting: a meta-analysis. The Lancet. Microbe 3, (2022).

63. Hyndman, R. J. & Khandakar, Y. Automatic Time Series Forecasting: The forecast Package for R. J. Stat. Softw. 27, 1–22 (2008).

64. Public Health England. JCVI advises on COVID-19 vaccine for people aged under 40. GOV.UK https://www.gov.uk/government/news/jcvi-advises-on-covid-19-vaccine-for-people-aged-under-40 (2021).

65. COVID-19 Genomic Surveillance – Wellcome Sanger Institute. https://covid19.sanger.ac.uk/downloads.

66. Willis, K. S. A. Coronavirus (COVID-19) Infection Survey: England. (2021).

67. Lauer, S. A. et al. The Incubation Period of Coronavirus Disease 2019 (COVID-19) From Publicly Reported Confirmed Cases: Estimation and Application. Ann. Intern. Med. 172, 577–582 (2020).

68. Ferretti, L. et al. The timing of COVID-19 transmission. Epidemiology (2020).

69. Li, Q. et al. Early Transmission Dynamics in Wuhan, China, of Novel Coronavirus-Infected Pneumonia. N. Engl. J. Med. 382, 1199–1207 (2020).

70. Nishiura, H., Linton, N. M. & Akhmetzhanov, A. R. Serial interval of novel coronavirus (COVID-19) infections. Int. J. Infect. Dis. 93, 284–286 (2020).

71. Bi, Q. et al. Epidemiology and transmission of COVID-19 in 391 cases and 1286 of their close contacts in Shenzhen, China: a retrospective cohort study. Lancet Infect. Dis. 20, 911–919 (2020).

72. Website. https://doi.org/10.1016/S2468-2667(20)30133-X doi:10.1016/S2468-2667(20)30133-X.

73. Population estimates for the UK, England and Wales, Scotland and Northern Ireland - Office for National Statistics. https://www.ons.gov.uk/peoplepopulationandcommunity/populationandmigration/populationestimates/bulletins/annualmidyearpopulationestimates/mid2018.

74. Docherty, A. B. et al. Features of 20 133 UK patients in hospital with covid-19 using the ISARIC WHO Clinical Characterisation Protocol: prospective observational cohort study. BMJ 369, (2020).

75. Optimising the COVID-19 vaccination programme for maximum short-term impact. GOV.UK https://www.gov.uk/government/publications/prioritising-the-first-covid-19-vaccine-dose-jcvi-statement/optimising-the-covid-19-vaccination-programme-for-maximum-short-term-impact.

